# Measuring aversion to health inequality in Canada: an equity-efficiency trade-off experiment

**DOI:** 10.1101/2024.09.27.24314482

**Authors:** Nicolas Iragorri, Shehzad Ali, Sharmistha Mishra, Beate Sander

## Abstract

**OBJECTIVES:** To estimate the extent to which Canadians are averse to health inequalities, a critical component for equity-informative economic evaluations but lacking in the Canadian context.

**METHODS:** We conducted three experiments among a representative sample of adults living in Canada to elicit value judgements about reducing income-related health inequality vs. improving population health. Each experiment compared two programs: (Experiment 1) universal and tailored vaccination; (Experiment 2) non-specific prevention programs; (Experiment 3) generic health care programs. The programs varied in terms of efficiency (additional life years), and income-related health inequality. Preferences were elicited using benefit-trade off analysis and were classified as: pro-rich (maximizing the health of individuals with the highest income); health maximizer (maximizing total health); weighted prioritarian (willing to trade some health to reduce inequalities); maximin (only improving the health of the individuals with the lowest income); and egalitarian (minimizing health inequalities).

**RESULTS:** We recruited 1,000 participants per experiment. Preferences for the vaccination, prevention, and generic experiments were distributed as follows: pro-rich (Atkinson Index<0): 31%, 22%, and 16% respectively; health maximizers (Atkinson Index=0): 2%, 3%, and 2%, respectively; weighted prioritarians (Atkinson Index>0): 13%, 19%, and 22% respectively; maximins (Atkinson Index=∞): 0%, 1%, and 3%, respectively; egalitarian (Atkinson Index undefined): 54%, 55%, and 57%, respectively. The median responses reflected a preference for minimizing health inequalities across the three experiments.

**CONCLUSIONS:** Our findings suggest a strong aversion to health inequality among people living in Canada with over half of respondents willing to minimize health inequalities regardless of the impact on efficiency.

## 1. Introduction

Economic evaluations of health interventions are important tools for health technology assessment (HTA) to guide decision making (1). While health agencies recommend employing frameworks that include health equity as a fundamental dimension to decision making (2), conventional economic evaluations aim to maximize overall health outcomes at a given budget, without explicitly considering distributional or equity effects (3–5). Recent developments in economic evaluation have advocated for Equity-Informative Economic Evaluations (EIEEs) to enable the systematic consideration of health equity effects in economic evaluations (6). EIEEs require understanding population preferences for reducing disparities and health inequality aversion (7), which can be elicited using equity-efficiency trade-off studies (7). However, preferences may vary across countries and the populations experiencing inequalities (8). Thus, measurement of health inequality aversion within the specific setting in which a health technology will be assessed is essential for robust, context-specific economic evaluations (9).

Canadian HTA Agencies, such as Canada’s Drug Agency and the National Advisory Committee on Immunization (NACI), have highlighted the importance of considering health equity when designing economic evaluations, with NACI explicitly recommending EIEEs (10,11). However, comprehensive data on societal health inequality aversion is lacking, a major obstacle to conducting EIEEs in Canada (12). A recent review of equity-efficiency trade-off studies found that 70% of studies were conducted in Europe (8). Only one Canadian study was identified (13). Social welfare functions were the most common methodologies used to quantify health inequality aversion; over 60% of studies estimated Atkinson inequality aversion indices. Overall, results indicated strong aversion to health inequality in the UK, continental Europe, and in the US (8). Canadian evidence was inconclusive due to a single trade-off scenario design (13).

Equity-efficiency trade-offs are commonly assessed using benefit trade-off experiments, a survey method that measures individual preferences (8). With this approach, participants are asked to evaluate hypothetical health programs that differ in their impact on overall health and health-related inequalities. These experiments have often compared abstract interventions (e.g., program A versus program B) to estimate inequality aversion while avoiding language and themes that might affect responses due to unobserved cognitive effects (14). However, research indicates that using concrete scenarios lowered the proportion of extreme inequality aversion in the UK, when compared to abstract interventions (14). These comparisons have yet to be conducted in Canada. Therefore, our main objectives were to elicit the level of health inequality aversion among a representative sample of the Canadian adult population, and to compare health inequality aversion across three main types of health programs, ranging from specific to abstract: (i) vaccination; (ii) prevention; and (iii) abstract.

## 2. Methods

### 2.1. Study design

We conducted a benefit trade-off experiment to elicit health inequality aversion in Canada. We provided respondents with a survey that described the difference in life expectancy between populations in the highest and the lowest household income quintiles. Preferences were elicited based on the respondents’ choice of hypothetical public health programs that differed in their impact on health-adjusted life expectancy for population groups in pairwise comparisons. Atkinson inequality indices were mathematically derived to determine the magnitude of the health equity-efficiency trade-off based on participant responses. We followed the consensus-based checklist for reporting of survey studies (15).

We adapted the experiment from Ali et al. (14) and developed different versions for three health programs, ranging from concrete to generic. The first version compared universal and tailored vaccination programs. Universal vaccination assumed that vaccines for a generic respiratory disease were offered to everyone in Canada. A tailored vaccination program was defined as a policy designed to prioritize vaccine uptake among populations in the lowest household income quintile, because (i) members of low-income households might be at higher risk of infection (16); (ii) members of low-income households may face barriers to access and uptake of the vaccine (16). Therefore, tailored programs were designed to reach people regardless of their socioeconomic status, although greater efforts were placed to improve access for individuals from lower income households. The second experiment compared broad prevention programs (i.e., not specific to a condition): (i) universal prevention program and (ii) tailored prevention program. The third and last version compared generic programs (program A vs. program B). All interventions across the three experiments were assumed to cost the same to implement, i.e., were cost neutral across options.

### 2.2. Recruitment and administration

We surveyed a sample of the Canadian general population employing convenience sampling methods through AskingCanadians (17), a third-party recruitment company. We recruited, registered and consented members of AskingCanadians’ research panel who are people living in Canada, 18 years or older, and who opted-in to participate in online surveys. To ensure representativeness, AskingCanadians employs sampling profiling variables which are updated monthly and monitored against Statistics Canada data (17). Personally identifiable information was not collected. Research panel members were presented with a consent letter which explained that all participation was voluntary and confidential, and withdrawal was permitted at any point throughout the survey. Details on compensation and conflicts of interest were also provided. The study was approved by the University of Toronto Research Ethics Board.

AskingCanadians randomly selected research panel members. To reduce sampling bias and maximize population representativeness, we used quotas for stratified sampling based on region (i.e., Province/Territory), interlocked by age, gender, and household income categories employed by Statistics Canada (Appendix 1). The quotas included a +/- 5% variance and were balanced against Statistics Canada norms (17). We aimed to enroll 1,000 respondents per survey (3,000 total), to ensure 450 valid responses per survey, based on sample size calculations of published trade-off experiments (18). Respondents could only participate in one of the three surveys to avoid priming effects and response fatigue (19,20). Respondents could choose between French and English versions of the surveys.

### 2.3. Equity-efficiency trade-off experiment

Asking Canadians collected the following sociodemographic information, following Statistics Canada and Public Health Ontario definitions: gender (21), household income before taxes (22), ethnicity/ancestry (23), racial origin/lineage (22), and the highest level of education attainment (24). Age was captured as an open-ended question. We introduced the trade-off experiment, from which population preferences were elicited, by describing the baseline difference in life expectancy between populations in the highest and lowest household income quintiles in Canada. Individuals in the lowest household income quintile (mean annual household income $25,000) (25) have a life expectancy at birth of 68 years of life in full health, compared to 79 years for individuals in the highest household income quintile (mean household income $160,000) (25,26), which represents a baseline health gap of 11 years.

We then presented two hypothetical public health programs. Each was assumed to affect the life expectancy of both population groups. For the first experiment, we introduced two distinct vaccination strategies (universal versus tailored vaccination) for an undefined respiratory infectious disease. Although both programs improved overall health, the distribution of life-years gained differed across the populations. The distribution of health gains was informed by Ali et al. (27) and is summarized in (Appendix 2). U*niversal vaccination* represented seven additional years per person among the population in the highest household income quintile, and three additional years per person among the population in the lowest household income quintile. *Tailored vaccination* was associated with a three-year increase among those in the highest household income quintile, and an eight-year increase among those in the lowest quintile. We used the same distribution of health gains for the prevention and generic program experiments. We asked respondents to choose the program that the Canadian government should fund, based on the resulting health distribution and the health gap for each scenario (i.e., the equity-efficiency trade-off), assuming both programs cost the same. Respondents could also indicate that both programs were equally good.

Eight additional scenarios were presented within the same experiment, where the first intervention (e.g., *universal vaccination)* remained unaltered, and the health gains for the population in the lowest household income quintile in the alternative program (e.g., *tailored vaccination)* were reduced by one year in each subsequent scenario. Respondents were asked to choose between Universal (U), Tailored (T), or equally good (=). Logic checks were conducted by including a question that was asked twice (once in the introduction, and once among the trade-off scenarios) to check whether respondents could interpret the graphs and the survey questions. The complete questionnaires are available in Appendices 3-5.

### 2.4. Response classification and parameter elicitation (Base case)

We categorized the responses based on the classification system developed by Ali et al. (14). For example, if a respondent preferred *universal vaccination (U)* over *tailored vaccination (T)* across the eight scenarios, their response would be classified as UUUUUUUU. If a respondent chose *tailored vaccination* for the first four scenarios and switched to *universal vaccination* in the fifth, the response would be categorized as TTTTUUUU, or TTTT=UUU if the respondent was indifferent in the fifth scenario. The indifference point was identified as the scenario in which respondents chose ‘*U and T are equally good’*, or when they switched directly between T and U. Respondents who switched more than once between programs were not included in the base case analysis, as the estimation of the Atkinson Index requires a unique indifference point per respondent (i.e., logical response).

A total of 17 potential logical responses were categorized as per Ali et al. (14), and summarized in Table 1: (i) ‘pro-rich’, comprised of respondents who had a stronger preference for improving the health of the highest income quintile population. This group was further divided into three separate categories based on preference strength; (ii) ‘health maximizers’ included those who chose the program that maximized total health, regardless of the distribution; (iii) ‘weighted prioritarians’ (7 categories), were the respondents who were willing to forgo health in order to achieve a more equitable distribution; (iv) ‘maximins’, comprised of individuals only concerned with improving the health of the population in the lowest household income quintile; (v) egalitarians (5 categories), those who were willing to forgo health benefits even from those in the lower household income quintiles to reduce health inequality. Finally, we stratified the preference distributions by sociodemographic characteristics: age, gender, province, annual household income, racial origin, ethnicity/ancestry, and highest level of education attainment.

**Table 1:**
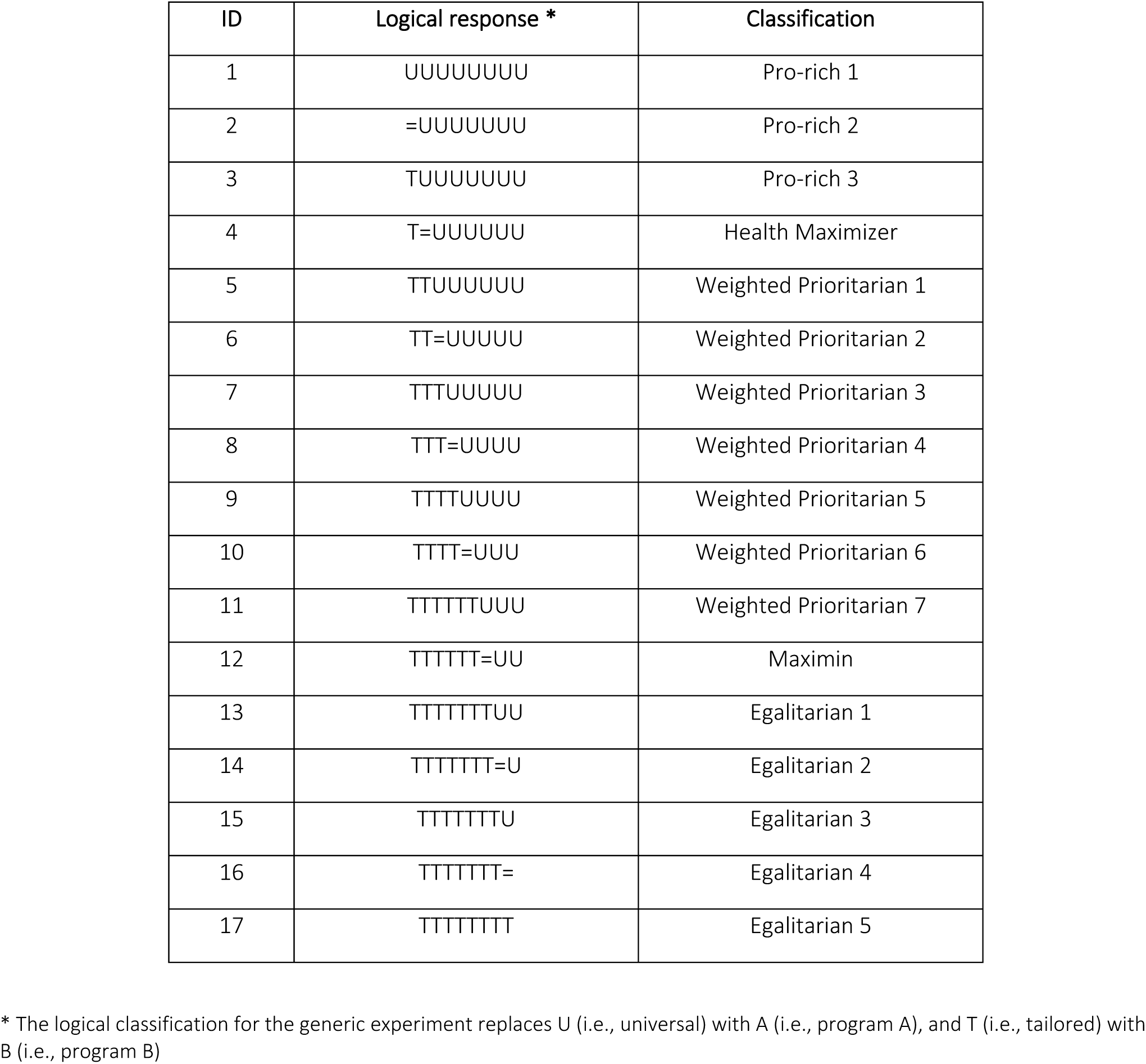
Classification system for logical responses.

We identified the indifference point in the responses to estimate the inequality aversion parameters. To do so, we computed the equally distributed equivalent (EDE) levels of health from the Atkinson Index, such that:

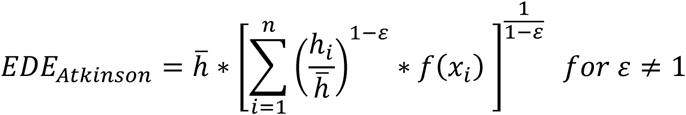

Where:

*i* = subgroup i(i.e., high or low income households),

*h_i_* = life expectancy of subgroup i,

*h*^-^ = average health of the population

*f(x_i_)* = proportion of individuals in subgroub i

ε = inequality aversion parameter

The EDE level of health represents the mean health-related social welfare of the hypothetical population considered in the trade-off experiments, comprised of the highest and lowest household income quintiles. The EDEs for these two populations are mathematically equivalent at the mean indifference point, representing a system of two equations (one EDE for each population group) with one unknown parameter (ε). When both equations are equal, the system can be algebraically solved to derive the inequality parameter ε. We estimated inequality aversion parameters for each response classification category with the Solver function in Visual Basics for Applications in MS Excel and estimated the median Atkinson index for each of the three experiments. Since the Atkinson index cannot be easily interpreted on its own, we estimated the isoelastic transformation of the EDE function to derive the marginal social value of a change in health (28) (i.e., how each additional life year accrued by individuals in the lowest household income quintile is valued relative to those in the highest quintile).

### 2.5. Response classification (sensitivity analysis)

A sensitivity analysis was conducted to include responses that did not meet the logical response definition. For responses with more than one switch, we assumed that the first switch represented the indifference point. For example, the response TTT=UUUT would not have been included in the base case but would have been modified to TTT=UUUU and included in the sensitivity analysis.

## 3. Results

### 3.1. Recruitment and sociodemographic characteristics

AskingCanadians approached a total of 33,286 panel members for the three experiments, of which 22,036 (66%) were excluded to avoid oversampling for specific populations and ensure representativeness of the Canadian population by region, age, gender, and income. Further, 6,804 (20%) individuals were screened out due to failed logic and graph literacy checks, and 1,902 (6%) voluntarily withdrew from completing the surveys. The experiments were completed by 3,000 individuals (9%). The median time to completion was 7.1 minutes per respondent.

Sociodemographic characteristics are summarized in Table 2 for each experiment. Overall, most respondents resided in Ontario (39%), Quebec (23%), British Columbia (13%), and Alberta (11%). The mean age was 48.6 years (SD=16.3 years), and approximately 51% identified as female. A total of 80% of questionnaires were completed in English. The three most often reported ethnicity/ancestry categories were European origins (57%), Asian origins (17%), and North American Aboriginal origins (11%). Most respondents identified their racial origin/lineage as White (74%), East Asian (9%), and South Asian (4%). The most common levels of education attainment were university certificate or diploma at bachelor level or above (35%), non-university certificate (24%), and secondary school diploma (22%). The most common categories for annual household income after tax were between $0 and $29,999 (36%), between $30,000 and $49,999 (21%), and between $50,000 and $69,999 (14%).

**Table 2:**
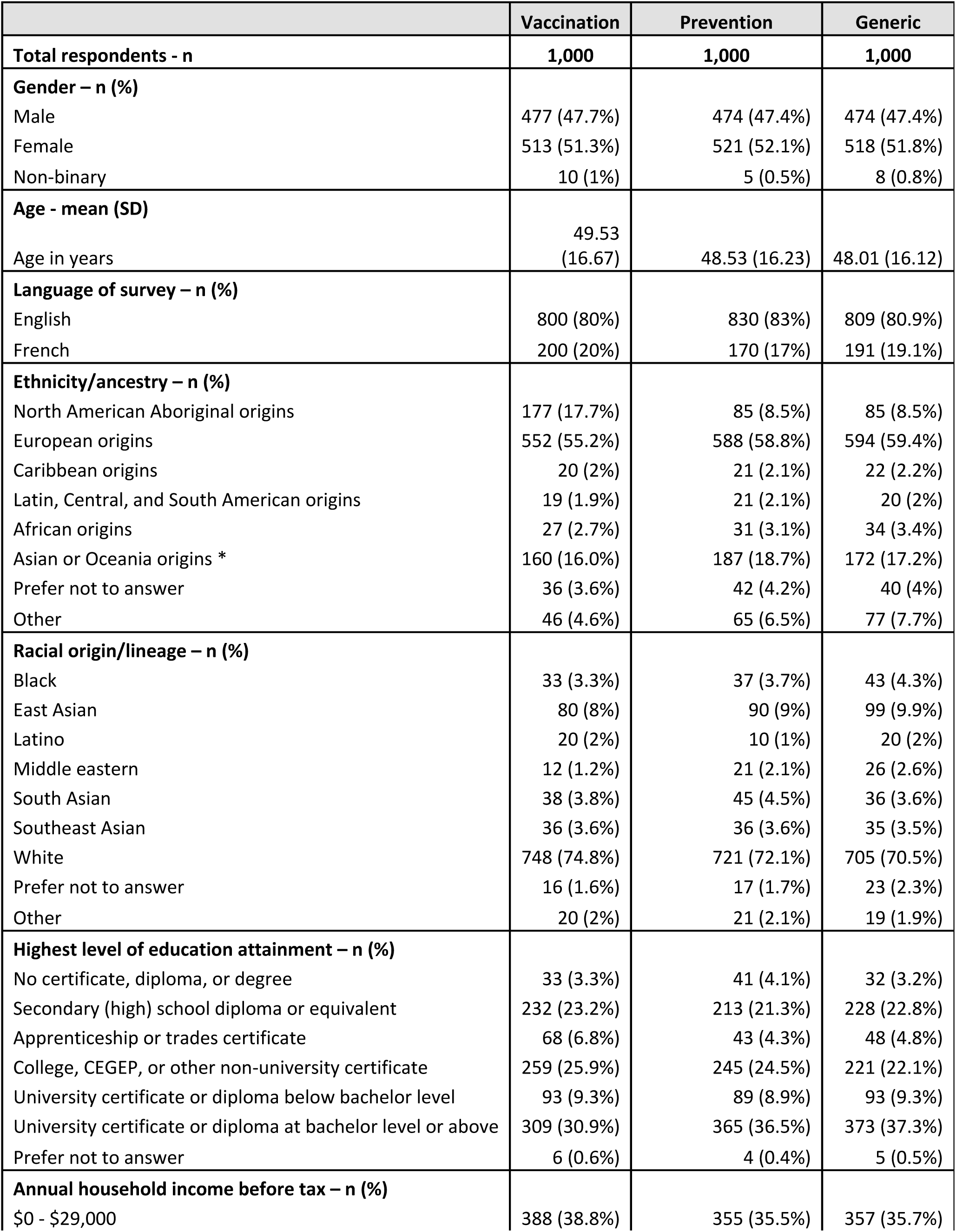
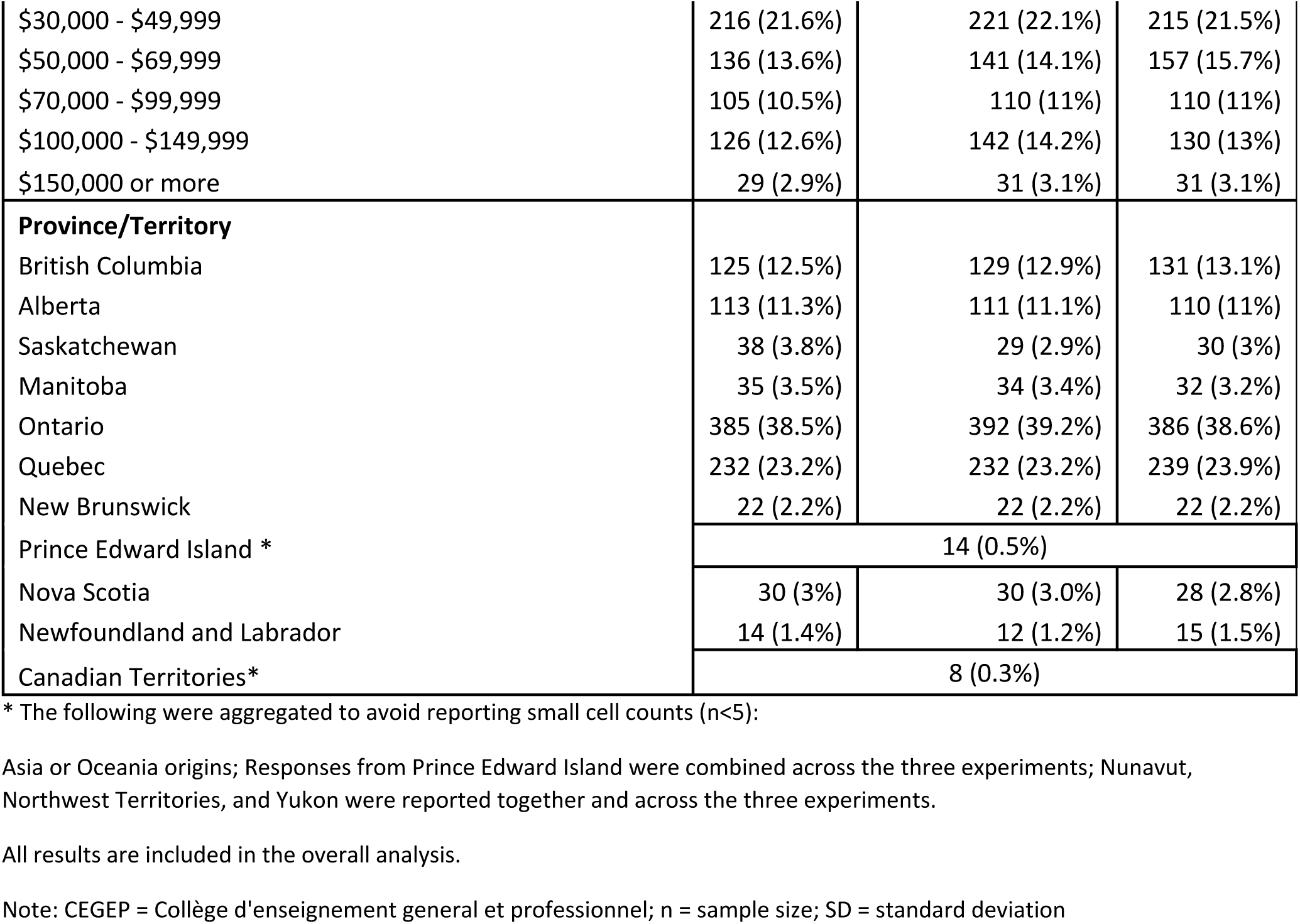
Demographic characteristics.

### 3.2. Response classification

We obtained 543 (54.3%), 477 (47.7%), and 506 (50.6%) logical responses from the vaccination, preventions, and generic experiments, respectively. For the vaccination experiment, approximately one third of the logical responses (31%) were classified as pro-rich (categories 1-3), 1% as health maximizers, 13% as weighted prioritarians (1–7), 2% as maximins, and 54% as egalitarians (1–5). For the *prevention experiment*, 22% responses were categorized as pro-rich (1–3), 1% as health maximizers, 19% as weighted prioritarians (1-7), 3% as maximins, and 55% as egalitarians (1-5). For the *generic experiment*, 16% responses were categorized as pro-rich (1-3), 2% as health maximizers, 22% as weighted prioritarians (1-7), 2% as maximins, and 57% as egalitarians (1-5). The distribution of the responses is summarized in Figure 1. The median respondent was egalitarian across the three experiments.

**Figure 1:**
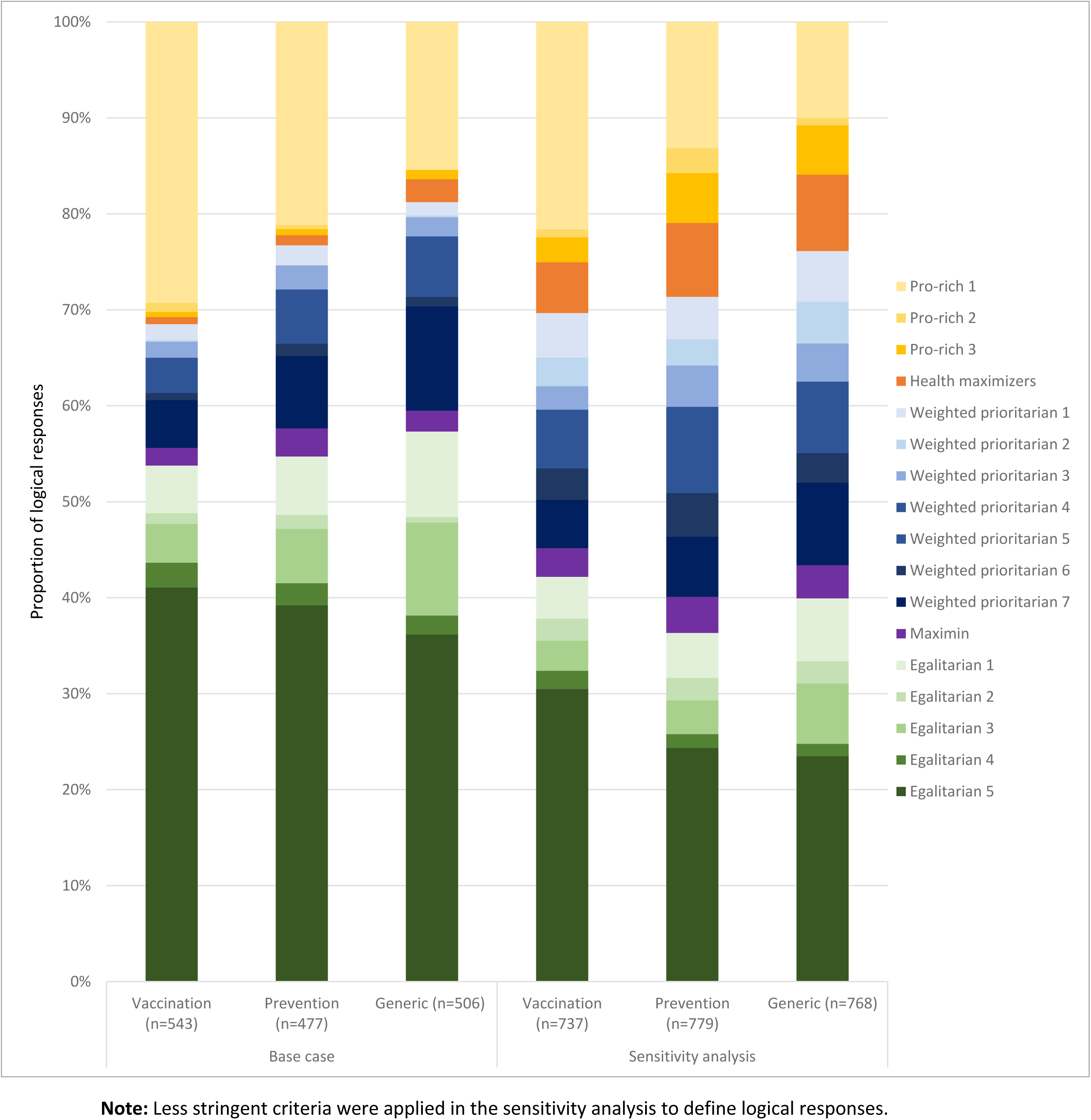
Distribution of logical responses for the base case and the sensitivity analysis among adults living in Canada.

### 3.3. Sensitivity analysis

With less restrictive inclusion criteria, we included a total of 737 (73.7%), 779 (77.9%), and 768 (76.8%) logical responses in the sensitivity analysis for the vaccination, prevention, and generic experiments, respectively. The proportions of responses were modified as follows: pro-rich responses were reduced for all three experiments when compared to the base case, at 25% for vaccination, 20% for prevention, and 15% for generic (i.e., 1 to 5 percentage point reduction). Health maximizers increased to 5%, 6%, and 8% for the vaccination, prevention, and generic experiments, respectively (i.e., 4 to 6 percentage point increase). Weighted prioritarians increased to 24%, 31%, and 32% for the vaccination, prevention, and generic experiments, respectively (i.e., 10 to 12 percentage point increase). Maximins increased to 3%, 4%, and 3%, for the vaccination, prevention, and generic experiments, respectively (i.e., 1 percentage point increase across experiments). Finally, the proportion of egalitarian responses were reduced at 42%, 36%, and 40% for the vaccination, prevention, and generic experiments, respectively (i.e., 11 to 18 percentage point reduction). The median respondents were weighted prioritarians across the three experiments. Figure 1 presents the distribution of logical responses for the sensitivity analysis.

### 3.4. Parameter estimation

Estimated Atkinson indices are presented in Table 3. Higher values indicate a higher level of aversion to health inequality. Aversion indices for responses in the pro-rich categories were between −2.451 and −0.860 (implied weight < 1), representing inequality-seeking preferences. The aversion index for health maximizers was equal to zero (implied weight = 1). Aversion indices ranged from 0.9 to 12.7 among weighted prioritarians (implied weight >1). Finally, the Atkinson Index for maximins reached virtual infinity (implied weight = ∞), while it remained undefined for egalitarians (implied weight undefined). The median respondent’s aversion index corresponded to the egalitarian 1 category for the base case of all three experiments. For the sensitivity analysis, the median respondent’s aversion index was categorized as weighted prioritarian 7 (Atkinson index = 12.7) for the vaccination and generic experiments, and weighted prioritarian 6 (Atkinson Index = 8.6) for the prevention experiment. We summarized the implied weight from the isoelastic transformation of the EDE in Figure 2. At the baseline average life expectancy of 68 years for individuals in the lowest household income quintile, and an Atkinson index of 12.7, respondents weighted each additional healthy life year gained by populations in the lowest household income quintiles 6.7 times higher relative to individuals in the highest household income quintile.

**Figure 2:**
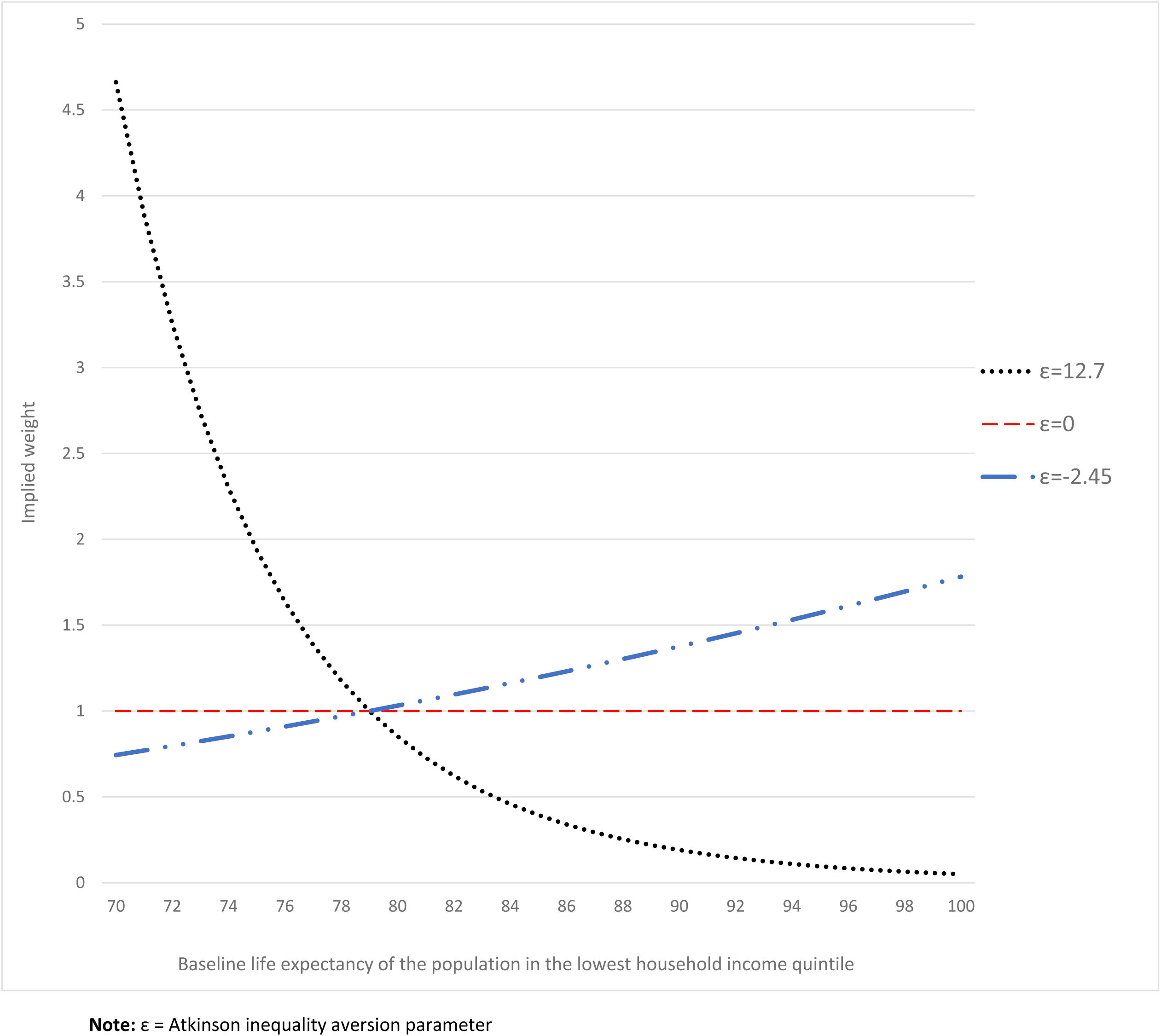
Marginal social value of a change in health, relative to the population in the highest household income quintile (i.e., life expectancy of 79 years), given ε=-2.45, ε=0, and ε=12.7

**Table 3:**
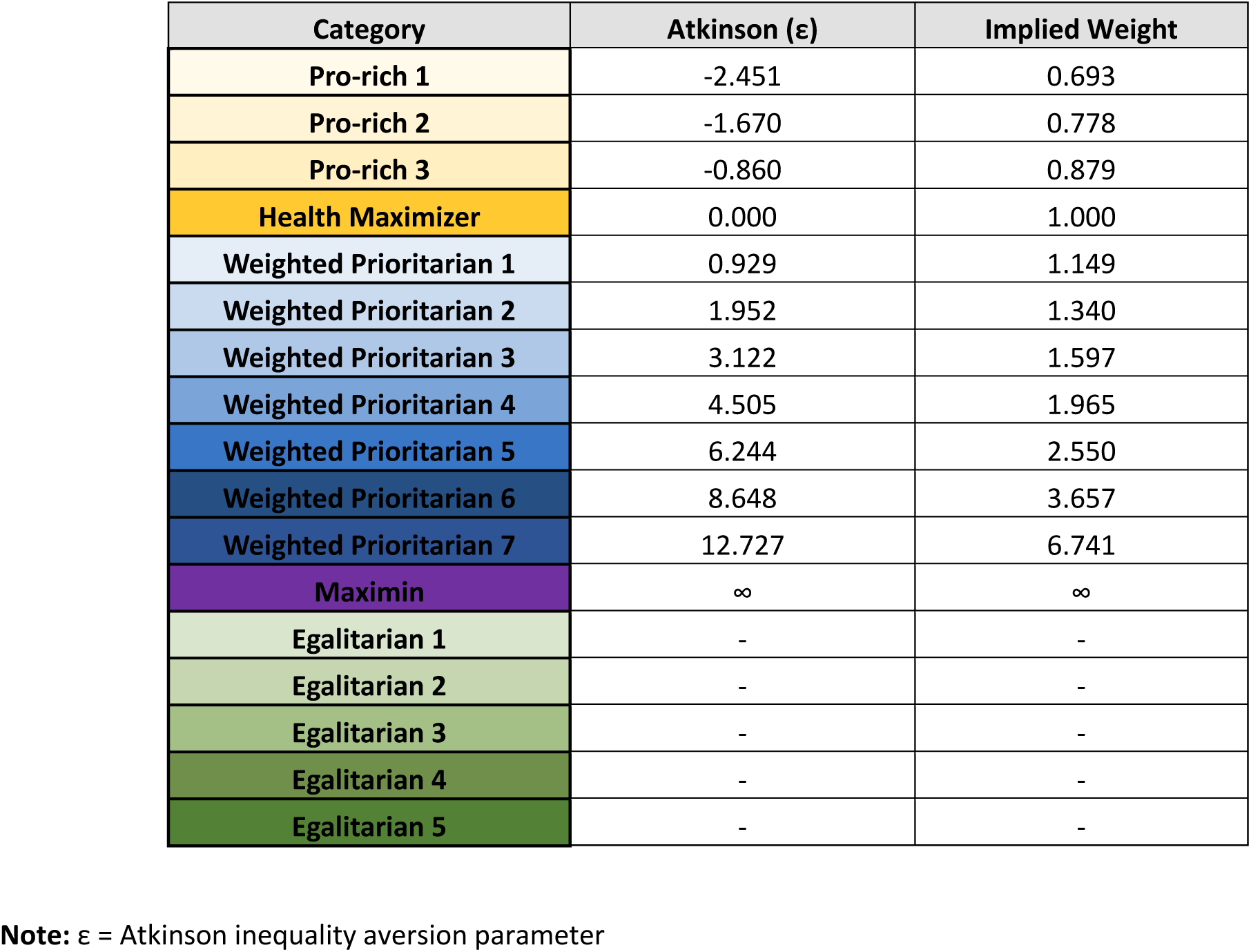
Parameter elicitation for each logical response classification.

### 3.5. Stratified results

We stratified the preference distributions by sociodemographic characteristics. The two main variables that showed a consistent effect throughout the three experiments were gender and income level. Women’s responses were more likely to be classified as egalitarian, while men’s responses were more often categorized as pro rich. The median woman’s response was classified between egalitarian 3 and egalitarian 4, and the median man’s response was between weighted prioritarian 5 and maximin, across the three experiments. For income level, the proportion of egalitarian responses was inversely related to income. Close to 60% of responses for respondents reporting a household income between $0 and $29,000 were classified as egalitarians. This proportion was reduced to approximately 50% among those with household income levels greater than or equal to $100,000. The stratified results for gender and income are summarized in Figure 3. Other variables, including age, highest education attainment, region, racial lineage, and ethnic origin did not affect the distribution of preferences consistently across the three experiments. All stratified results are reported in Appendix 6.

**Figure 3:**
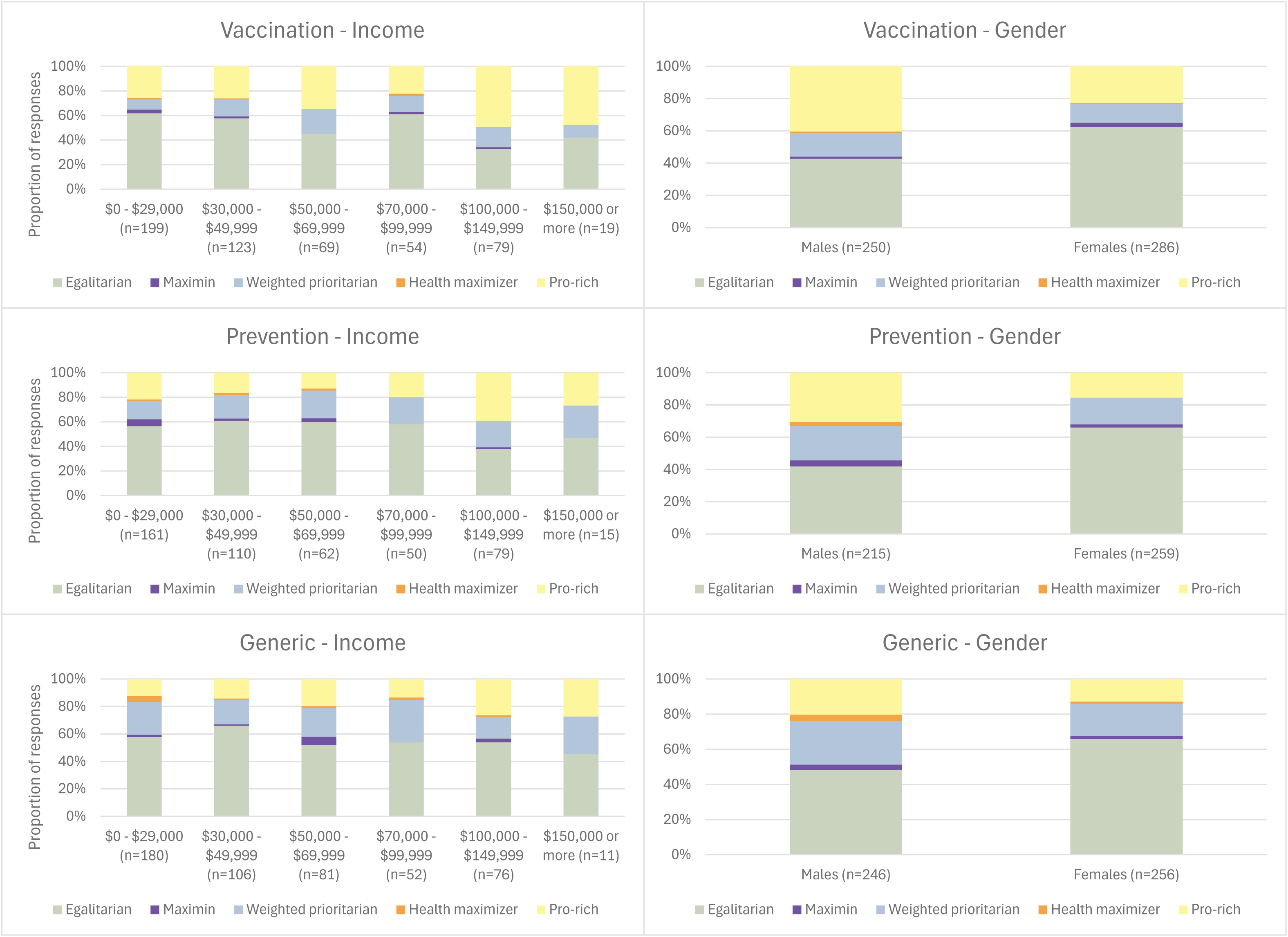
Stratified preferences across gender and income levels

## 4. Discussion

Our benefit trade-off analysis revealed considerable health inequality aversion among the Canadian population, with over half of the respondents willing to trade off efficiency for more equitable health outcomes. Furthermore, our results suggest that intervention type impacts the level of health inequality aversion. For vaccination programs, most responses were located at the extremes of the distribution (i.e., pro-rich or egalitarians). In contrast, for generic interventions the proportion of responses with inequality-seeking preferences was lower. Women had stronger preferences for reducing health inequalities than men. Additionally, respondents reporting an annual household income greater than or equal to $100,000 were less averse to health inequalities when compared to respondents with a reported annual household income under $50,000. Canadian evidence on population preferences for inequality aversion is limited to one study (13).

Hurley et al evaluated inequality aversion for income (ɛ=3.268) and health (ɛ =1.174) (13). The level of health inequality aversion was not statistically different from zero. The experiment included a single trade-off scenario, for which respondents had to choose between two hypothetical policies. Choice experiments with only one trade-off scenario can fail to capture preferences that might only be observed at the extremes of the health benefit distribution. Consequently, our study fills this important knowledge gap by providing robust inequality aversion indices that were estimated from a multi-choice trade-off experiment, which can directly inform EIEEs (29).

Our findings align with inequality aversion patterns observed in other countries. A recent scoping review found that ∼75% of studies that employed social welfare functions to assess equity-efficiency trade-offs reported evidence of inequality aversion (8). The closest example to our study was Robson et al., which employed a comparable experiment design and estimated a median Atkinson Index of 10.95 in the UK (27). Although the evidence suggests a high level of aversion to health inequality worldwide, studies have reported a considerable proportion of extreme responses (14). Further, previous research had indicated that benefit trade-off analyses with concrete programs would likely result in a reduction of extreme responses (14). However, we found a substantially higher proportion of inequality-seeking preferences and a more extreme bimodal distribution of preferences for the concrete program, i.e., vaccination. Our finding could potentially, in part, be explained by polarization with respect to vaccination in the aftermath of the COVID-19 pandemic (30), indicating that the specific type programs used in benefit trade-off analysis can impact the proportion of extreme responses in either direction.

Our study has some limitations. First, we compared income-related health inequalities only in the equity-efficiency trade-offs. However, decision-makers consider several equity dimensions in their deliberations and decisions, e.g., age, gender, ethnicity, geographic location, social capital, and health status (31). Therefore, future work should evaluate whether value judgements differ by equity dimensions considered in the trade-off experiment. Engaging in discussions with key Canadian decision-makers (e.g., health system decision-makers, patients, HTA agency representatives) could help identify equity domains relevant to real-world decisions. Second, some demographic groups had low numbers of respondents. While representative, we recruited a relatively low number of respondents from smaller Provinces and the Territories due to the quotas. Larger studies could ensure a greater number of respondents across demographic characteristics while maintaining representativeness. Our analysis, with a sample size of 3,000, is the largest among recently published equity-efficiency trade-off studies - where less than 15% of the studies recruited over 1,000 respondents (8), allowing us to explore the impact of program type and further stratification.

Our findings have important implications for HTA and policymaking. Leading HTA agencies recognized the importance of EIEEs to inform resource allocation and priority setting. However, a major limitation to the adoption of EIEEs in Canada is the lack of Canadian preferences for inequality aversion. Our study is fundamental in adopting EIEEs to inform policy decision-making.

## 5. Conclusion

People living in Canada showed considerable aversion to health inequalities between populations in the highest and lowest household income quintiles. More than half of the respondents were willing to trade off a considerable amount of health to achieve a more equitable distribution. These results were consistent across three different experiments (i.e., vaccination, prevention, and generic). Abstract or generic experiments resulted in less extreme distributions. Women and respondents from lower-income households had greater aversion to health inequality. This evidence enables the integration of equity into economic evaluations to inform resource allocation decisions in Canada.

## Data Availability

All data produced in the present study are available upon reasonable request to the authors

## Supplementary material

## Appendix 1: Quotas established by AskingCanadians based on census data

### Individual quotas for gender, age, and province

|  | Census % | Quota (N/1,000) |
| --- | --- | --- |
| **Gender** |  |  |
| Male | 48.00% | 480 |
| Female | 52.00% | 520 |
| **Age (years)** |  |  |
| 18-34 | 27.90% | 279 |
| 35-44 | 16.90% | 169 |
| 45-54 | 20.10% | 201 |
| 55-64 | 16.50% | 165 |
| 65+ | 18.60% | 186 |
| **Province** |  |  |
| Ontario | 38.40% | 384 |
| British Columbia | 13.10% | 131 |
| Alberta | 11.00% | 110 |
| Manitoba | 3.60% | 36 |
| Saskatchewan | 3.10% | 31 |
| Quebec **<u>French</u>** | 23.90% | 239 |
| Nova Scotia | 2.80% | 28 |
| New Brunswick | 2.10% | 22 |
| Newfoundland | 1.40% | 15 |
| Prince Edward Island | 0.30% | 4 |
| Northwest Territories | 0.10% | 1 |
| Nunavut | 0.10% | 1 |
| Yukon Territory | 0.10% | 1 |

### Interlocked quotas for income and region

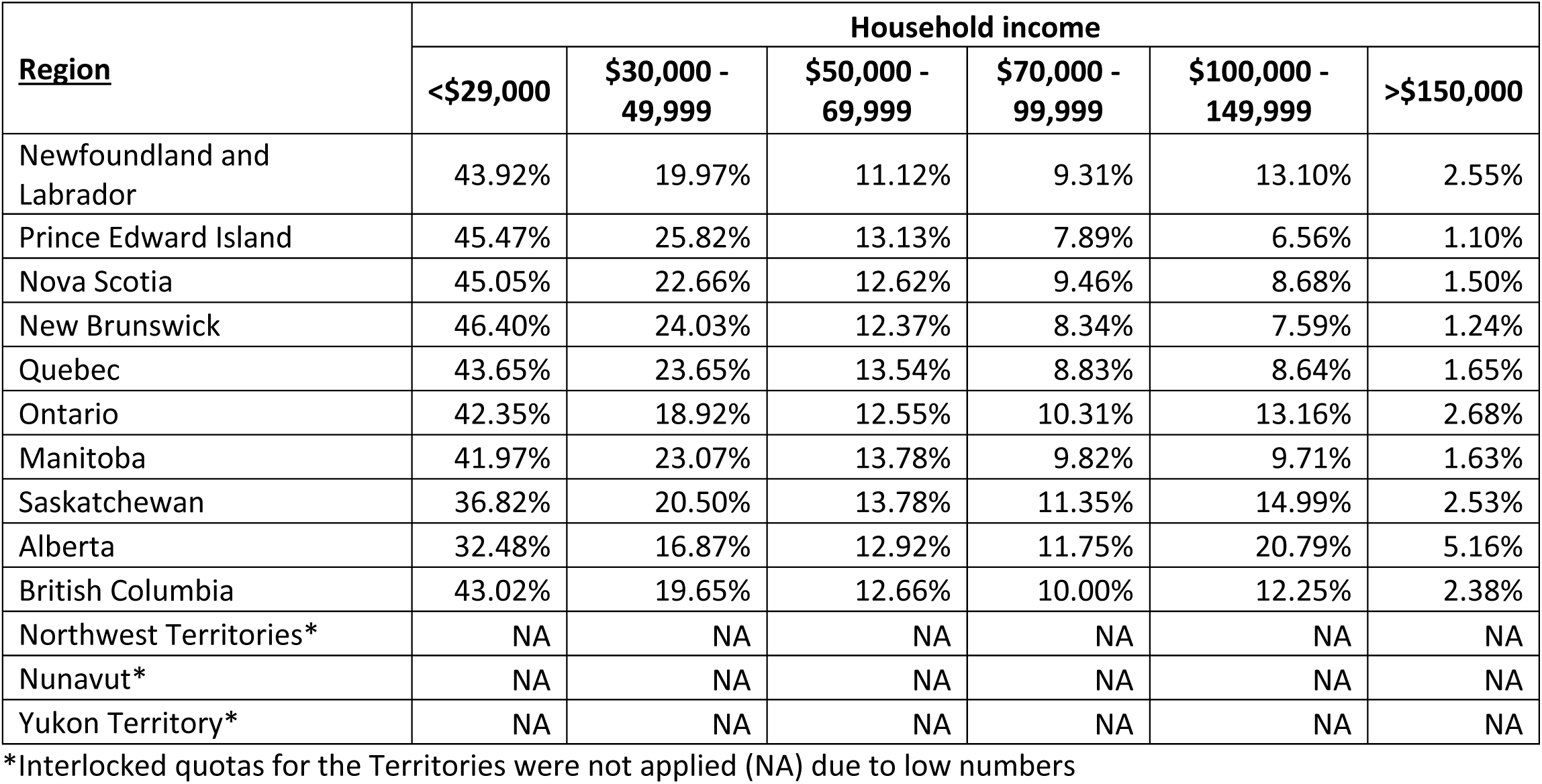

### Expected recruitment across income and region

| **Income** | **<\$29,000** | **\$30,000 - 49,999** | **\$50,000 - 69,999** | **\$70,000 - 99,999** | **\$100,000 - 149,999** | **>\$150,000** |
| --- | --- | --- | --- | --- | --- | --- |
| Newfoundland and Labrador | 7 | 3 | 2 | 1 | 2 | 0 |
| Prince Edward Island | 2 | 1 | 1 | 0 | 0 | 0 |
| Nova Scotia | 13 | 6 | 4 | 3 | 2 | 0 |
| New Brunswick | 10 | 5 | 3 | 2 | 2 | 0 |
| Quebec Eng | 21 | 11 | 6 | 4 | 4 | 1 |
| Quebec Fr | 83 | 45 | 26 | 17 | 17 | 3 |
| Ontario | 163 | 73 | 48 | 40 | 51 | 10 |
| Manitoba | 15 | 8 | 5 | 4 | 3 | 1 |
| Saskatchewan | 11 | 6 | 4 | 4 | 5 | 1 |
| Alberta | 36 | 19 | 14 | 13 | 23 | 6 |
| British Columbia | 56 | 26 | 17 | 13 | 16 | 3 |
| Northwest Territories* | NA | NA | NA | NA | NA | NA |
| Nunavut* | NA | NA | NA | NA | NA | NA |
| Yukon Territory* | NA | NA | NA | NA | NA | NA |

\*Interlocked quotas for the Territories were not applied (NA) due to low numbers

### Expected recruitment across gender and region

| **Gender** | **Male** | **Female** |
| --- | --- | --- |
| Newfoundland and Labrador | 7 | 8 |
| Prince Edward Island | 2 | 2 |
| Nova Scotia | 13 | 15 |
| New Brunswick | 11 | 11 |
| Quebec Eng | 23 | 25 |
| Quebec Fr | 92 | 99 |
| Ontario | 184 | 200 |
| Manitoba | 17 | 19 |
| Saskatchewan | 15 | 16 |
| Alberta | 53 | 57 |
| British Columbia | 63 | 68 |
| Northwest Territories* | NR | NR |
| Nunavut* | NR | NR |
| Yukon Territory* | NR | NR |

\*Interlocked quotas for the Territories were not applied due to low numbers

### Expected recruitment across age group and region

| **Age** | **18-34** | **35-44** | **45-54** | **55-64** | **65+** |
| --- | --- | --- | --- | --- | --- |
| Newfoundland and Labrador | 4 | 3 | 3 | 2 | 3 |
| Prince Edward Island | 1 | 1 | 1 | 1 | 1 |
| Nova Scotia | 8 | 5 | 6 | 5 | 5 |
| New Brunswick | 6 | 4 | 4 | 4 | 4 |
| Quebec Eng | 13 | 8 | 10 | 8 | 9 |
| Quebec Fr | 53 | 32 | 38 | 32 | 36 |
| Ontario | 107 | 65 | 77 | 63 | 71 |
| Manitoba | 10 | 6 | 7 | 6 | 7 |
| Saskatchewan | 9 | 5 | 6 | 5 | 6 |
| Alberta | 31 | 19 | 22 | 18 | 20 |
| British Columbia | 37 | 22 | 26 | 22 | 24 |
| Northwest Territories* | NR | NR | NR | NR | NR |
| Nunavut* | NR | NR | NR | NR | NR |
| Yukon Territory* | NR | NR | NR | NR | NR |

\*Interlocked quotas for the Territories were not applied due to low numbers

## Appendix 1: Health gains across eight scenarios

| Scenarios | Health gains |  |  |  |
| --- | --- | --- | --- | --- |
|  | Universal vaccination / Program A /<br>Universal prevention |  | Tailored vaccination / Program B /<br>Tailored prevention |  |
|  | Highest<br>household<br>income quintile | Lowest<br>household<br>income quintile | Highest<br>household<br>income quintile | Lowest<br>household<br>income quintile |
| 1 | 7 | 3 | 3 | 8 |
| 2 | 7 | 3 | 3 | 7 |
| 3 | 7 | 3 | 3 | 6 |
| 4 | 7 | 3 | 3 | 5 |
| 5 | 7 | 3 | 3 | 4 |
| 6 | 7 | 3 | 3 | 3 |
| 7 | 7 | 3 | 3 | 2 |
| 8 | 7 | 3 | 3 | 1 |

Note: the health gains were included as per Ali et al. (14)

## Appendix 3: Vaccination Survey

### Understanding public views on funding the health system

Welcome to our online survey. This survey was designed to help understand views around health inequalities in Canada.

This questionnaire should take about 10-15 minutes to complete.

Your responses will be kept strictly anonymous and used only for research purposes.

Please click on the NEXT button below to start the survey. You can navigate backwards and forwards within the survey and change your original responses at any time before pressing the final ‘SUBMIT’ button at the end.

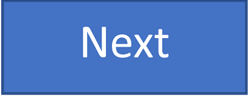

We will ask four questions to assess your eligibility for the survey.

Your responses will be kept strictly anonymous and used only for research purposes.

1. Your age?

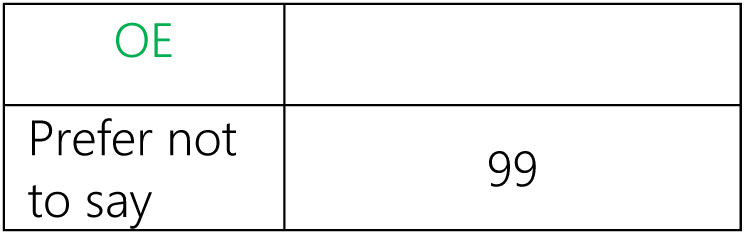
2. Your gender?

|  |  |
| --- | --- |
| OE |  |
| Prefer not to say | 99 |

3. In which province/territory do you live?

**2. Your gender?**
|  |  |
| --- | --- |
| Male | 1 |
| Female | 2 |
| Non-Binary | 3 |
| Prefer not to say | 99 |

4. What are the first three characters of your postal code? (Format A1A)

**3. In which province/territory do you live?**
|  |  |
| --- | --- |
| Alberta | 1 |
| British Columbia | 2 |
| Manitoba | 3 |
| Newfoundland and Labrador | 4 |
| New Brunswick | 5 |
| Northwest Territories | 6 |
| Nova Scotia | 7 |
| Nunavut | 8 |
| Ontario | 9 |
| Prince Edward Island | 10 |
| Quebec | 11 |
| Saskatchewan | 12 |
| Yukon Territory | 13 |
| I live outside of Canada | 14 |
| Prefer not to say | 99 |

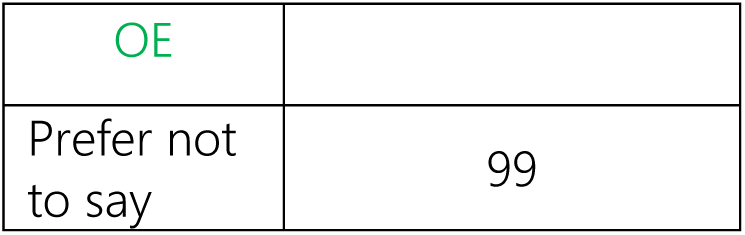 In the next few screens, you will be provided with an introduction to this survey, with some information for you to review. You will then be asked some questions based on this information. **Introduction** There are differences between populations with the **highest and lowest household incomes** in Canada in terms of how long people live, their quality of life, and their access to healthcare. If we divide the population into five groups of equal size (each group = 20% of the population), then in 2019:
  - The highest-earning 20% of the population had an average household income after tax of **$160,000** per year.
  - The lowest earning 20% of the population had an average household income after tax of **$25,000** per year.

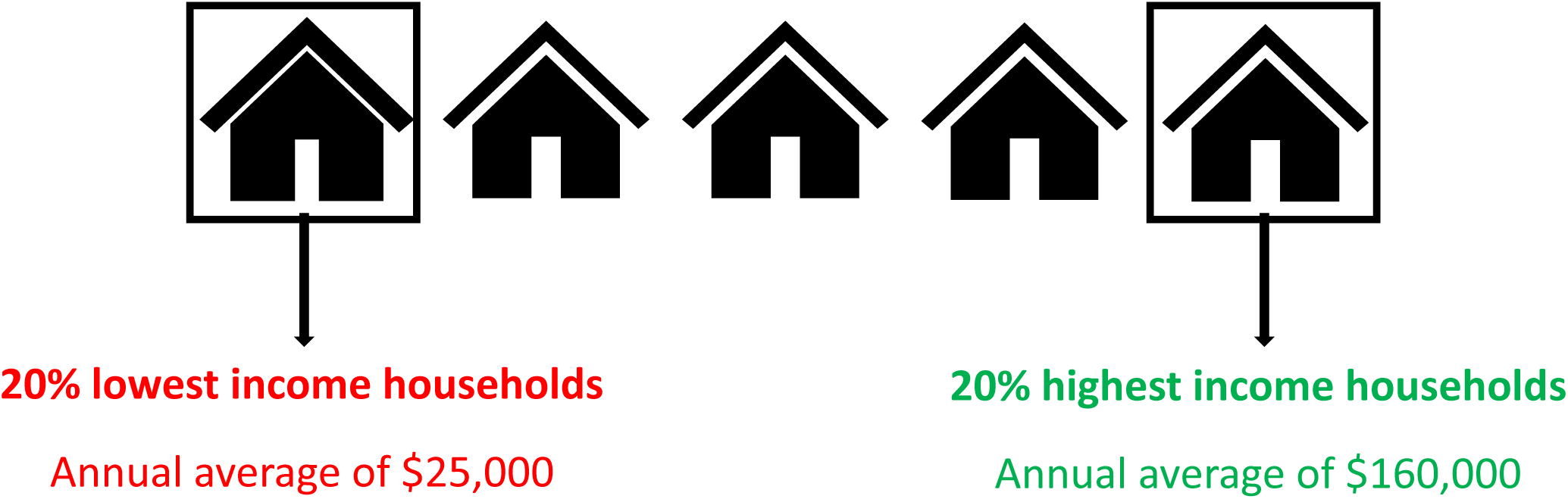
  - While actual length of life and health vary between individuals, on average, the **20% highest household income population** experience **79 years** of life in full health.
  - On the other hand, the **20% lowest household income population** experience **68 years** of life in full health. For example, someone who has 79 years in full health might have lived to 85 years old, but with less than full health. These are averages across the opulation of Canada. Each individual’s actual length of life and health can vary considerably from these averages. Introduction
  - Imagine that you are asked to choose between **two vaccination** programs to prevent a **respiratory infectious disease**. Respiratory diseases are caused by organisms such as viruses or bacteria that affect mainly the respiratory system (e.g., lungs and/or throat). The organisms can be spread by coughing, sneezing or face-to-face contact.
  - The first choice is a **tailored vaccination program** that is designed to prioritize vaccine uptake among **low-income households**, because of two reasons: (1) low-income households might be more at risk of getting infected; (2) low-income households may face barriers to access vaccines. So, the objective of this tailored program is to understand why vaccination among low-income households is relatively lower, and then modify the program to help increase the number of people in this population group who get vaccinated.
  - The second choice is a **universal vaccination program**, where the vaccines are offered to everyone in Canada, but no specific efforts are made to increase vaccination rates among individuals from **low-income households.** This means low-income households will always be less likely to access vaccines.
  - Both programs improve the life expectancy of the two populations, but to different levels. Assume that both programs **cost the same** to implement.
  - The programs offer the opportunity for vaccination. However, vaccination is not mandatory for anyone.

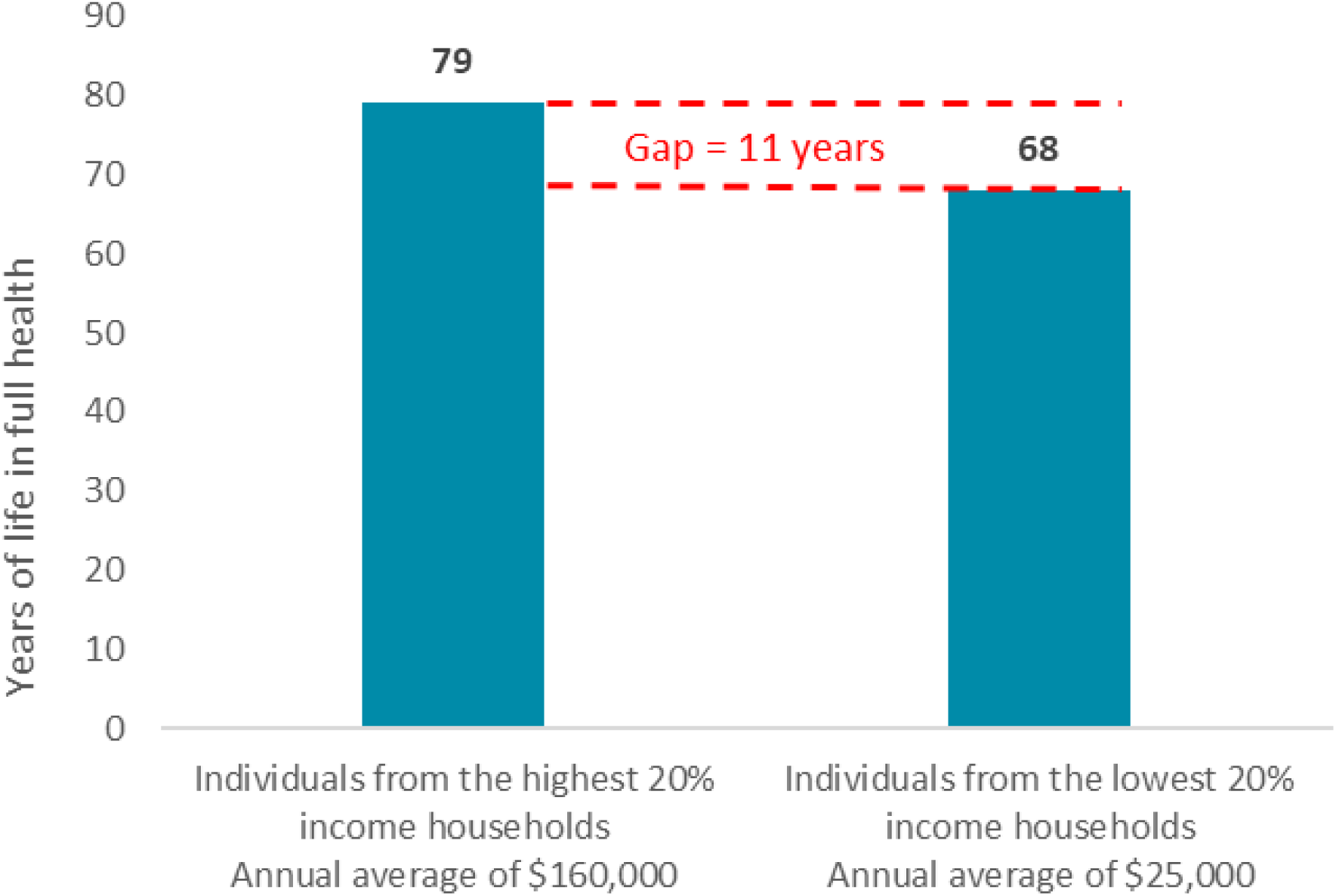

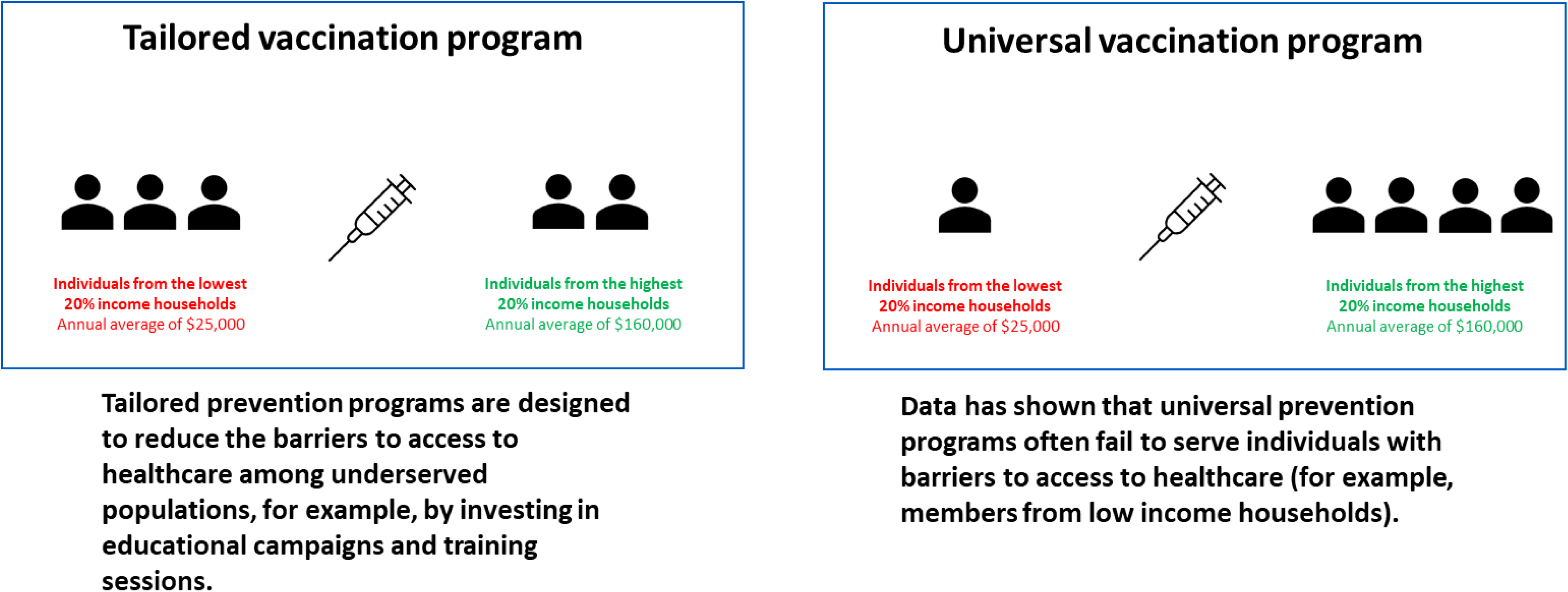

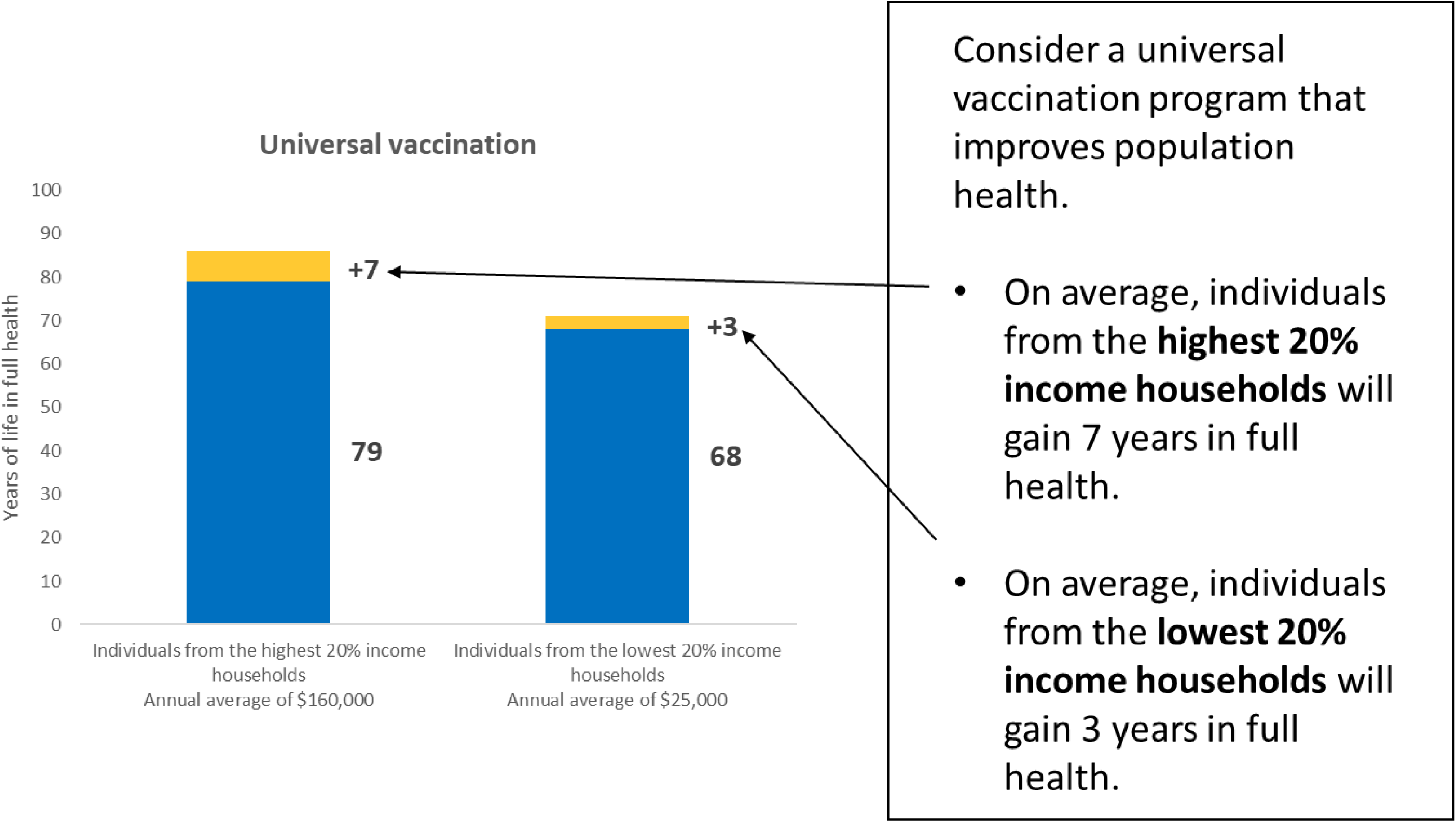

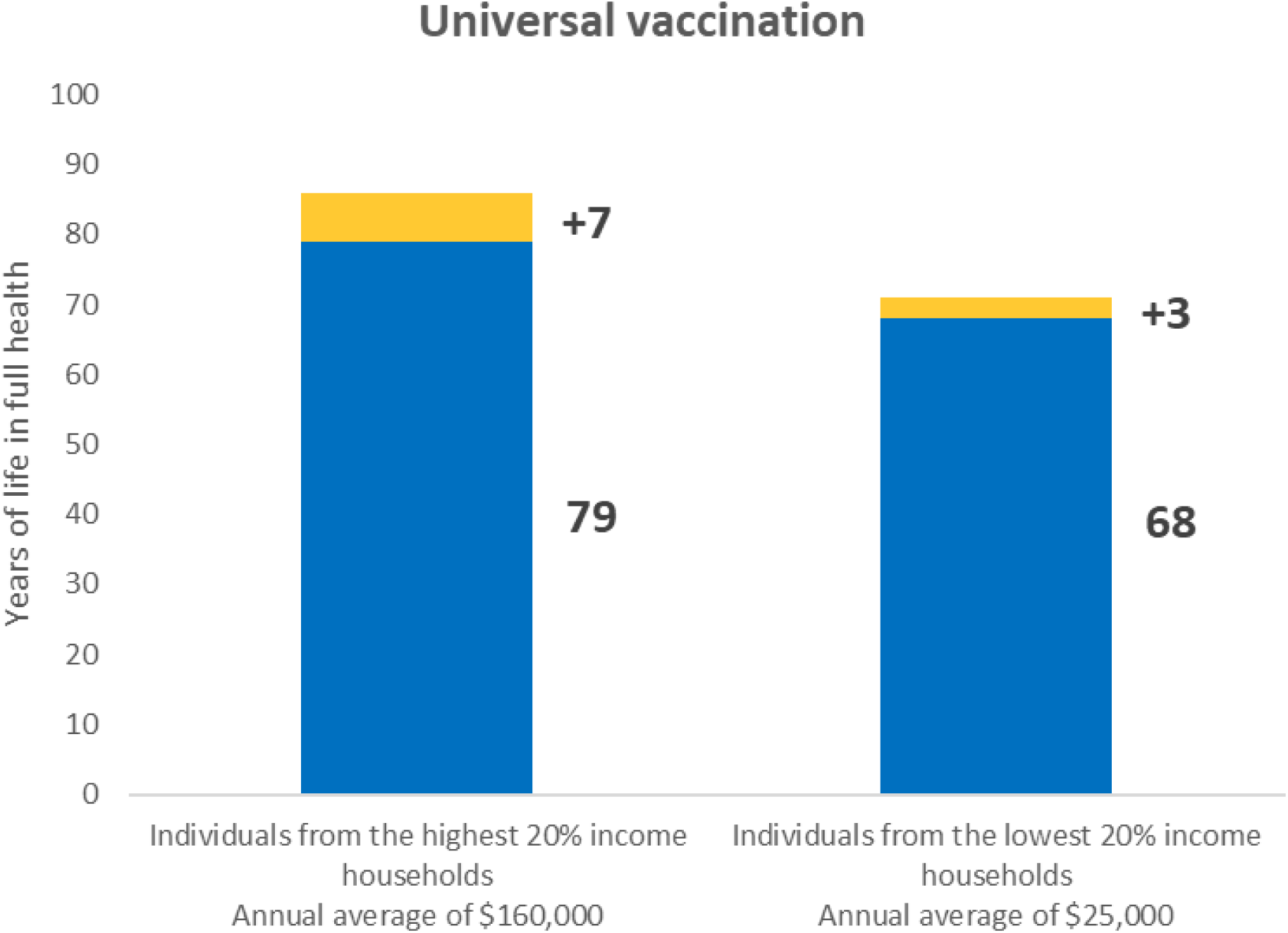
5. Which population group has the highest life expectancy AFTER implementing the vaccination program?

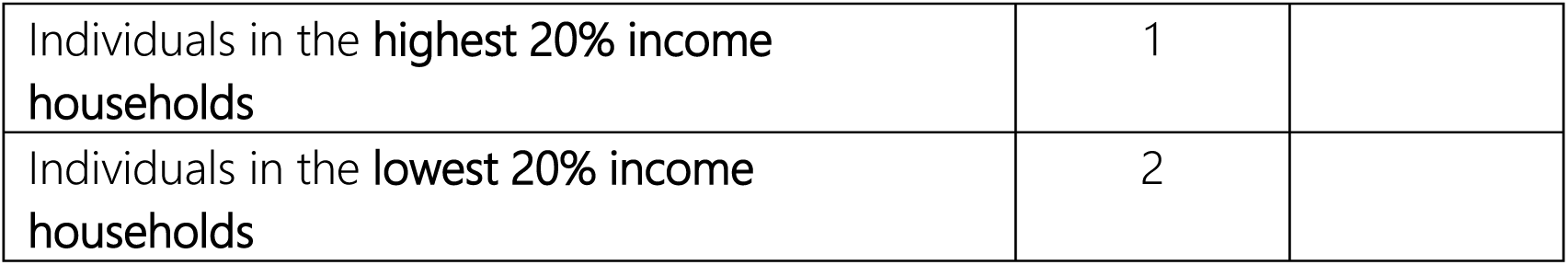

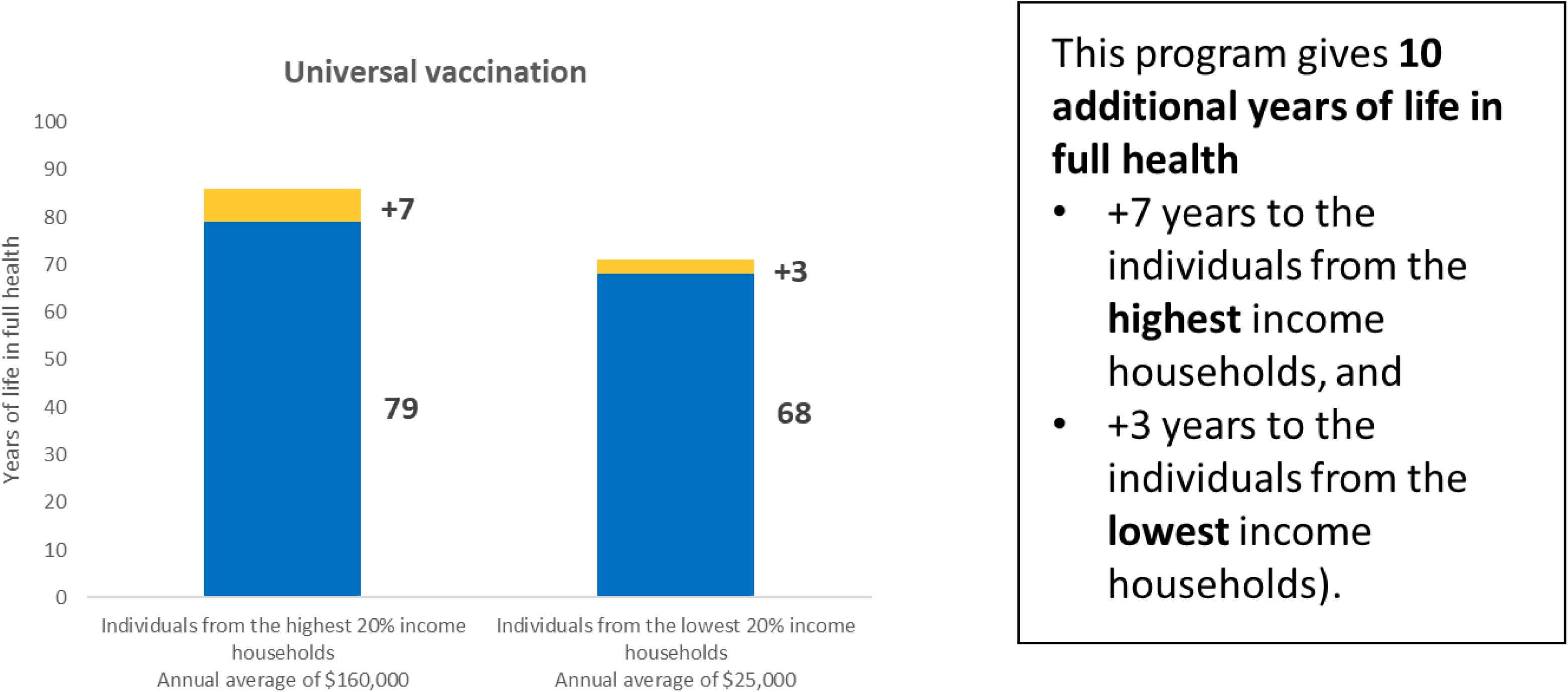

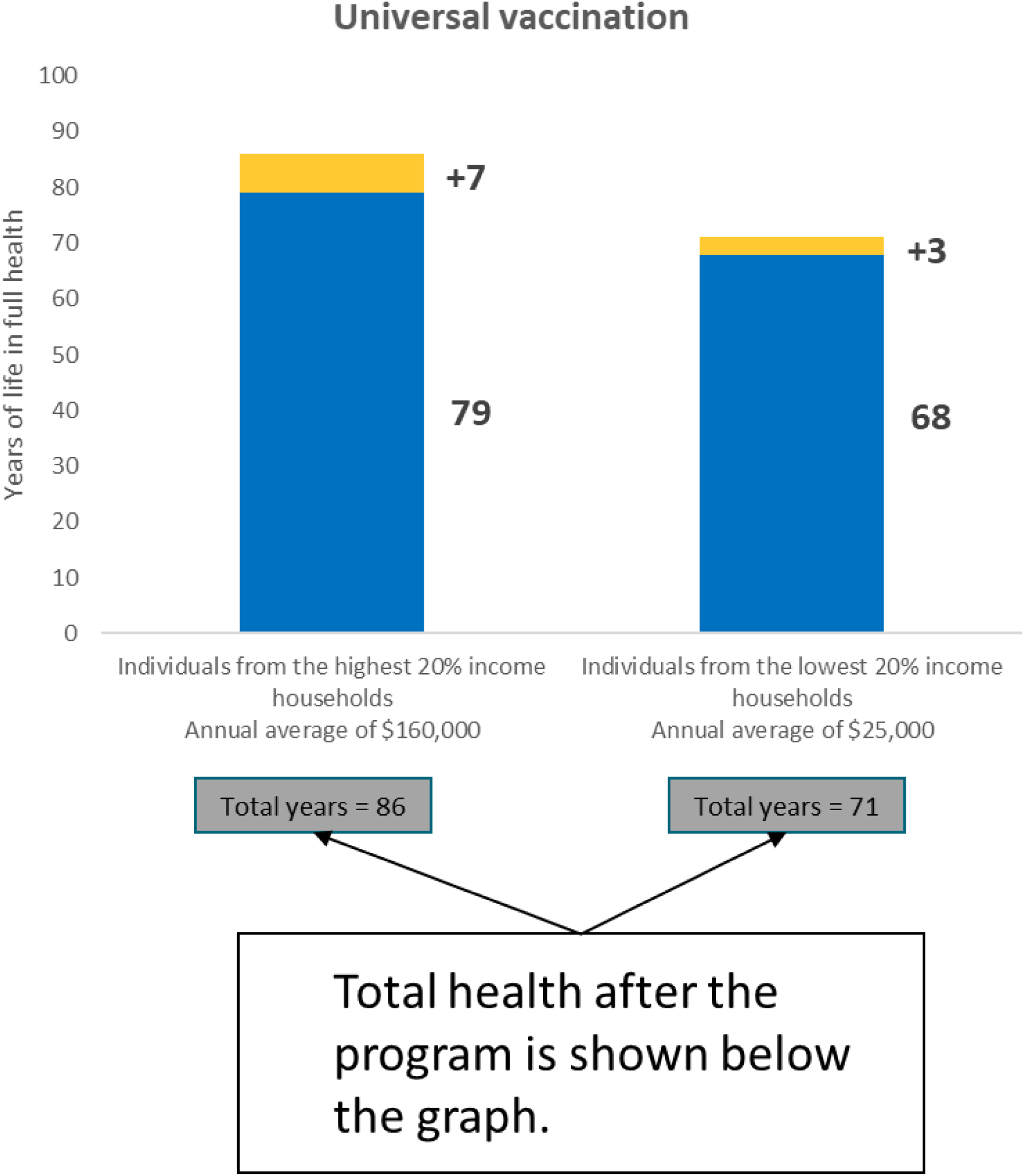

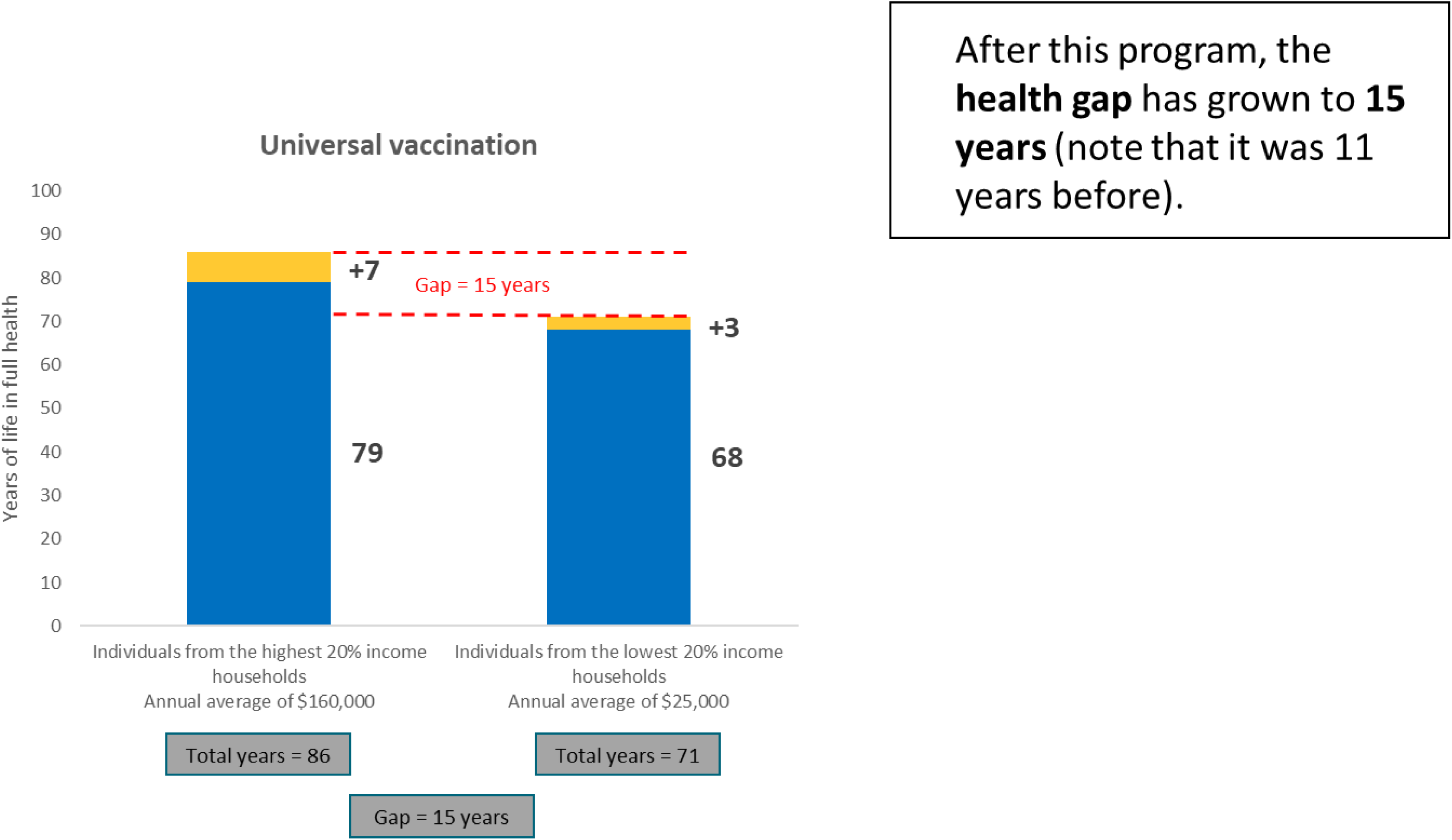

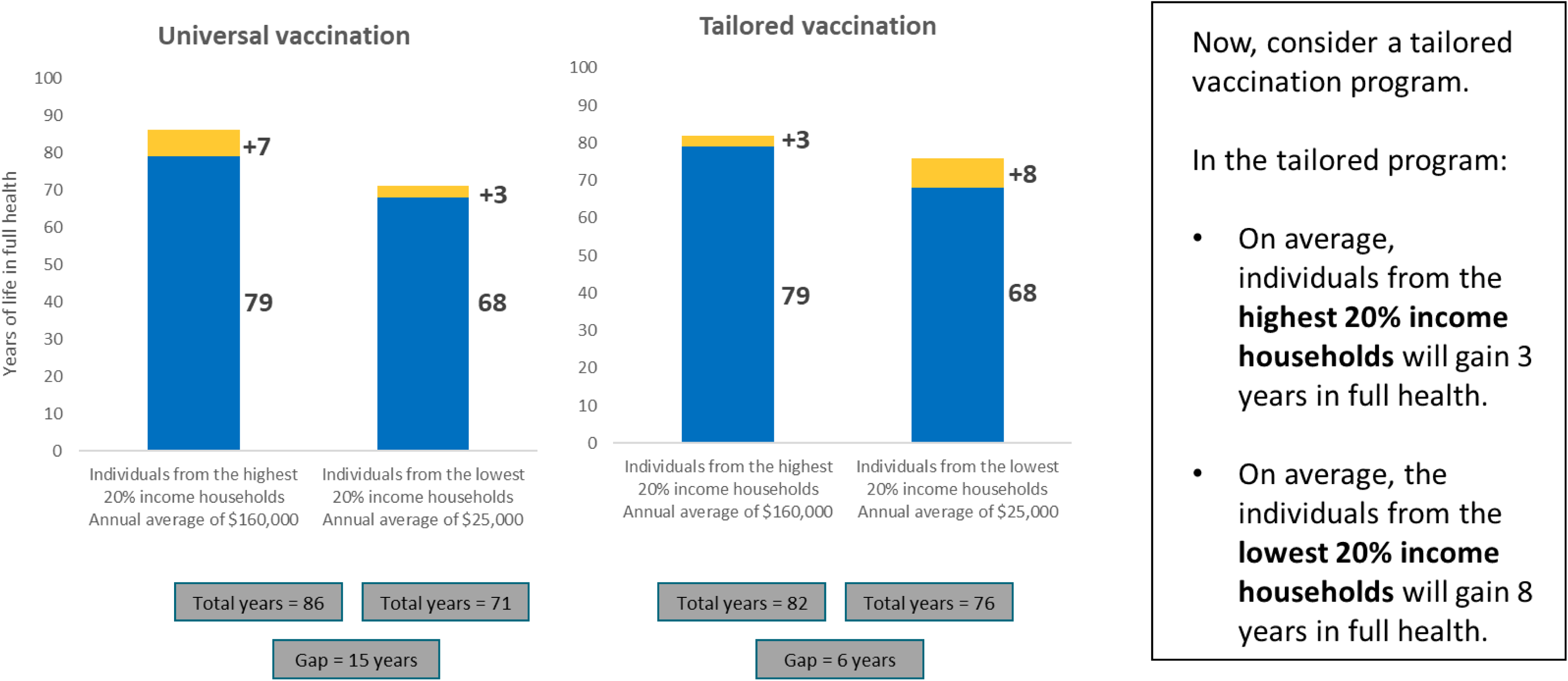

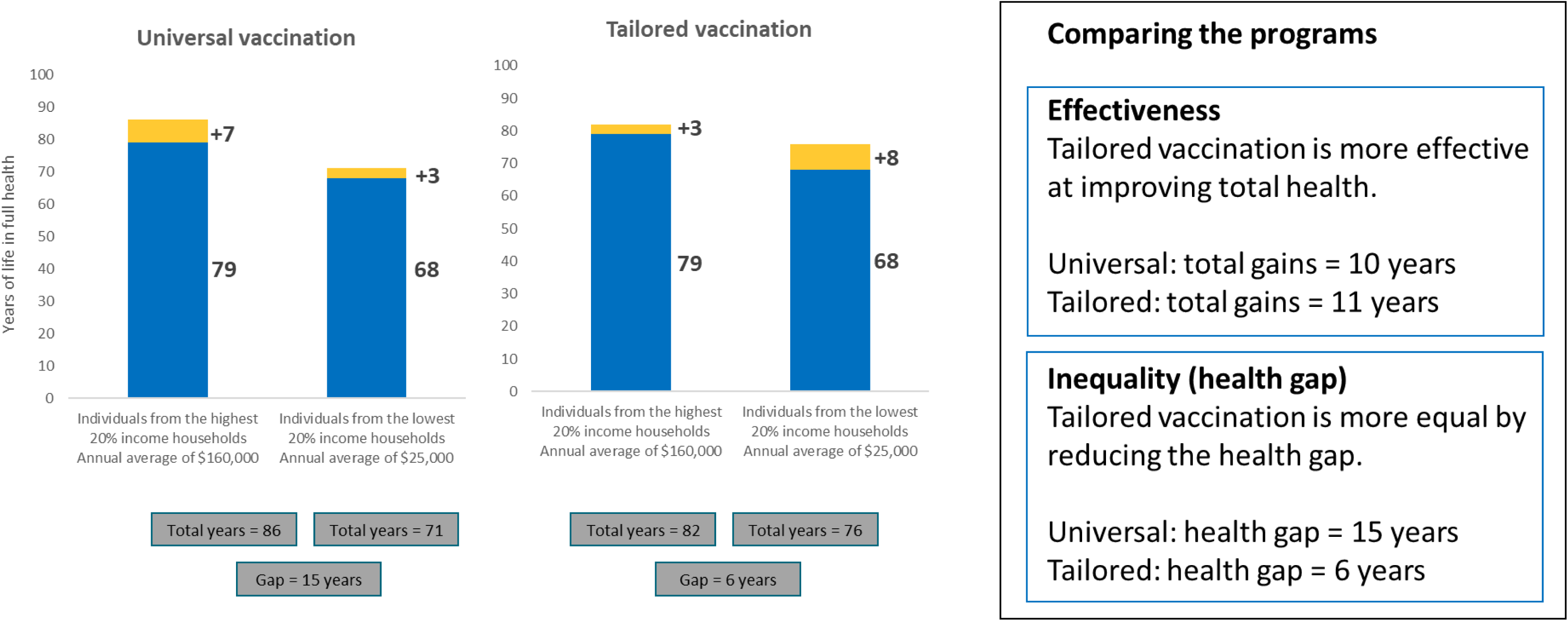

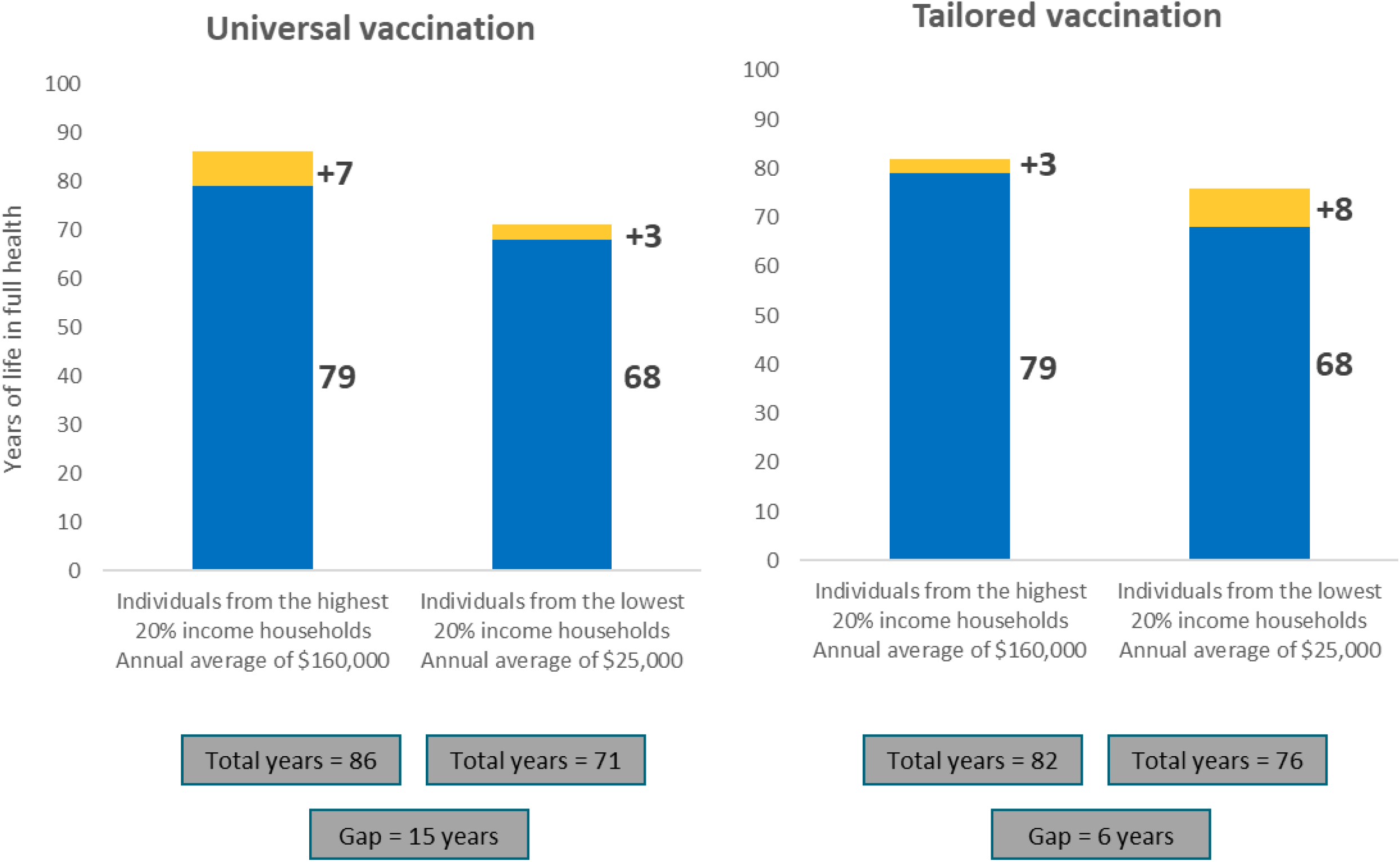
6. Which program would you choose?

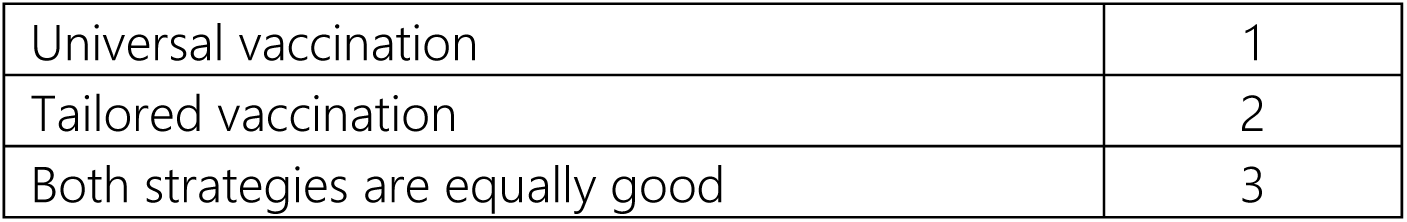 Survey questions Which program should the government choose? Answer the next 8 scenarios considering:
  - The programs offer the opportunity for vaccination. However, vaccination is not mandatory for anyone
  - We cannot pay for both programs — a choice must be made.
  - “Either program is good” means you don’t mind which one is chosen.
  - Both programs cost exactly the same.
  - The only difference between the programs is how much health is gained by the individuals in the highest and lowest 20% income households.
  - The population in the middle is not affected.

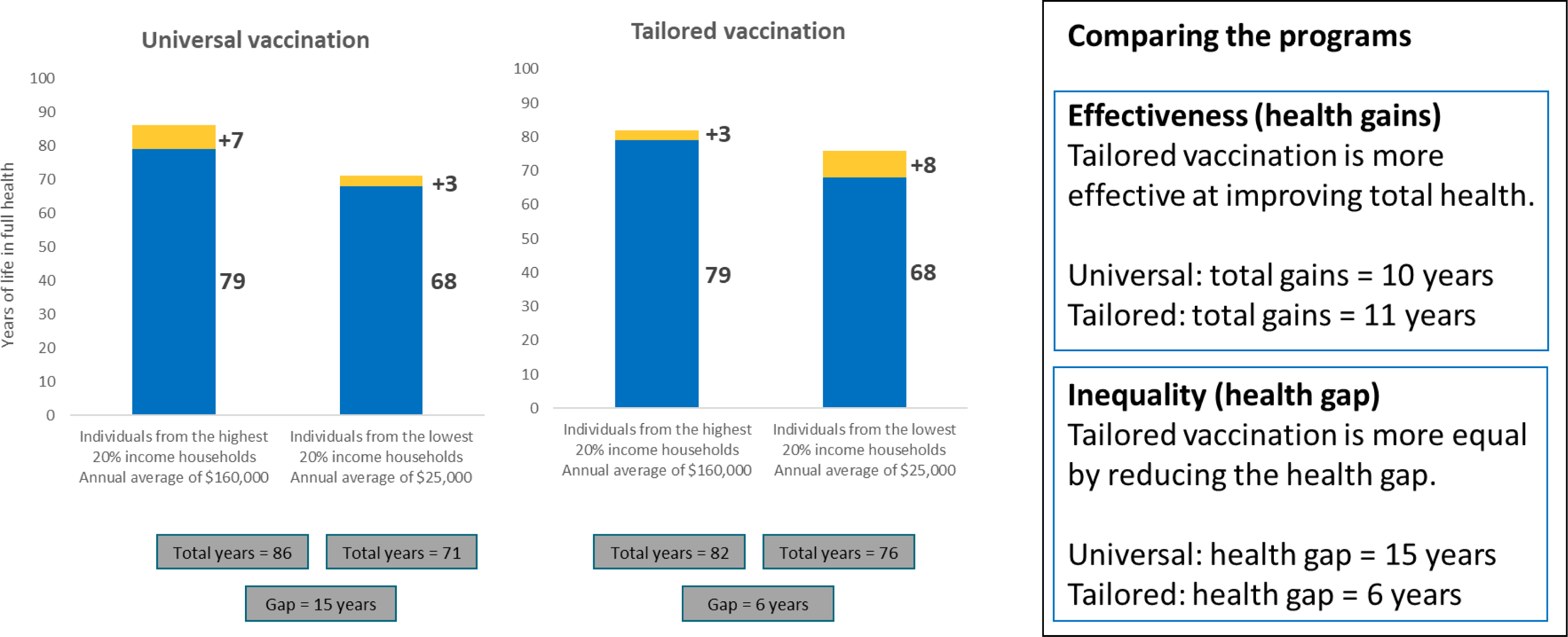
7. Which program would you choose?

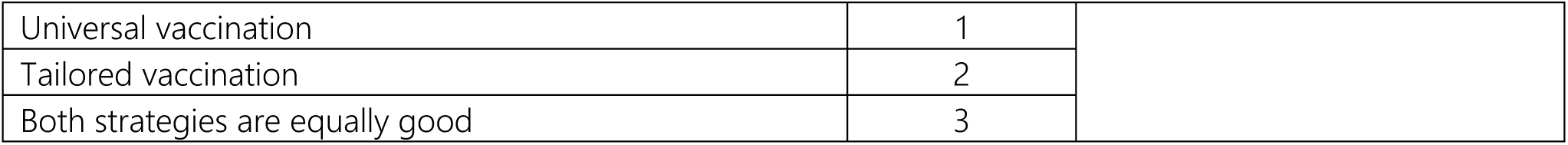

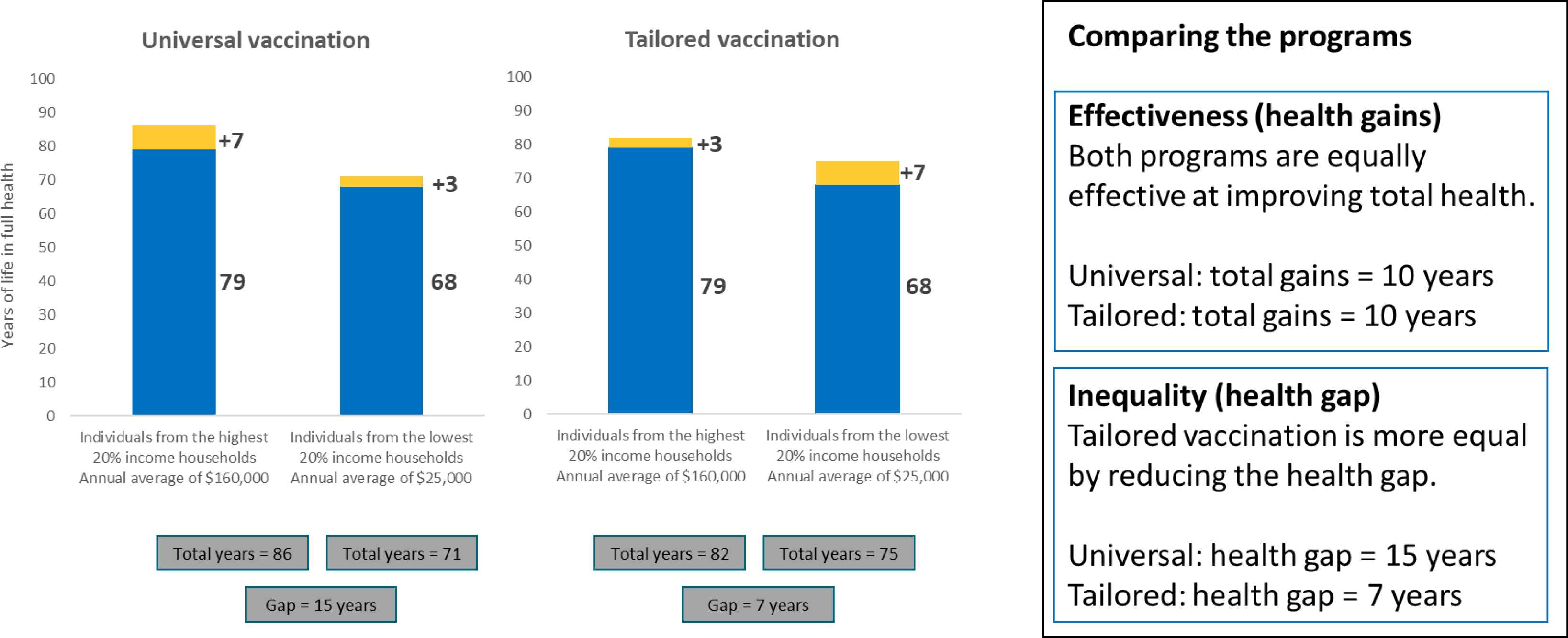
8. Which program would you choose?

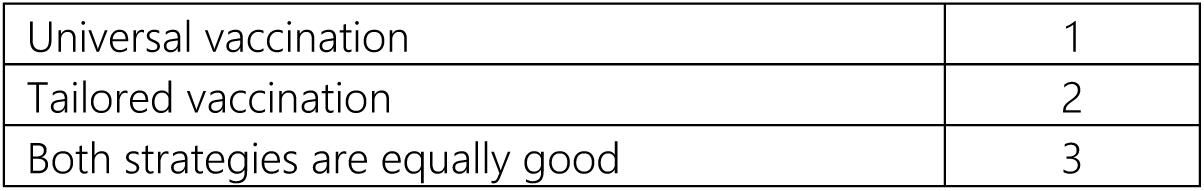

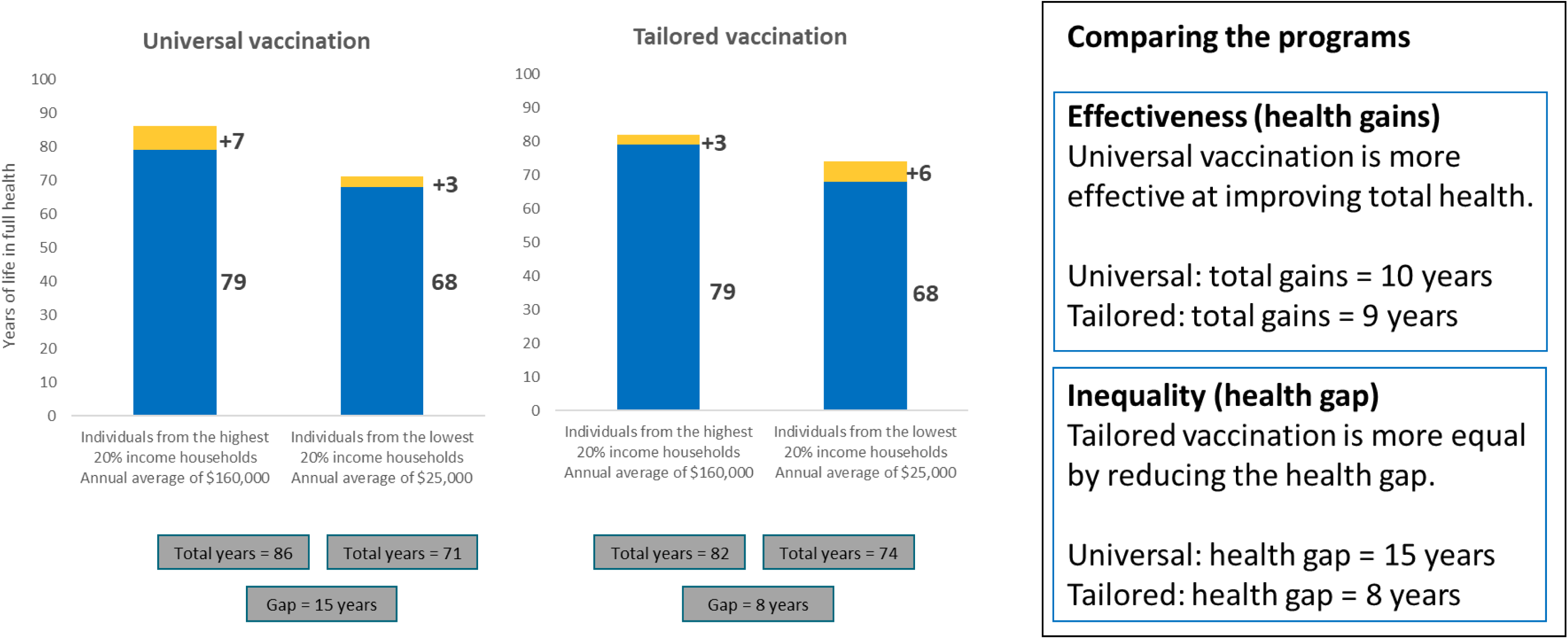
9. Which program would you choose?

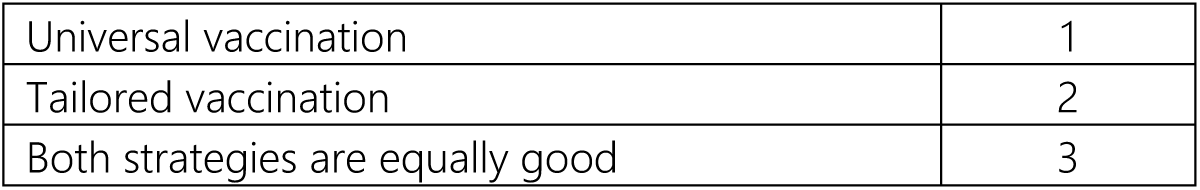

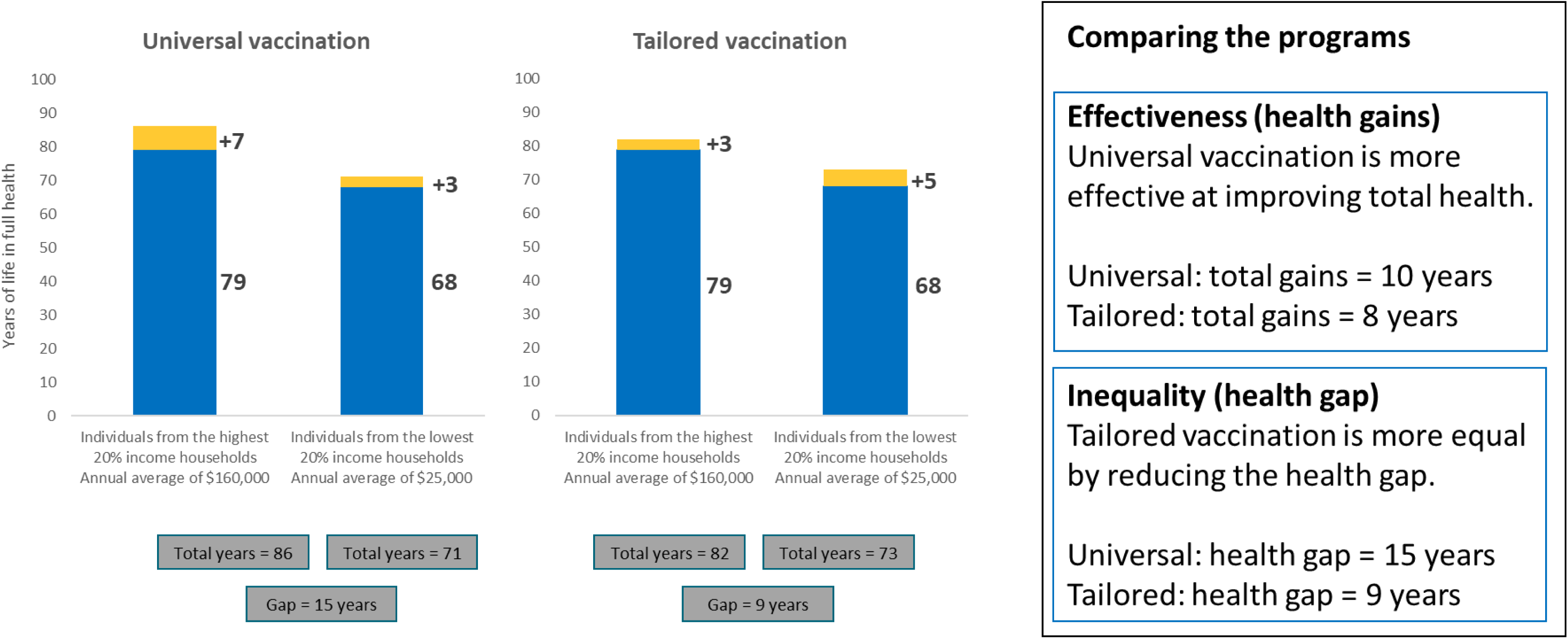
10. Which program would you choose?

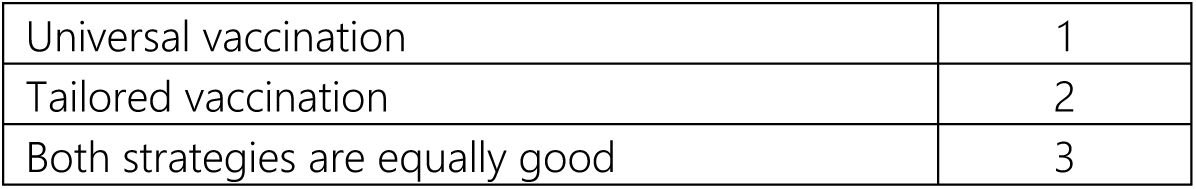

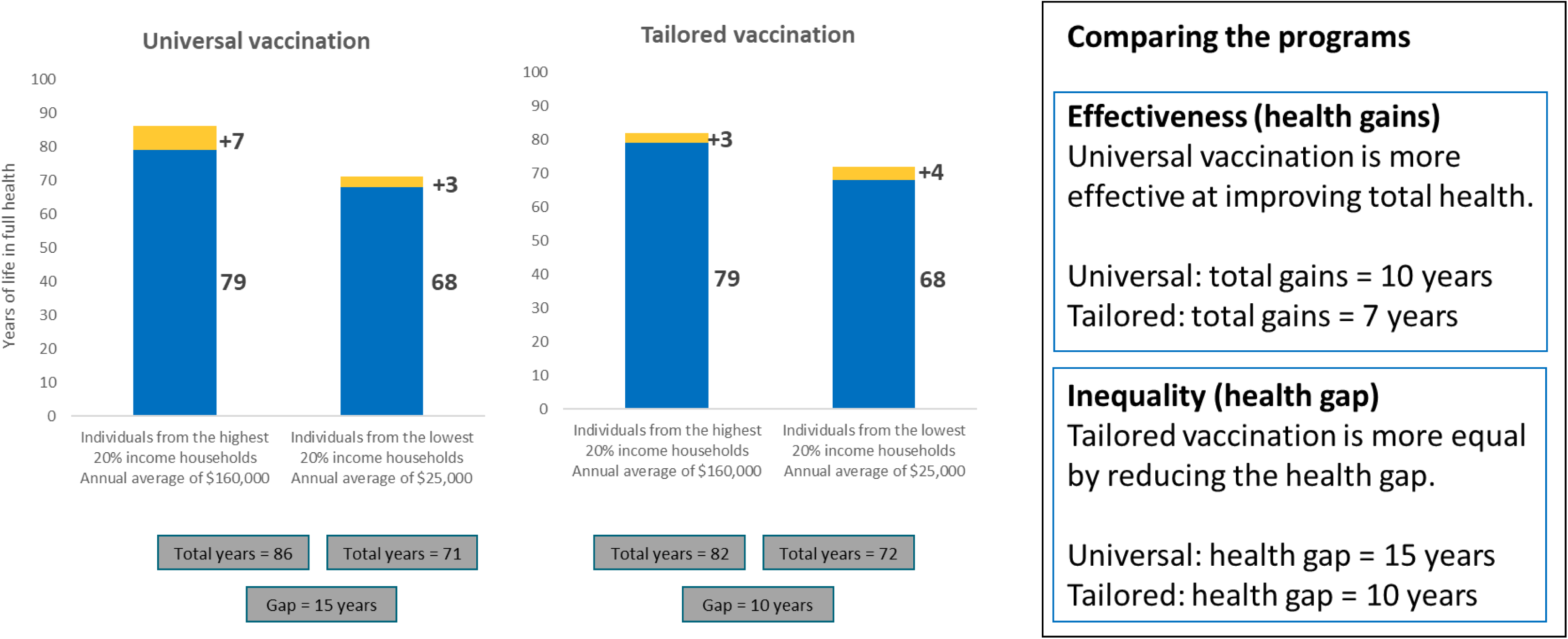
11. Which program would you choose?

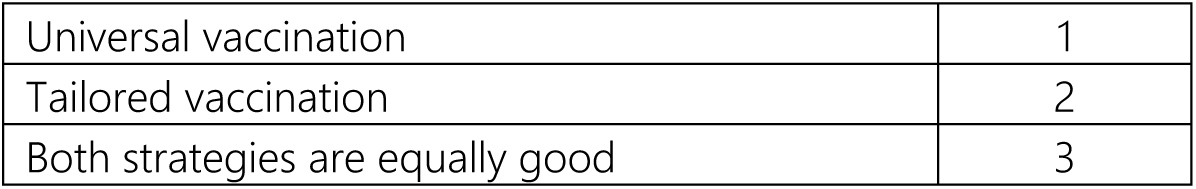

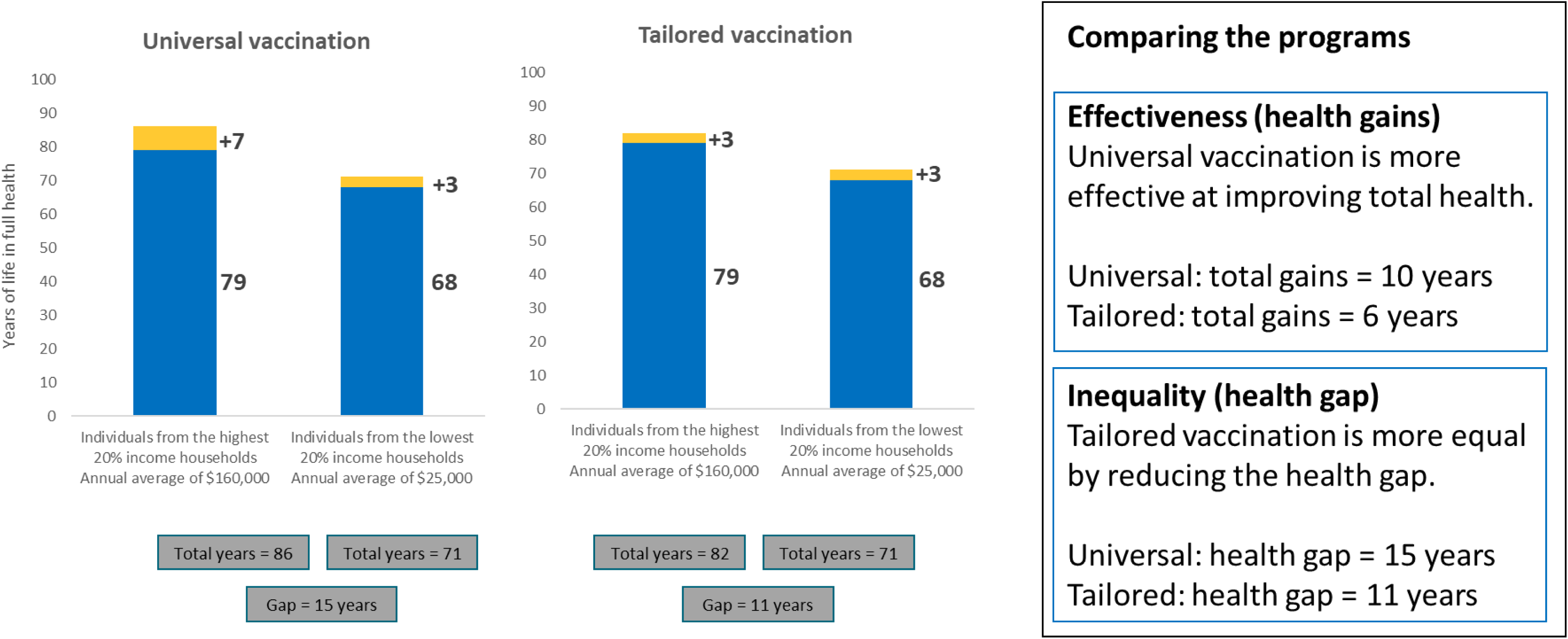
12. Which program would you choose?

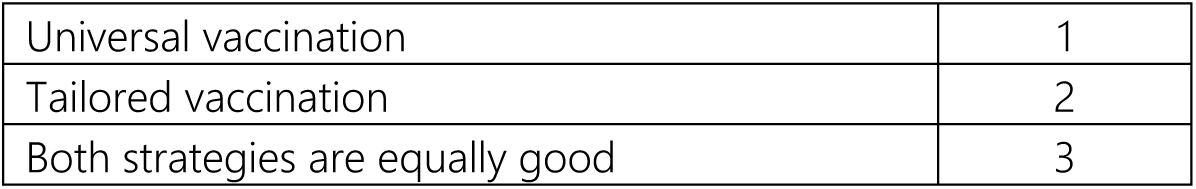

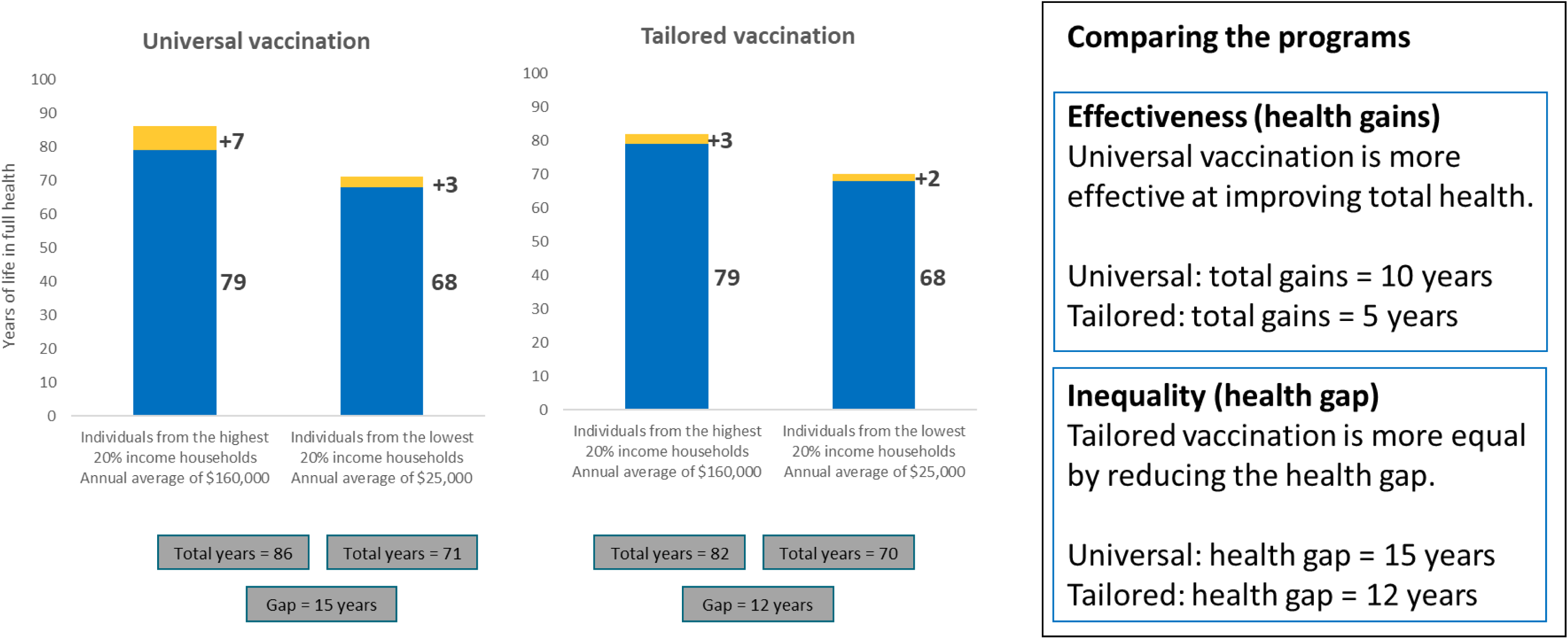
13. Which program would you choose?

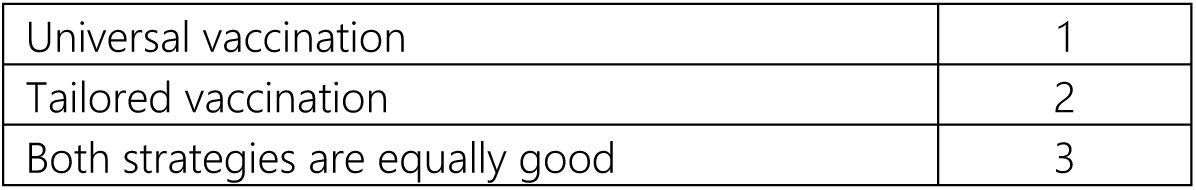

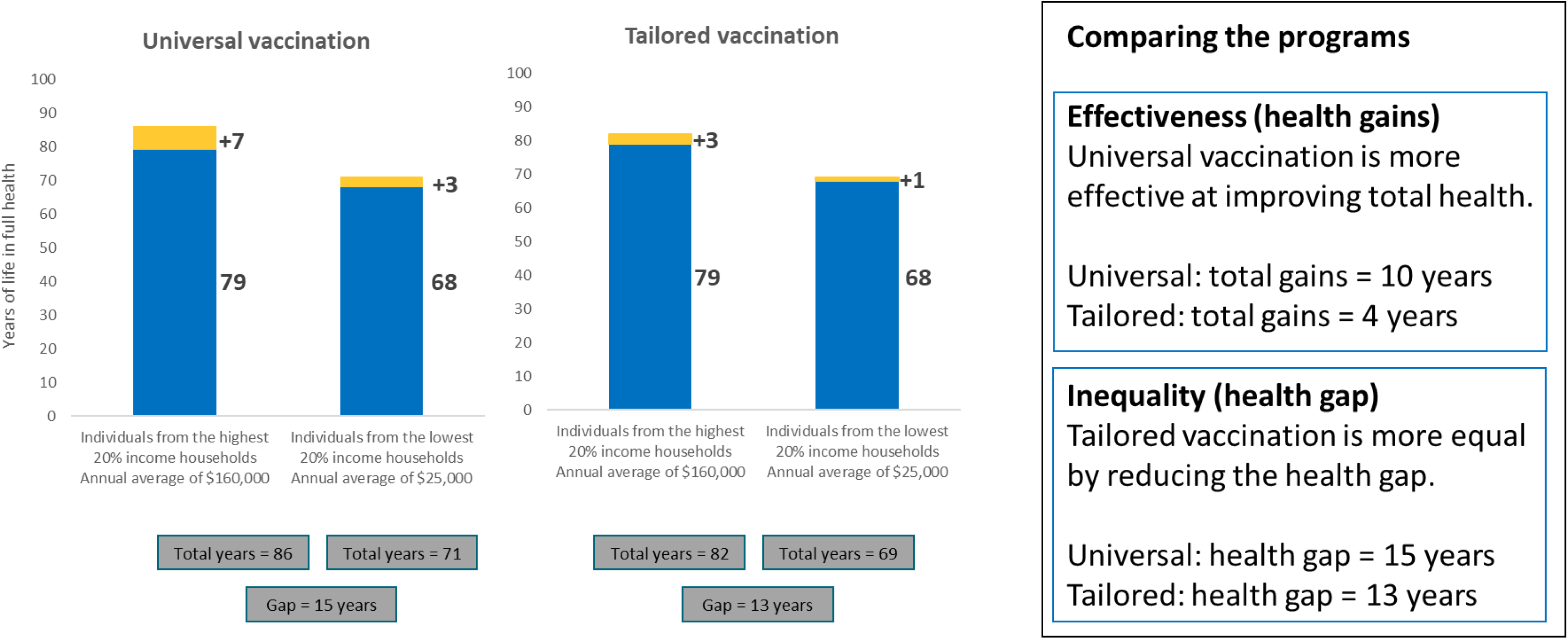
14. Which program would you choose?

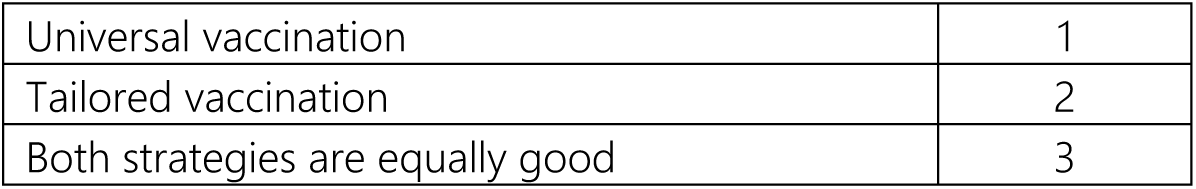 General information This information will help us understand whether people of different backgrounds may have different views. Your responses will be kept strictly anonymous and used only for research purposes.
15. How would you self-identify with respect to ethnicity and/or ancestry?

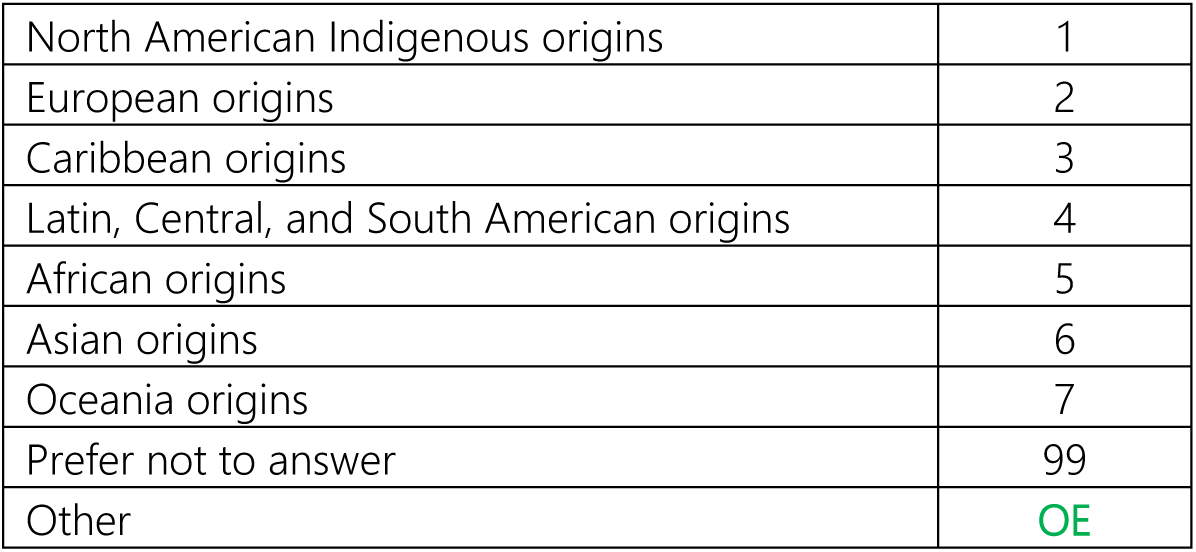
16. How do you self-identify with respect to racial origin/lineage?

17. What is your highest level of education attainment?

16. How do you self-identify with respect to racial origin/lineage?
|  |  |
| --- | --- |
| Indigenous (please specify) | OE |
| Black | 1 |
| East Asian | 2 |
| Latino | 3 |
| Middle eastern | 4 |
| South Asian | 5 |
| Southeast Asian | 6 |
| White | 7 |
| Prefer not to answer | 99 |
| Other | OE |

18. What was your total household income before taxes last year?

17. What is your highest level of education attainment?
|  |  |
| --- | --- |
| No certificate, diploma, or degree | 1 |
| Secondary (high) school diploma or equivalent | 2 |
| Apprenticeship or trades certificate | 3 |
| College, CEGEP, or other non-university certificate | 4 |
| University certificate or diploma below bachelor level | 5 |
| University certificate or diploma at bachelor level or above | 6 |
| Prefer not to answer | 99 |

19. Including yourself, how many family members live in your household?

18. What was your total household income before taxes last year?
|  |  |
| --- | --- |
| \$0 - \$29,000 | 1 |
| \$30,000 - \$49,999 | 2 |
| \$50,000 - \$69,999 | 3 |
| \$70,000 - \$99,999 | 4 |
| \$100,000 - \$149,999 | 5 |
| \$150,000 or more | 6 |
| Prefer not to answer | 99 |

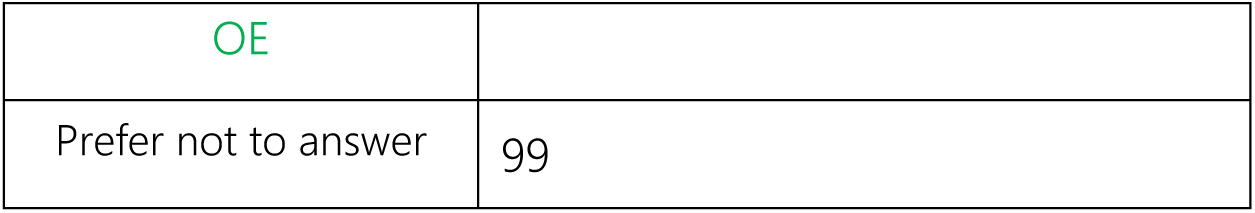 Vaccination-related questions This questionnaire presented a series of hypothetical scenarios that involve public policies related to vaccination. The following questions relate to your opinion towards vaccination. Your responses will be kept strictly anonymous and used only for research purposes.
20. Have you been vaccinated in the past three (3) years? (for example, vaccine for hepatitis, streptococcus pneumoniae, influenza, etc.)

21. What are the main reasons why you haven’t received a vaccine in the past three years?

|  |  |
| --- | --- |
| Yes | 1 |
| No | 2 |
| Prefer not to say | 99 |

22. What are the main reasons you don’t want to be vaccinated?

**21. What are the main reasons why you haven't received a vaccine in the past three years?**
|  |  |
| --- | --- |
| I do not want to be vaccinated at this time | 1 |
| I do not want to be vaccinated at all | 2 |
| I have an appointment in the future | 3 |
| I haven't been able to get an appointment yet | 4 |
| I don't know where to get vaccinated | 5 |
| I don't think it's necessary | 6 |
| Prefer not to answer | 99 |
| Other | OE |

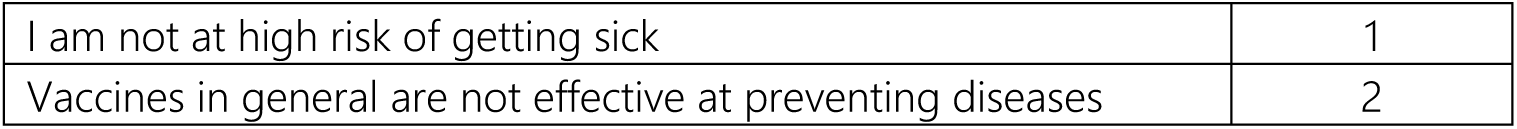
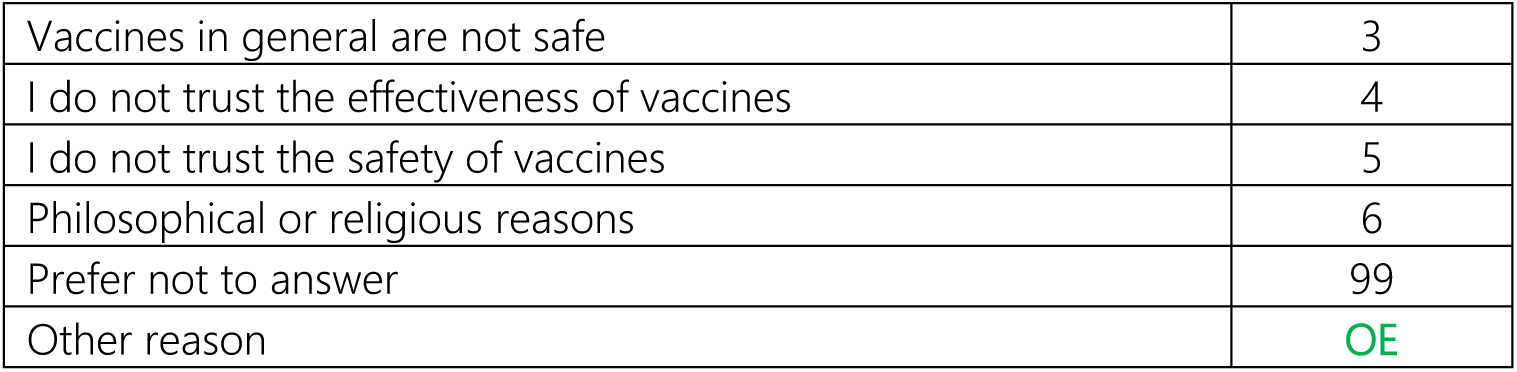 Vaccination
23. Please use this space to share any comments regarding vaccination policies in Canada

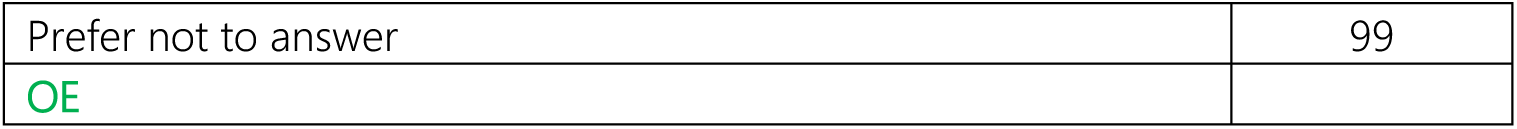 Final comments Please answer the following questions
24. How clear were the instructions to complete this survey?

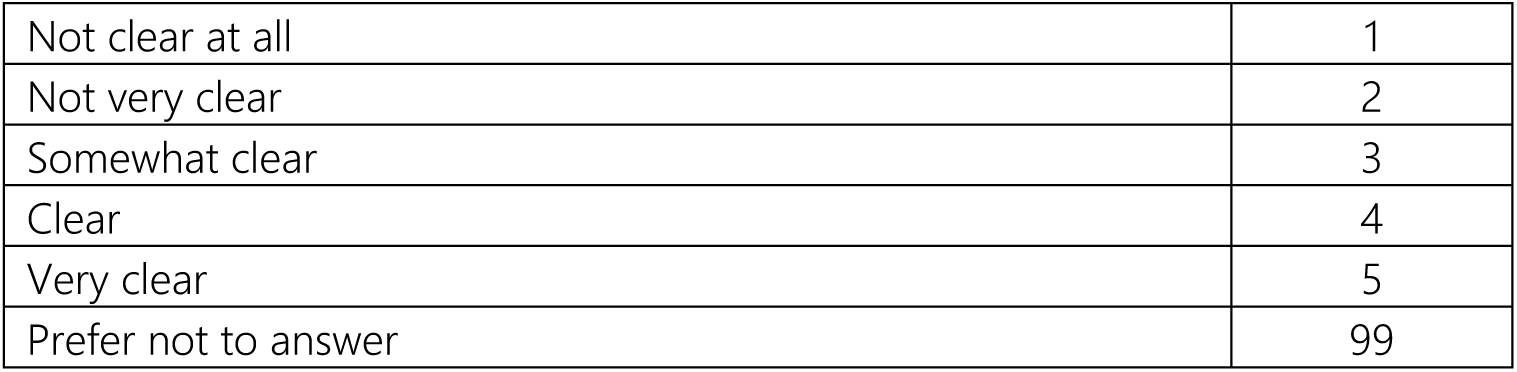
25. How clear were the definitions for the tailored and universal vaccination programs?

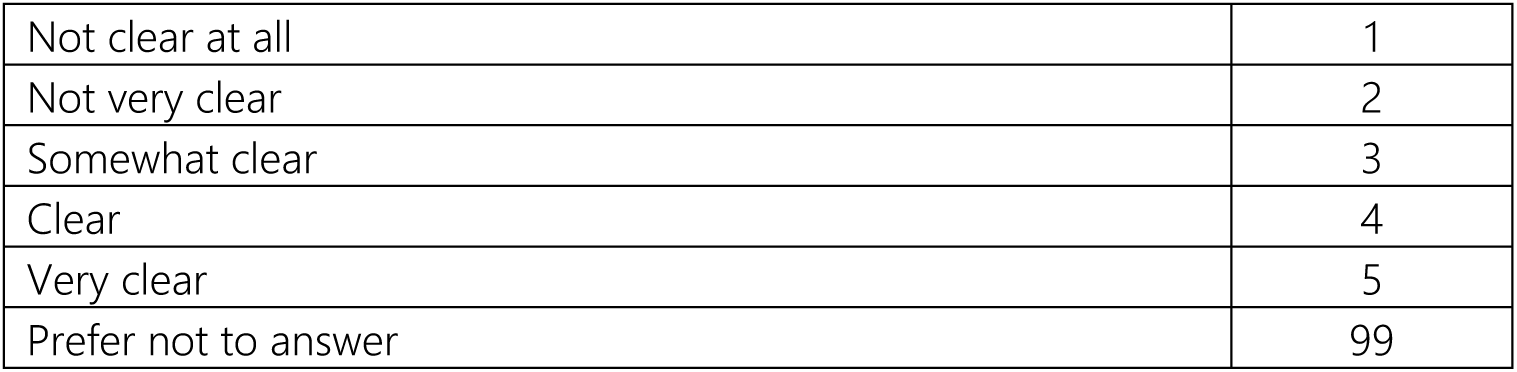
26. Please use this space to make any final comments.

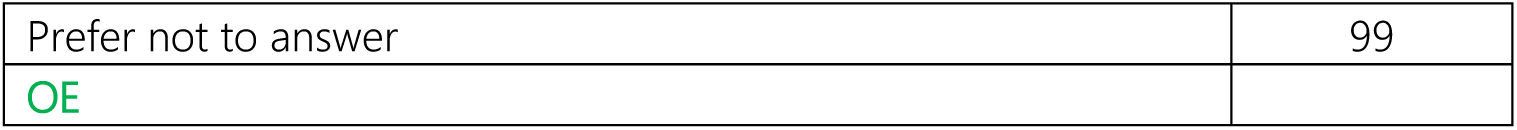

## Appendix 4: Prevention survey

### Understanding public views on funding the health system

This questionnaire should take about 10-15 minutes to complete.

Your responses will be kept strictly anonymous and used only for research purposes.

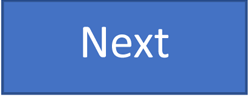

We will ask four questions to assess your eligibility for the survey. Your responses will be kept strictly anonymous and used only for research purposes.

1. Your age?

2. Your gender?

|  |  |
| --- | --- |
| OE | TERMINATE if age<br>< 18 |
| Prefer not<br>to say | 99 |

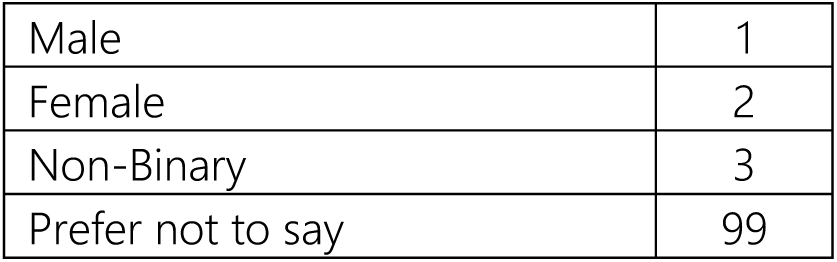
3. In which province/territory do you live?

4. What are the first three characters of your postal code? (Format A1A)

**3. In which province/territory do you live?**
|  |  |  |
| --- | --- | --- |
| Alberta | 1 |  |
| British Columbia | 2 |  |
| Manitoba | 3 |  |
| Newfoundland and Labrador | 4 |  |
| New Brunswick | 5 |  |
| Northwest Territories | 6 |  |
| Nova Scotia | 7 |  |
| Nunavut | 8 |  |
| Ontario | 9 |  |
| Prince Edward Island | 10 |  |
| Quebec | 11 |  |
| Saskatchewan | 12 |  |
| Yukon Territory | 13 |  |
| I live outside of Canada | 14 | TERMINATE |
| Prefer not to say | 99 |  |

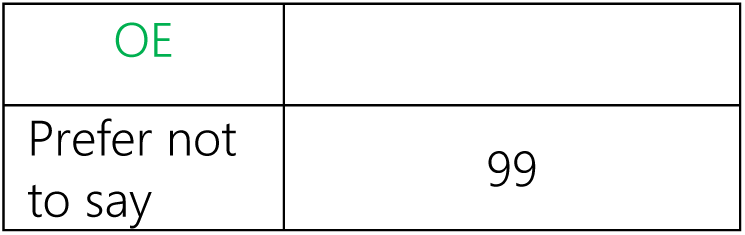 In the next few screens, you will be provided with an introduction to this survey, with some information for you to review. You will then be asked some questions based on this information. Introduction There are differences between populations with the **highest and lowest household incomes** in Canada in terms of how long people live, their quality of life, and their access to healthcare. If we divide the population into five groups of equal size (each group = 20% of the population), then in 2019:
  - The highest-earning 20% of the population had an average household income after tax of **$160,000** per year.
  - The lowest-earning 20% of the population had an average household income after tax of **$25,000** per year.

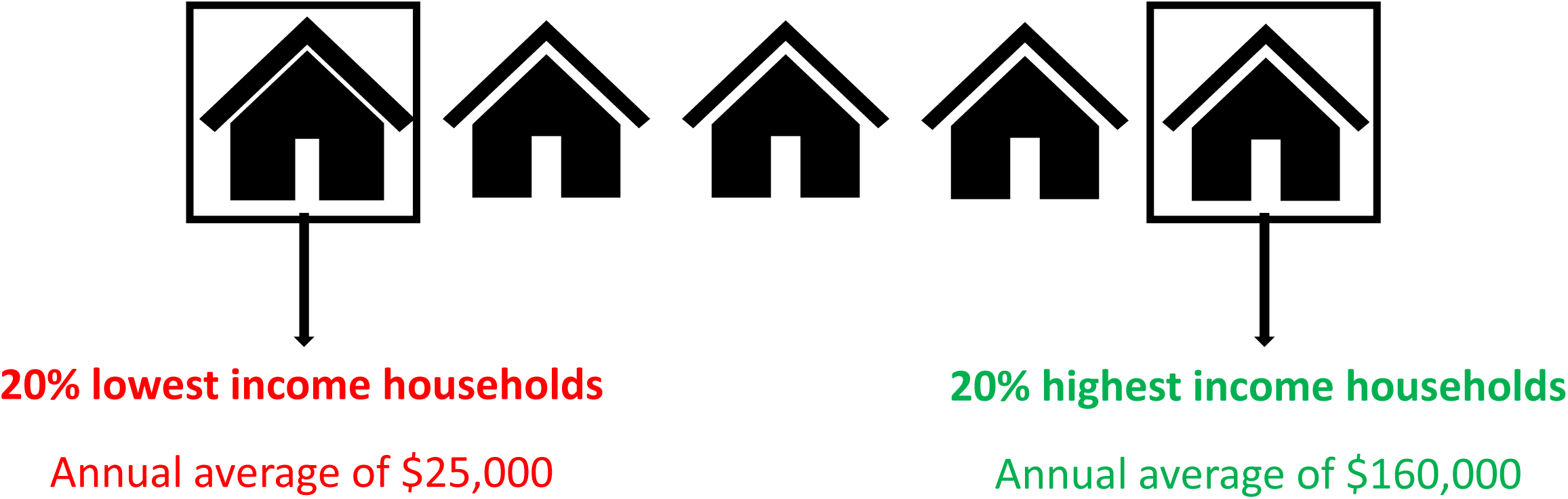
  - While actual length of life and health vary between individuals, on average, the **20% highest household income population** experience **79 years** of life in full health.
  - On the other hand, the **20% lowest household income population** experience **68 years** of life in full health.
  As an For example, someone who has 79 years in full health might have lived to 85 years old, but with less than full health towards the end of their life. These are averages across the whole population of Canada. Each individual’s actual length of life and health can vary considerably from these averages.

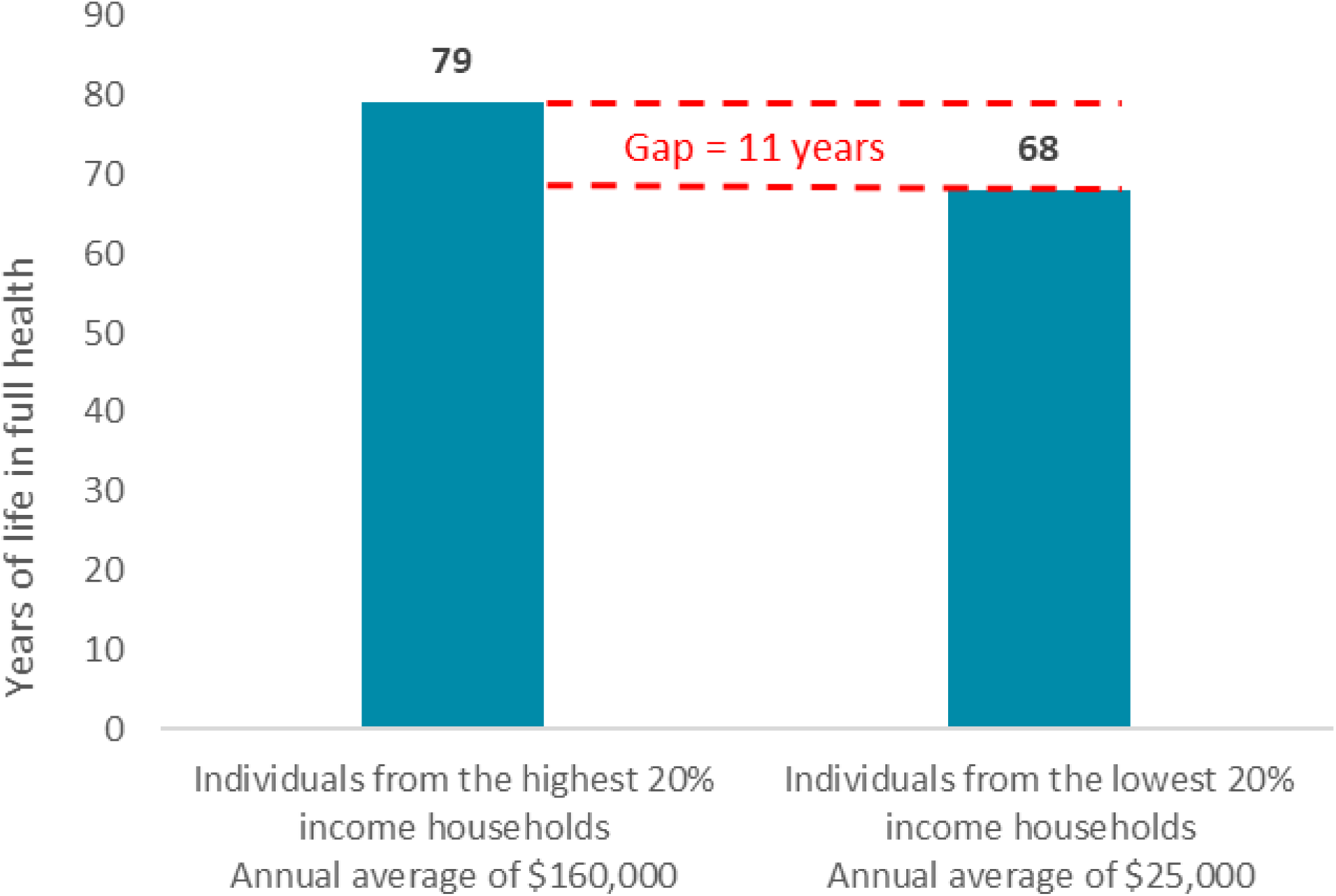

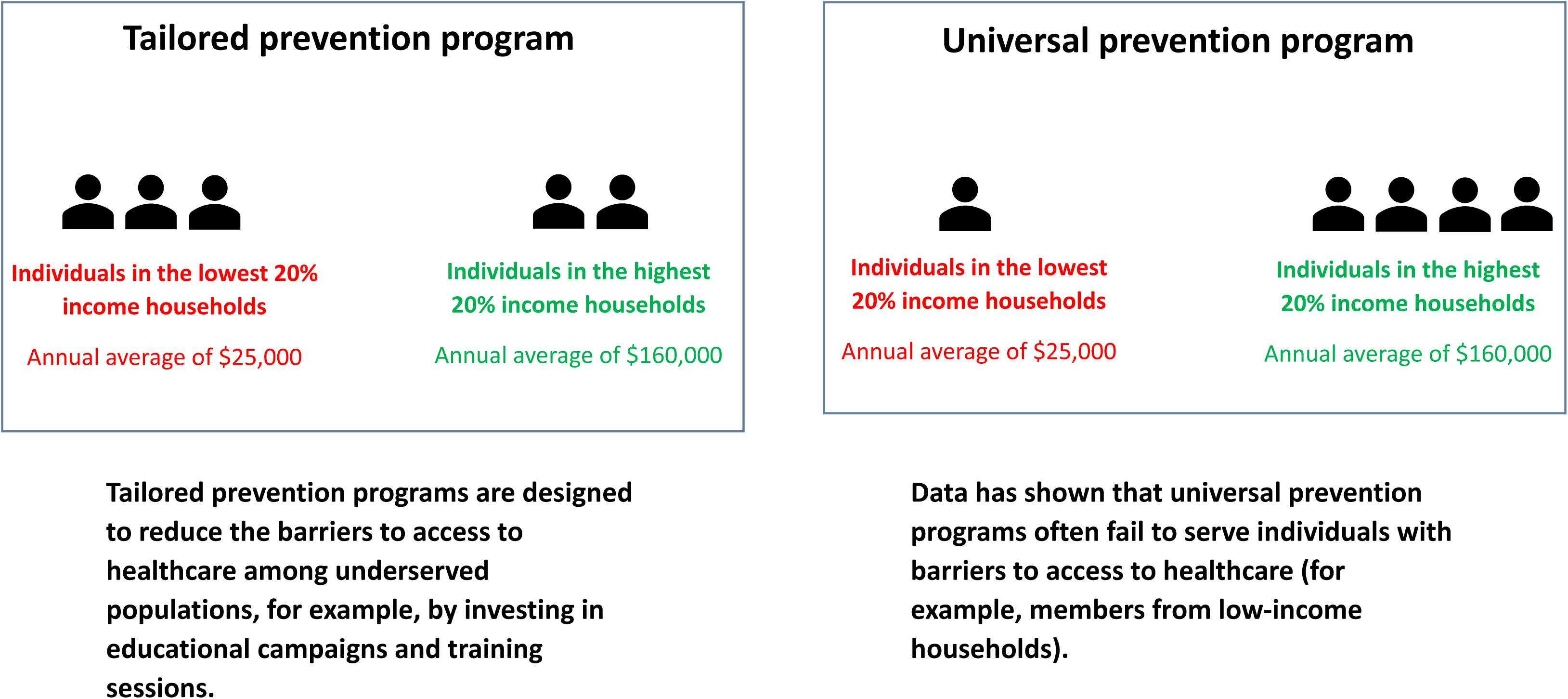

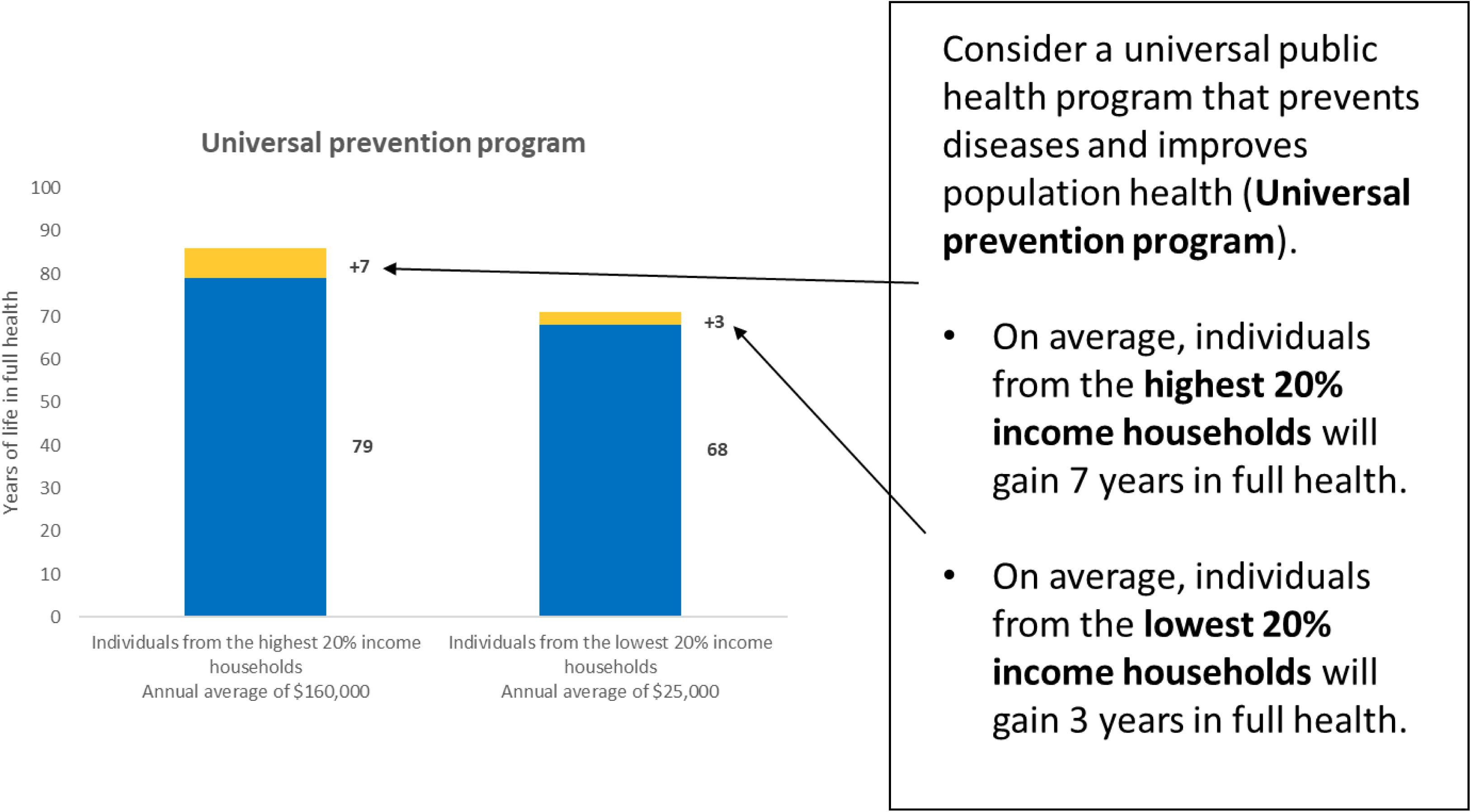

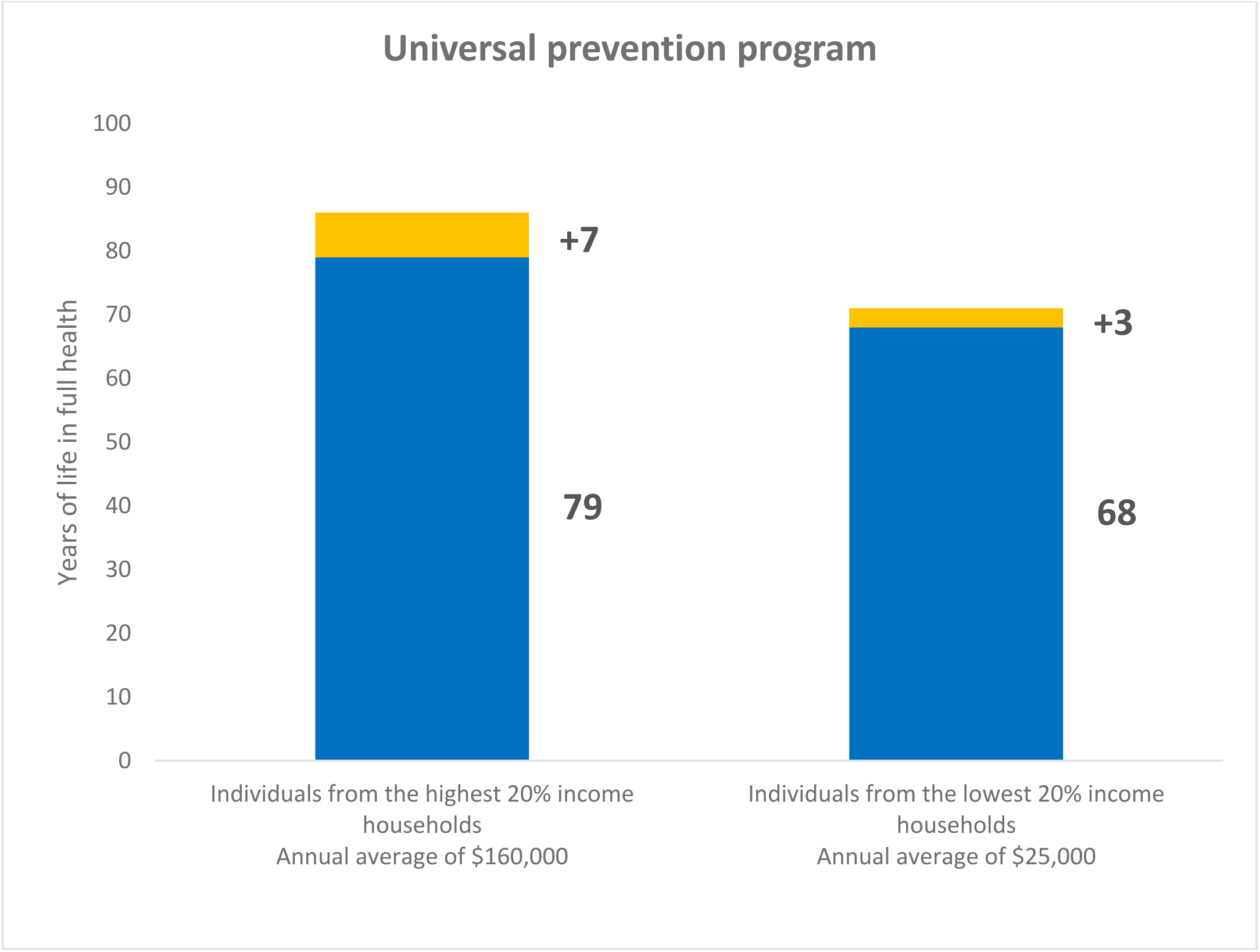
5. Which population group has the highest life expectancy AFTER implementing Universal prevention program?

|  |  |  |
| --- | --- | --- |
| Individuals in the highest 20% income households | 1 |  |
| Individuals in the lowest 20% income households | 2 | TERMINATE |

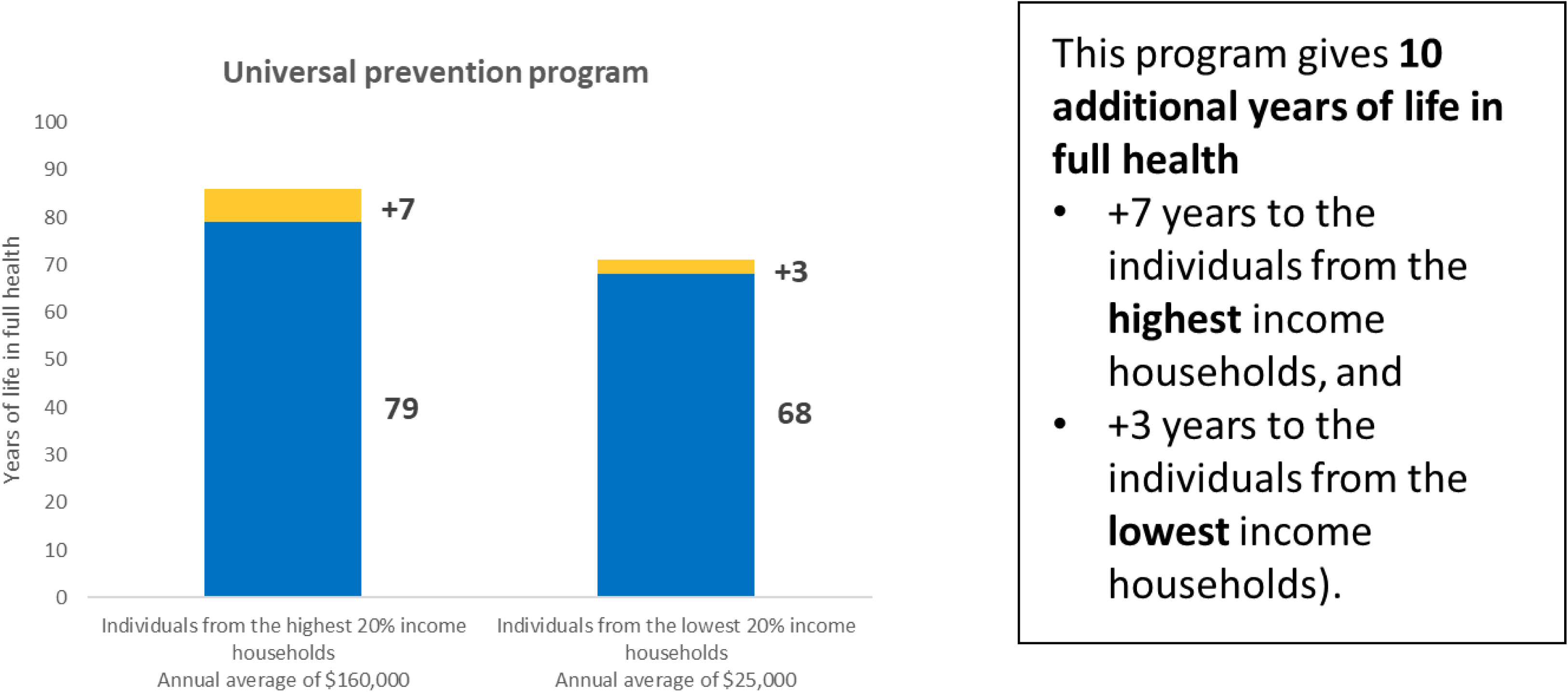

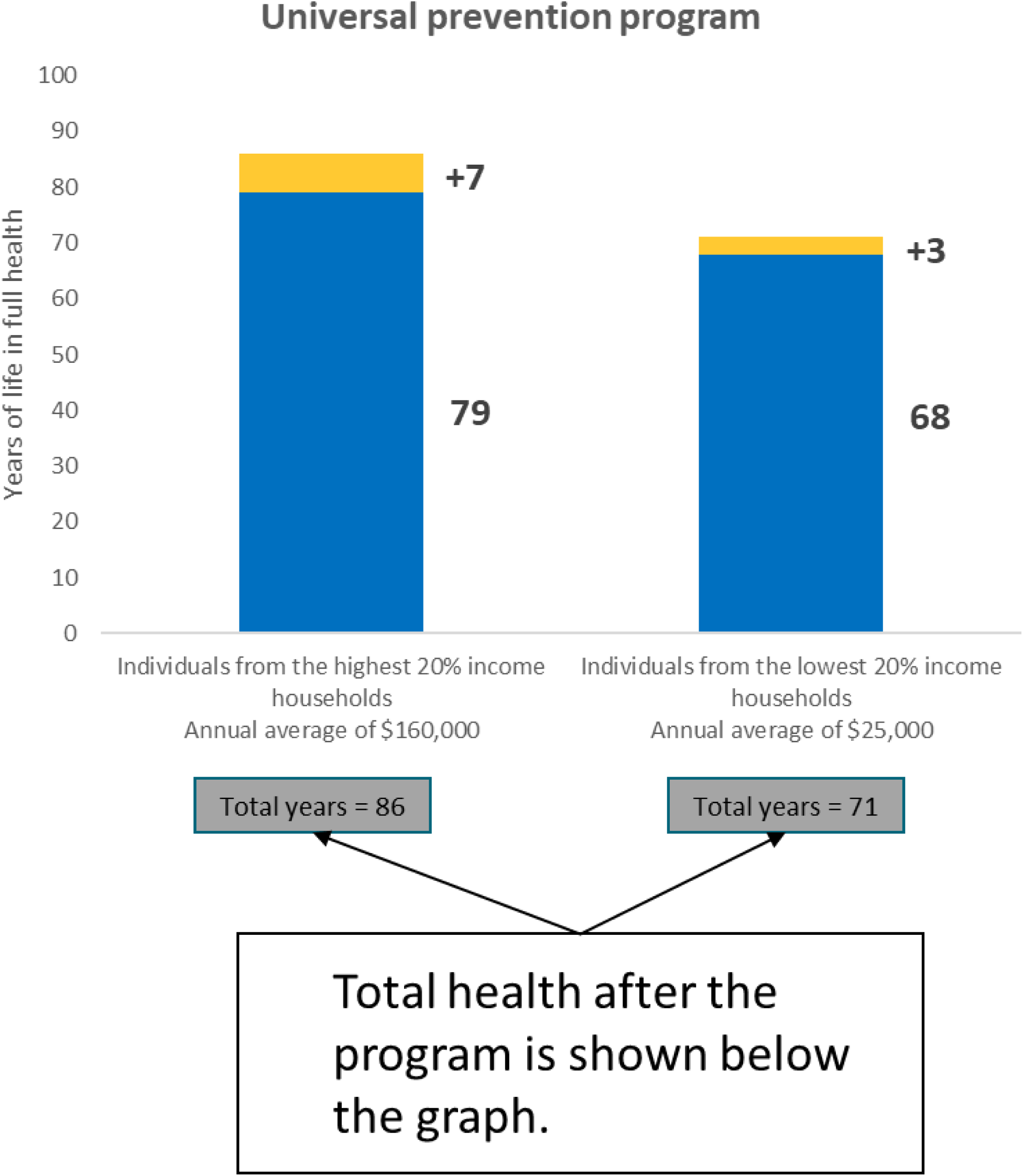

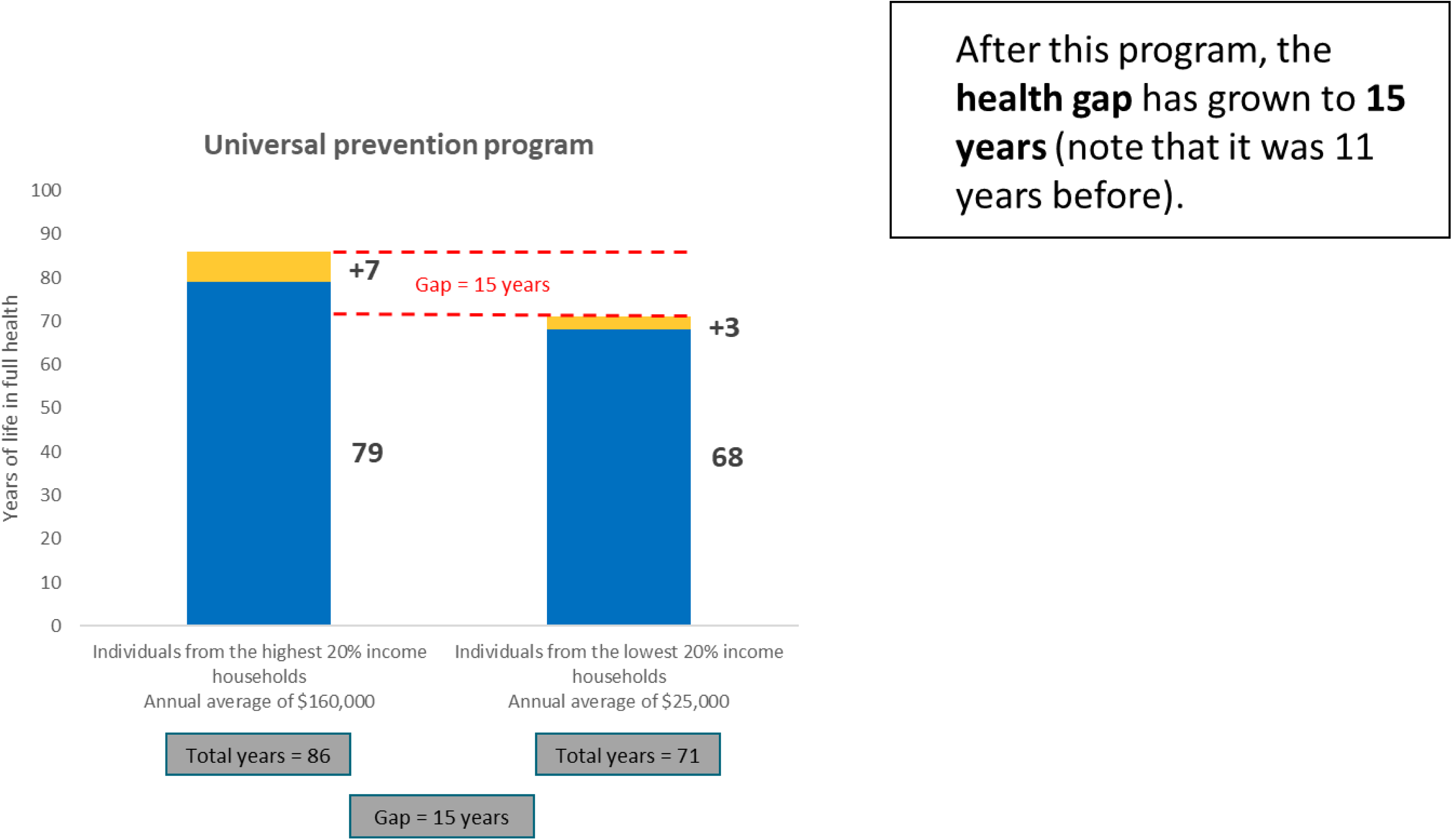

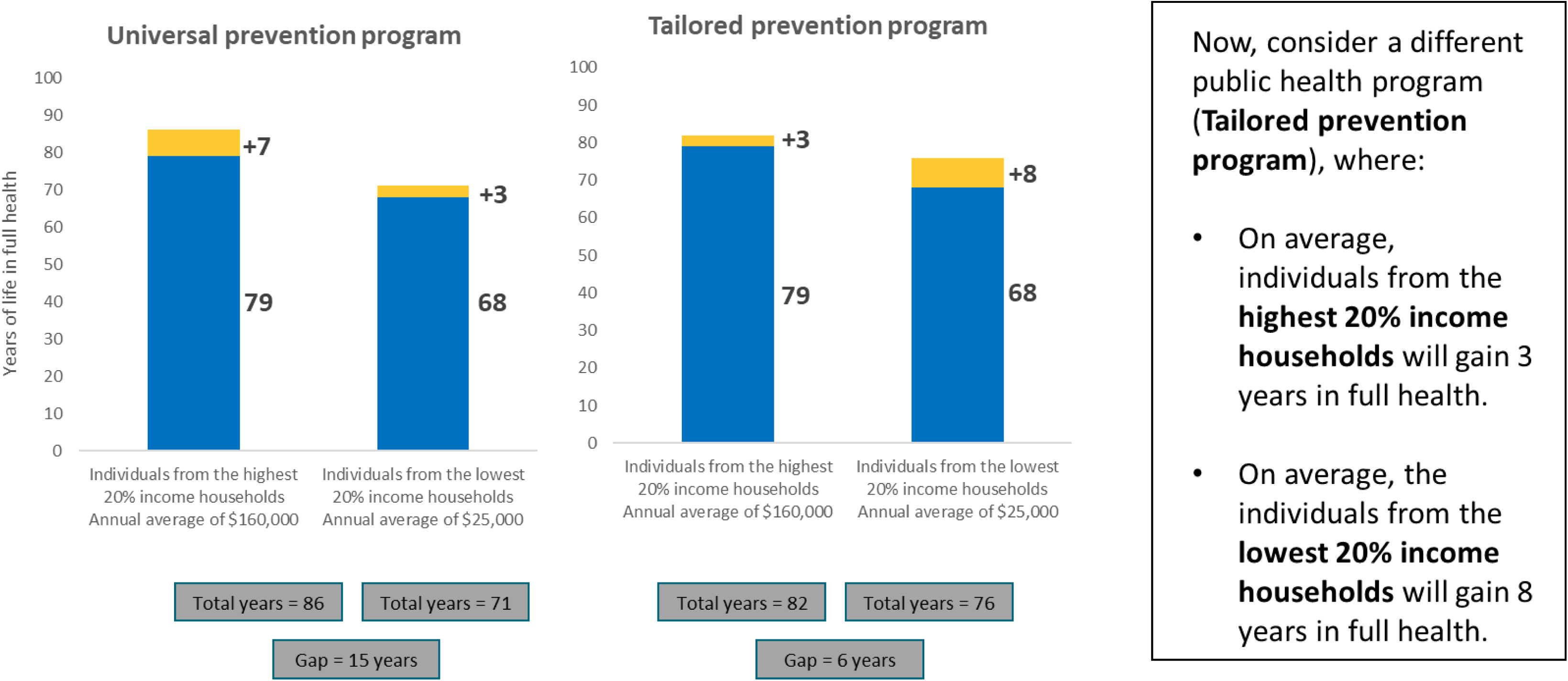

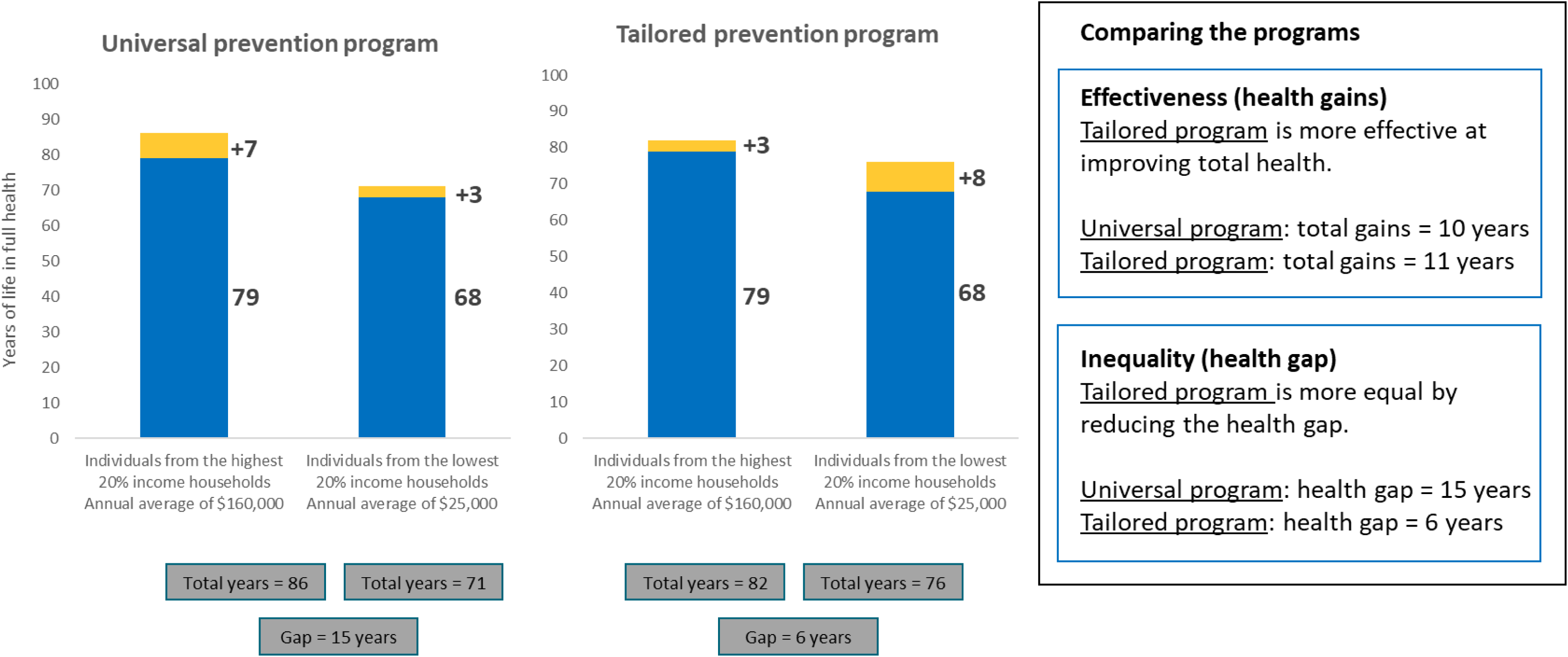

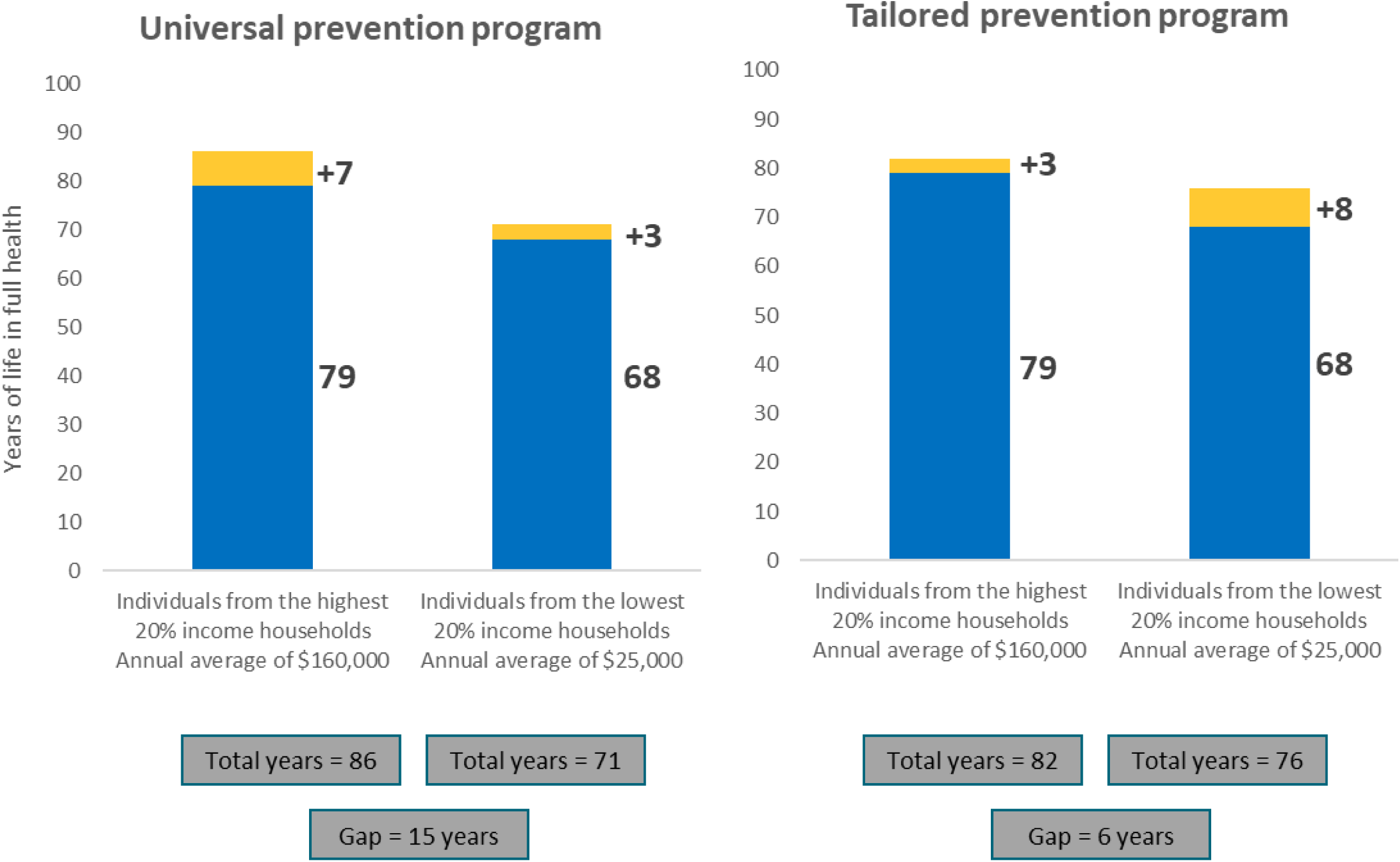
6. Which program would you choose?
7. 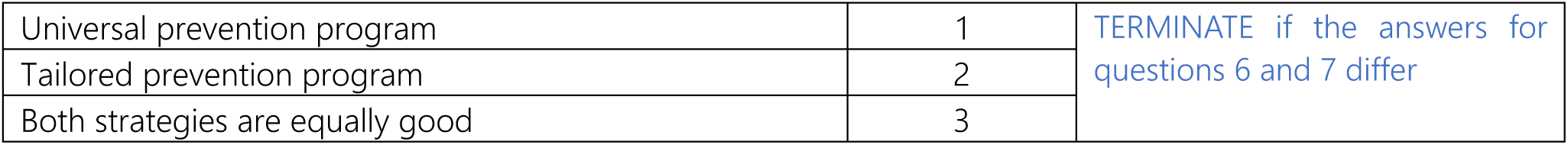 Which program should the government choose? Answer the next 8 scenarios considering:
  - We cannot pay for both programs — a choice must be made.
  - “Either program is good” means you don’t mind which one is chosen.
  - Both programs cost exactly the same.
  - The only difference between the programs is how much health is gained by the individuals in the highest and lowest 20% income households.
  - The population in the middle is not affected.

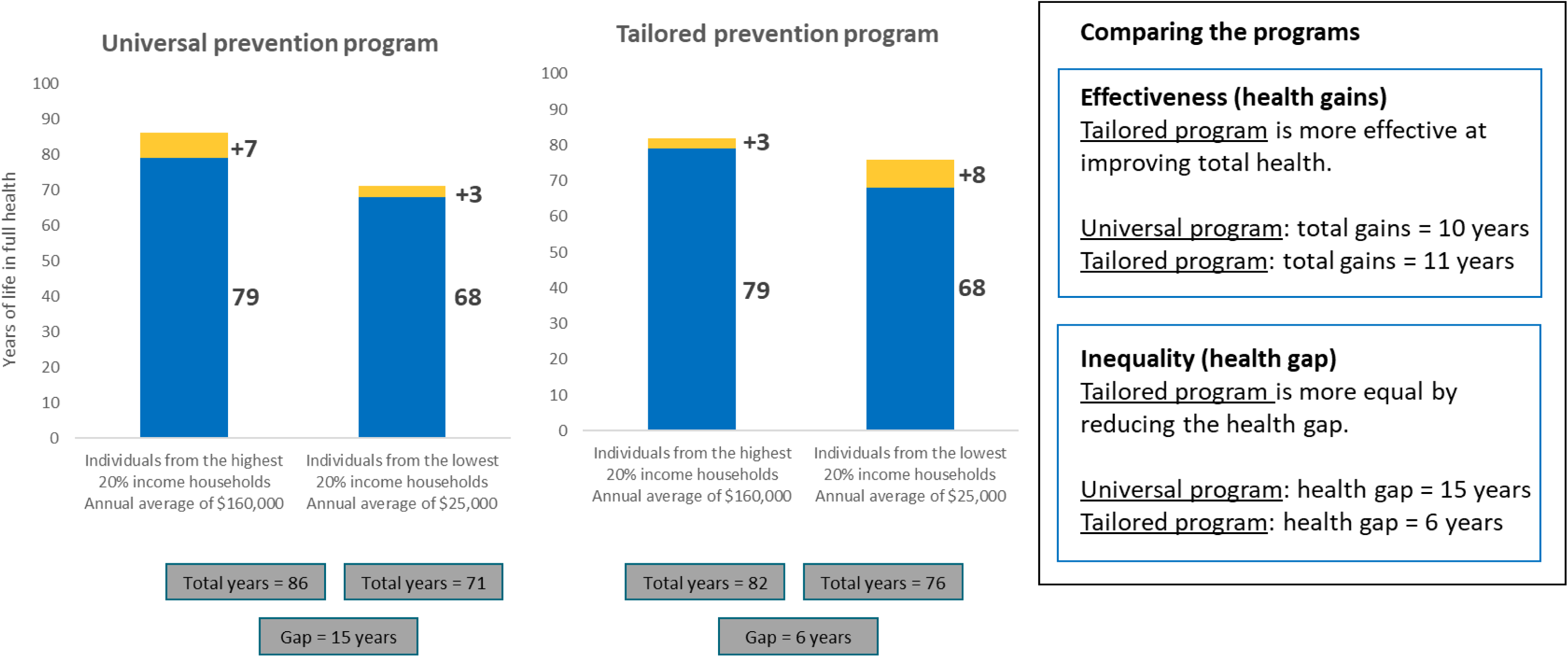
8. Which program would you choose?

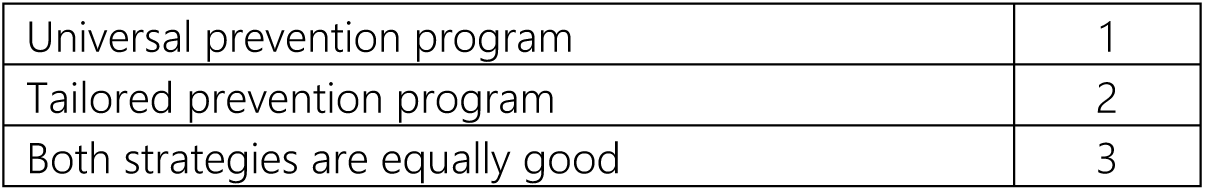

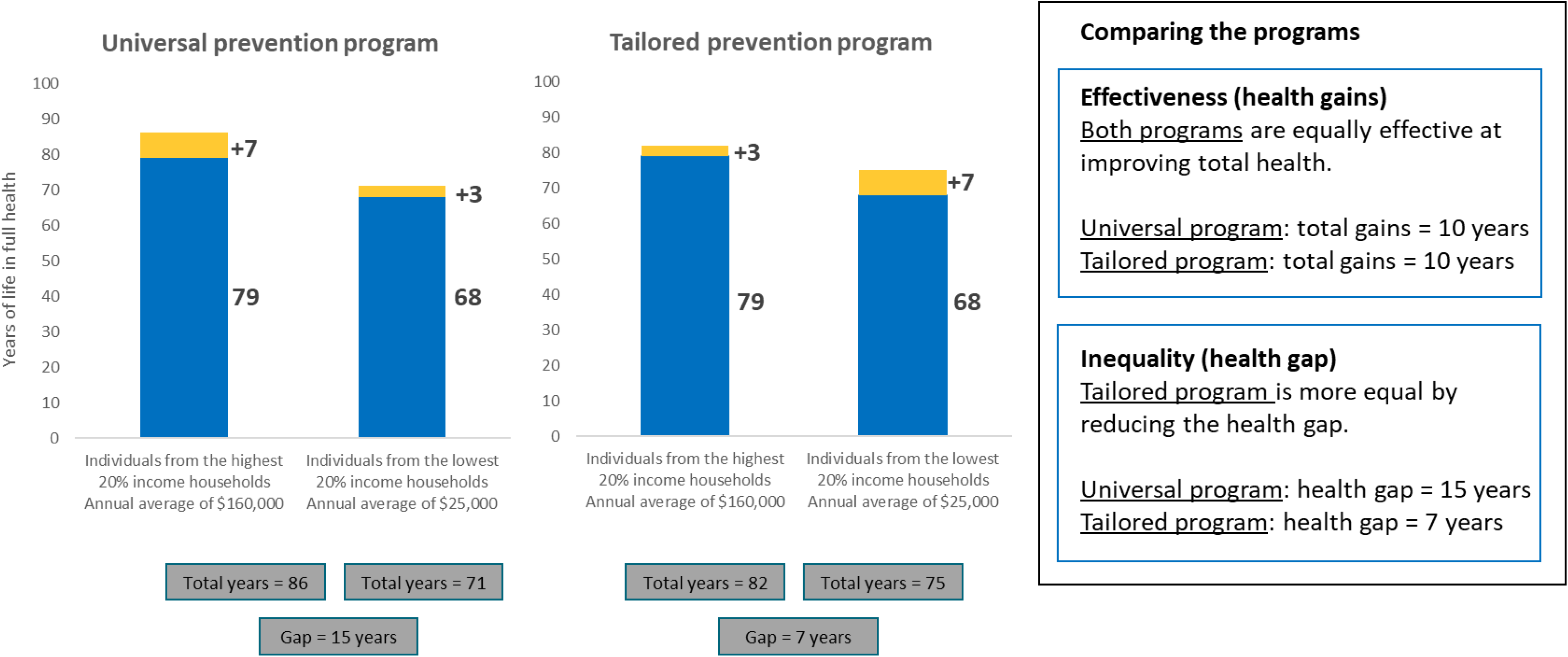
9. Which program would you choose?

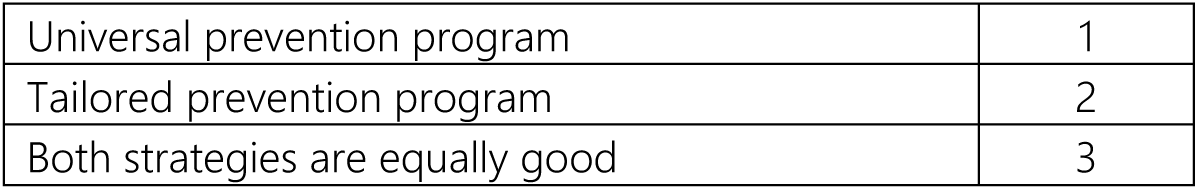

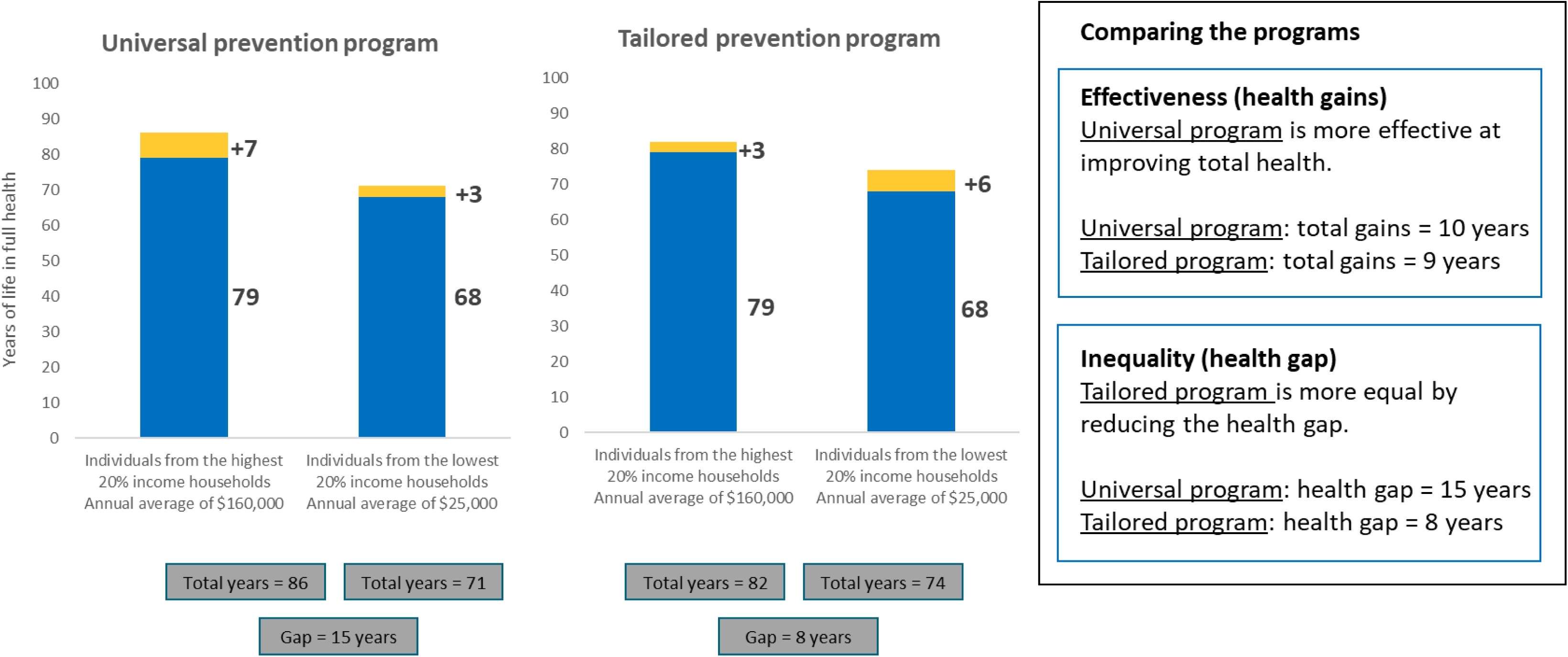
10. Which program would you choose?

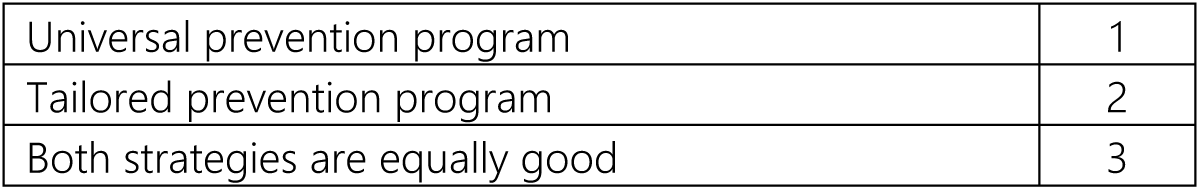

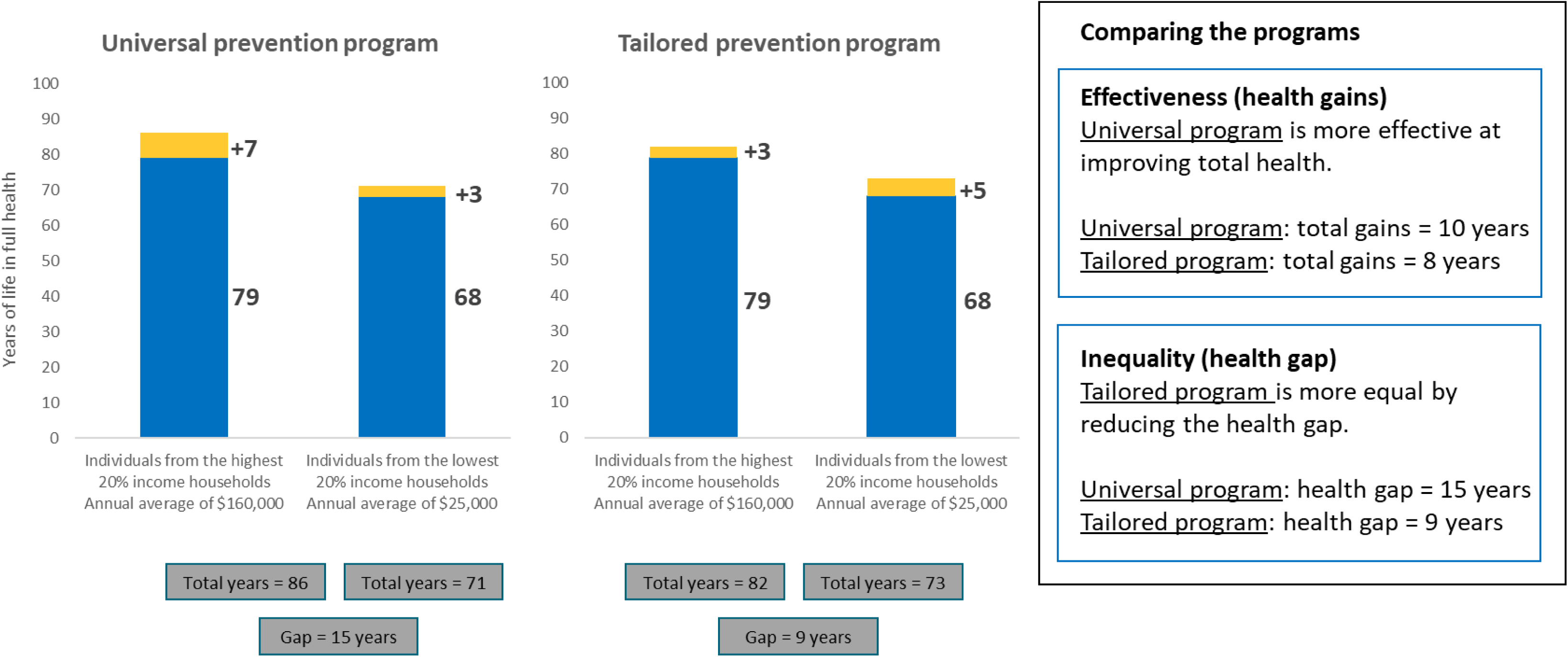
11. Which program would you choose?

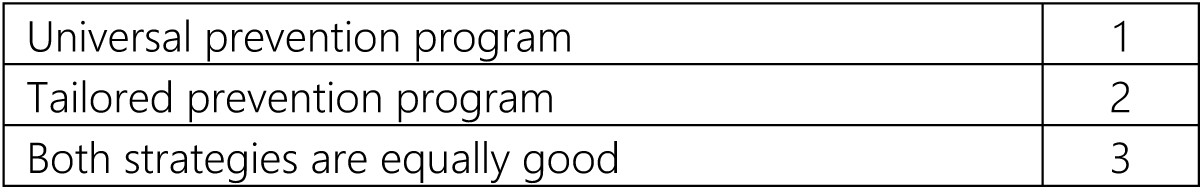

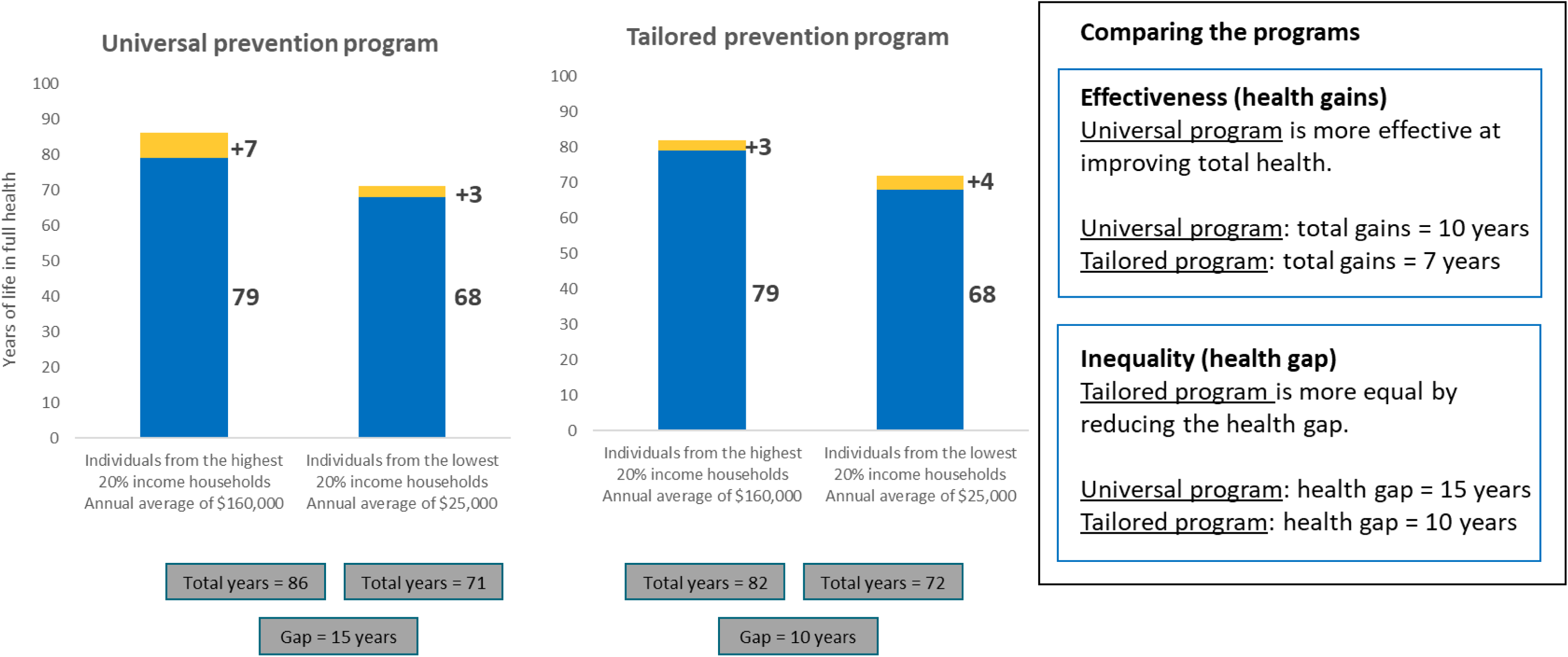
12. Which program would you choose?

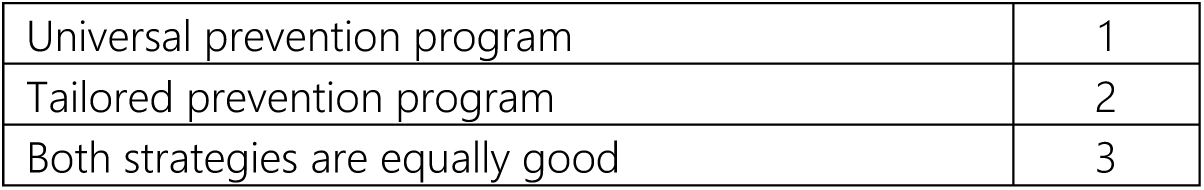

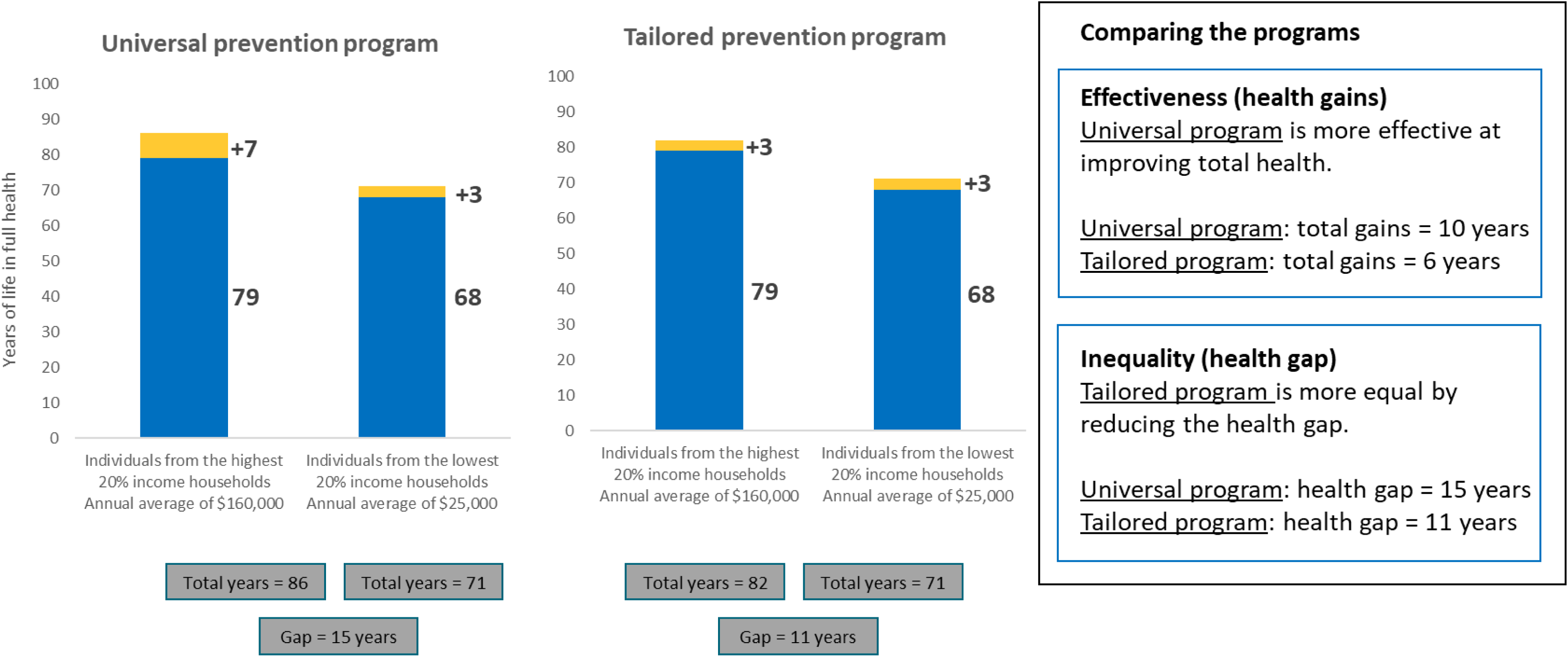
13. Which program would you choose?

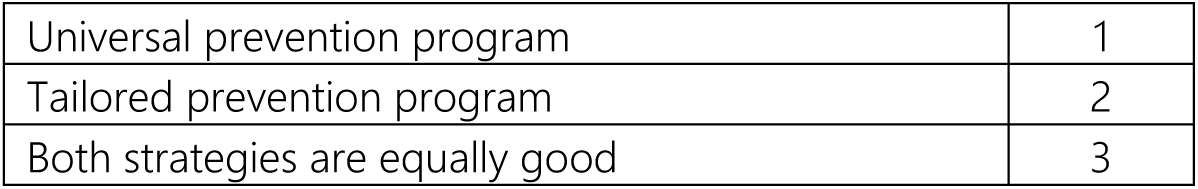

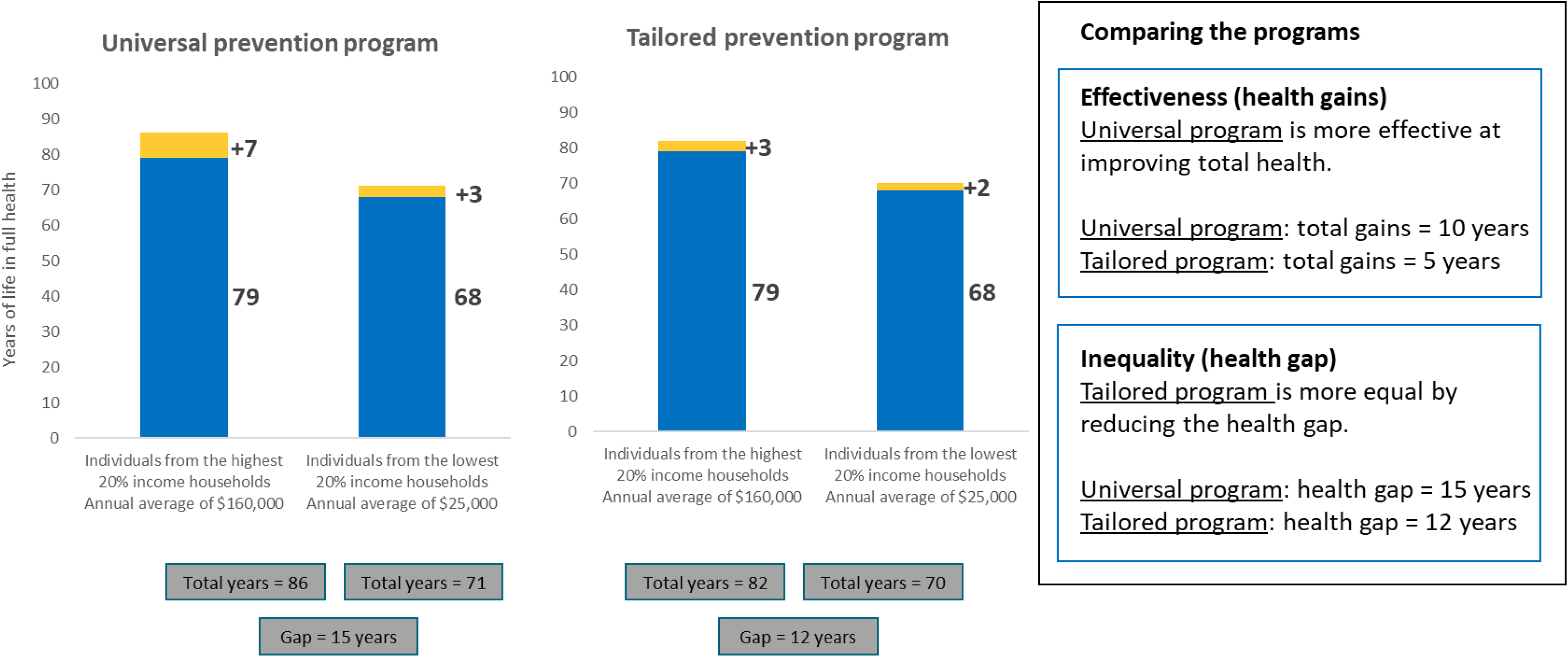
14. Which program would you choose?

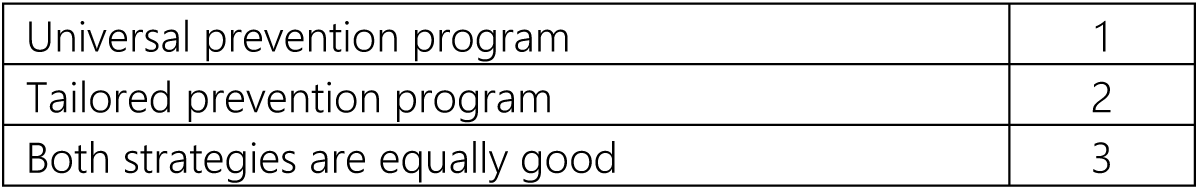

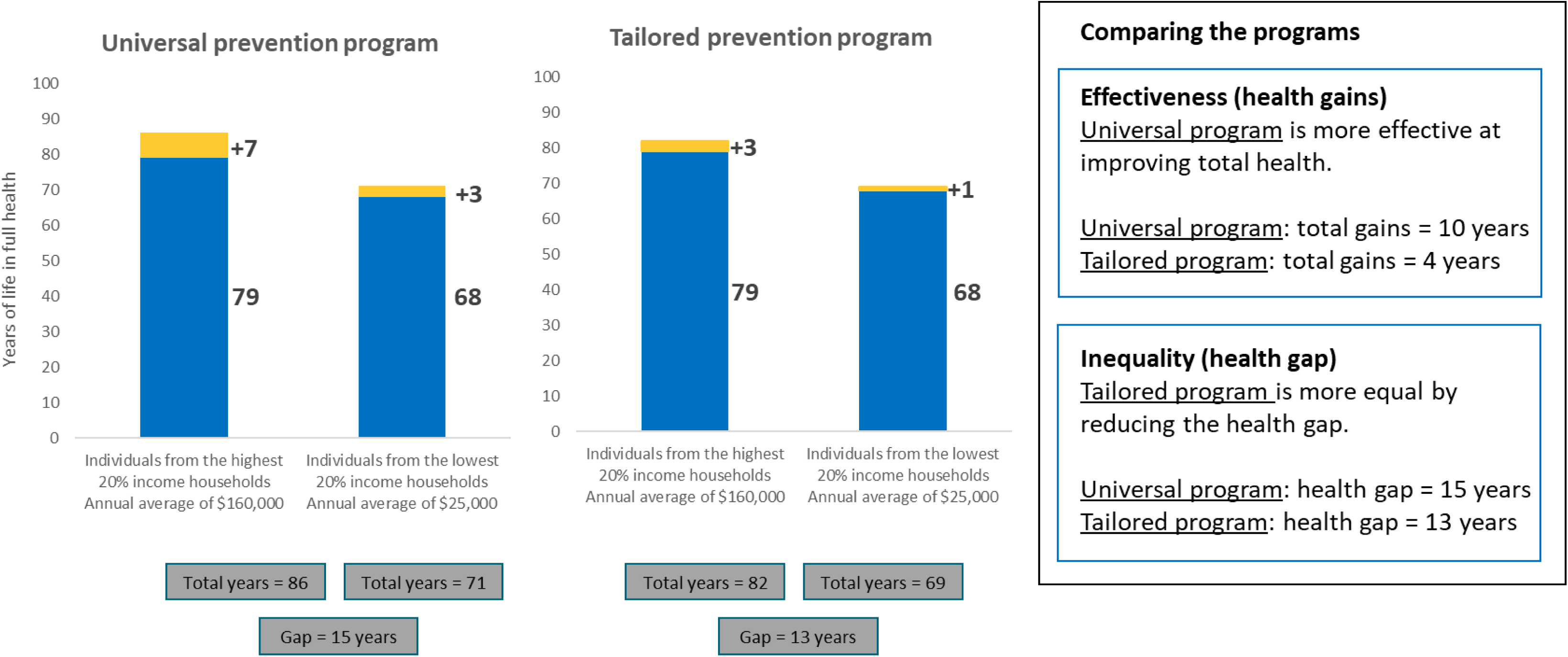
15. Which program would you choose?

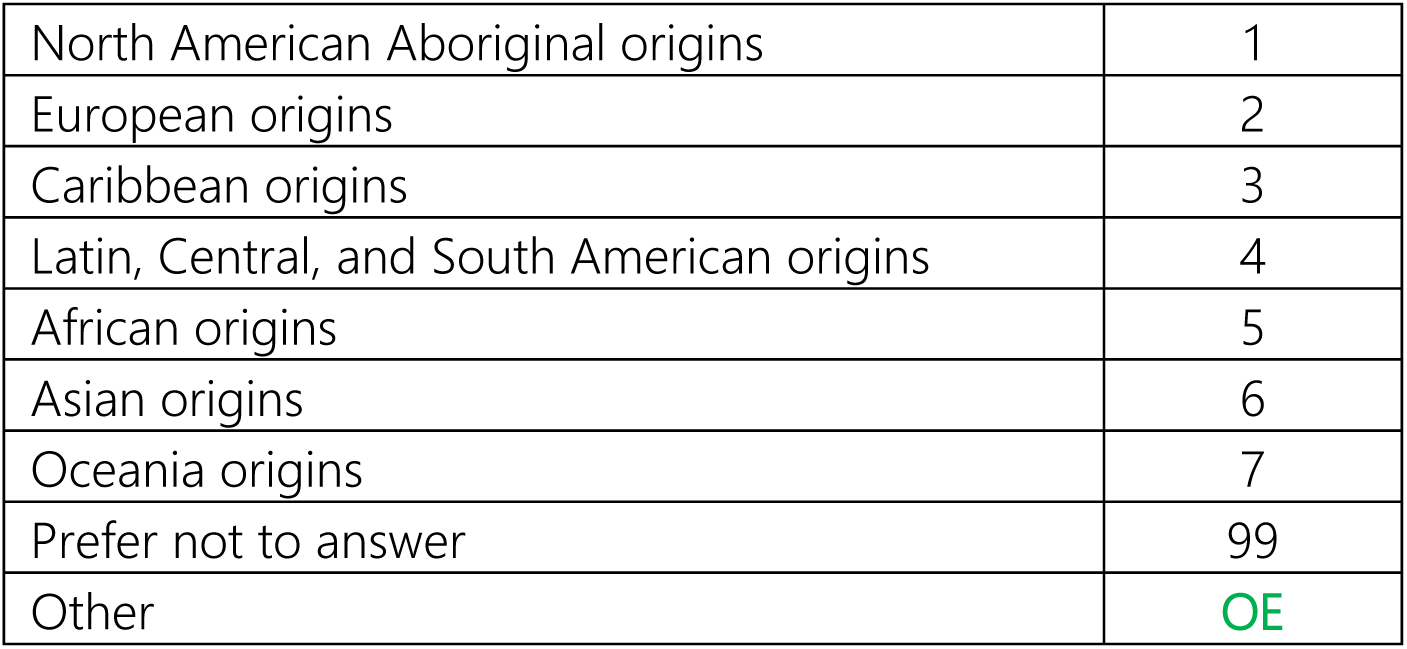 General information This information will help us understand whether people of different backgrounds may have different views. Your responses will be kept strictly anonymous and used only for research purposes.
16. How would you self-identify with respect to ethnicity and/or ancestry?

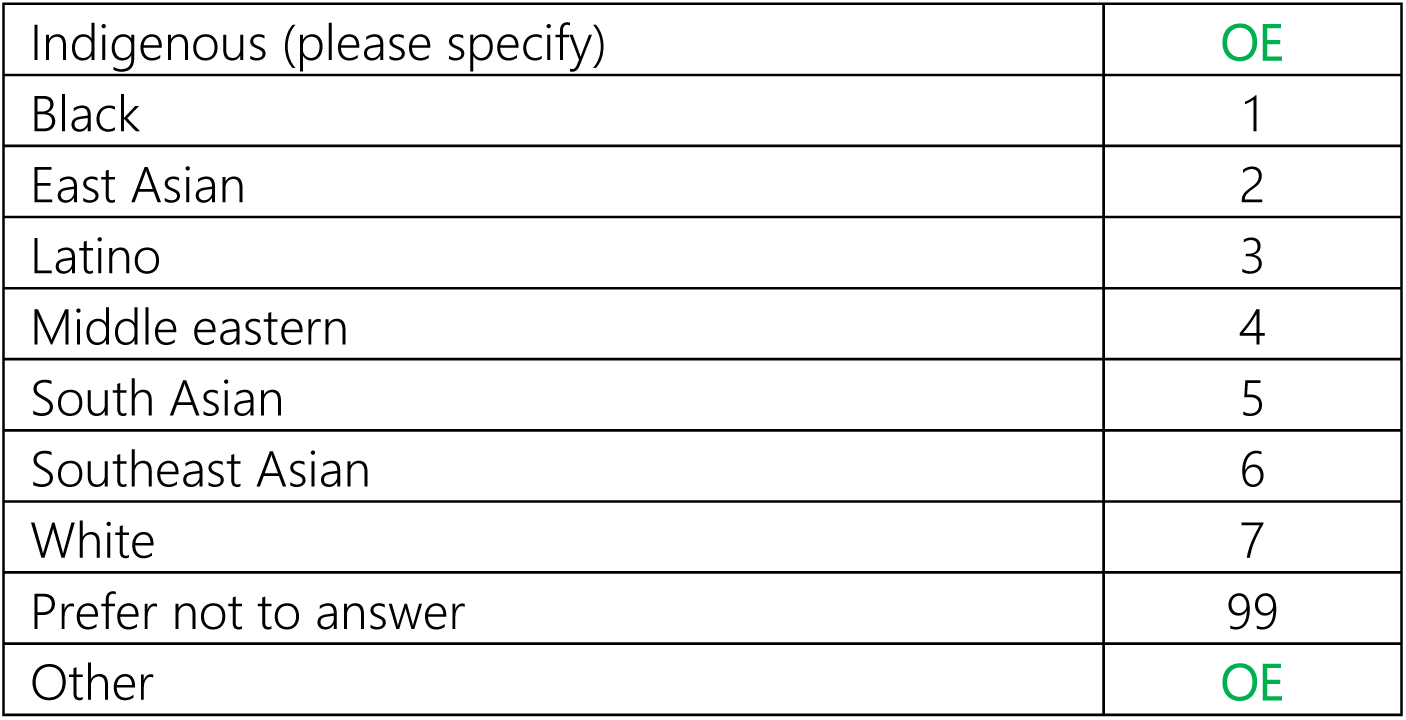
17. How do you self-identify with respect to race?

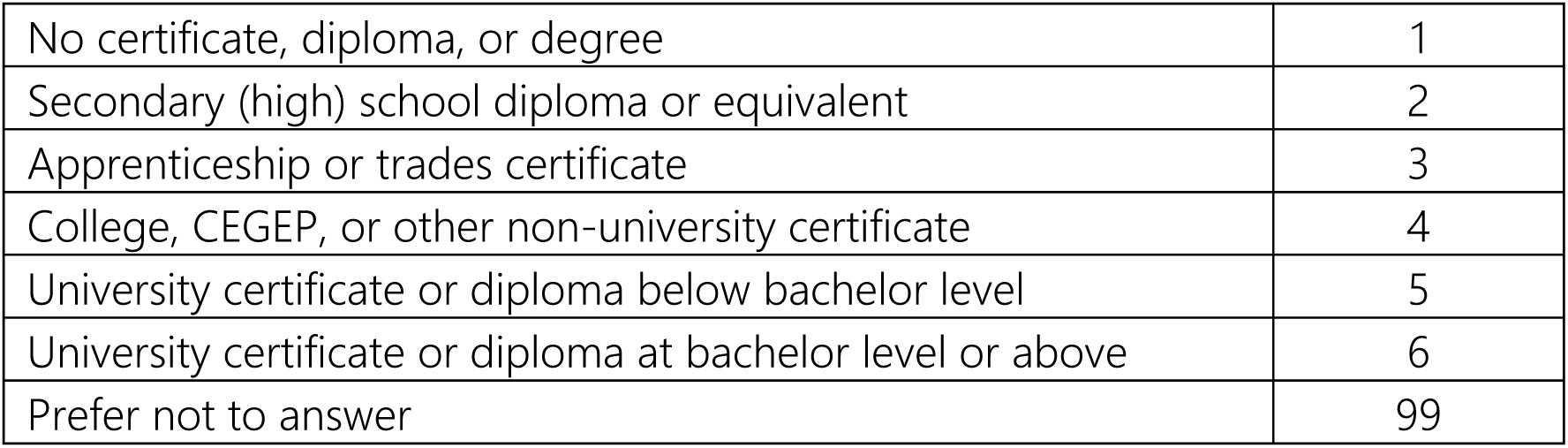
18. What is your highest level of education attainment?

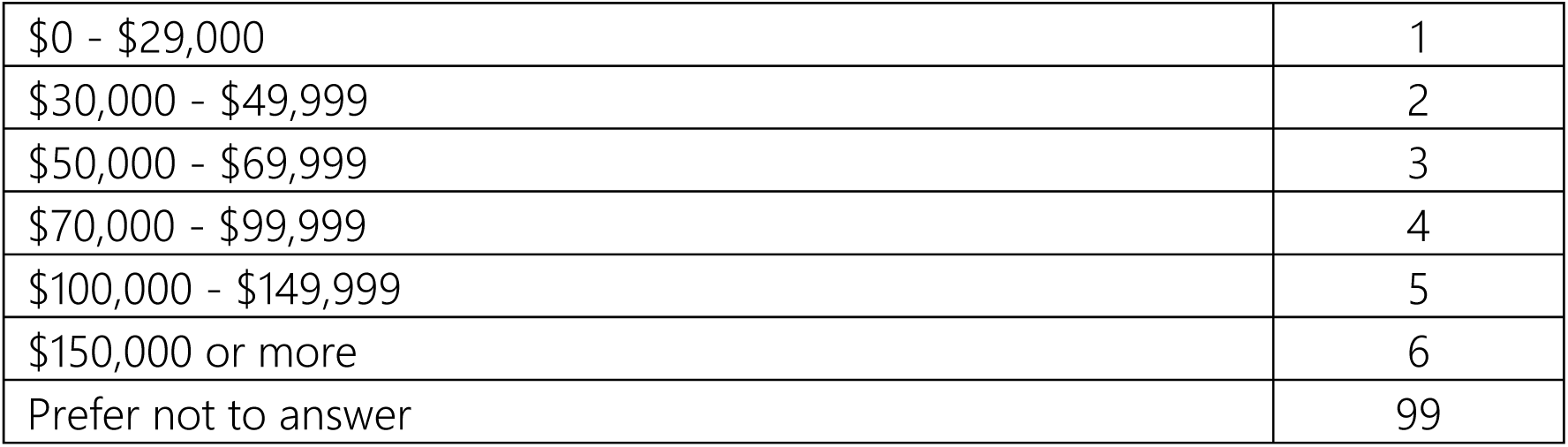
19. What was your total household income before taxes last year?

20. Including yourself, how many family members live in your household?

20. **Including yourself, how many family members live in your household?**
|  |  |
| --- | --- |
| OE |  |
| Prefer not to answer | 99 |

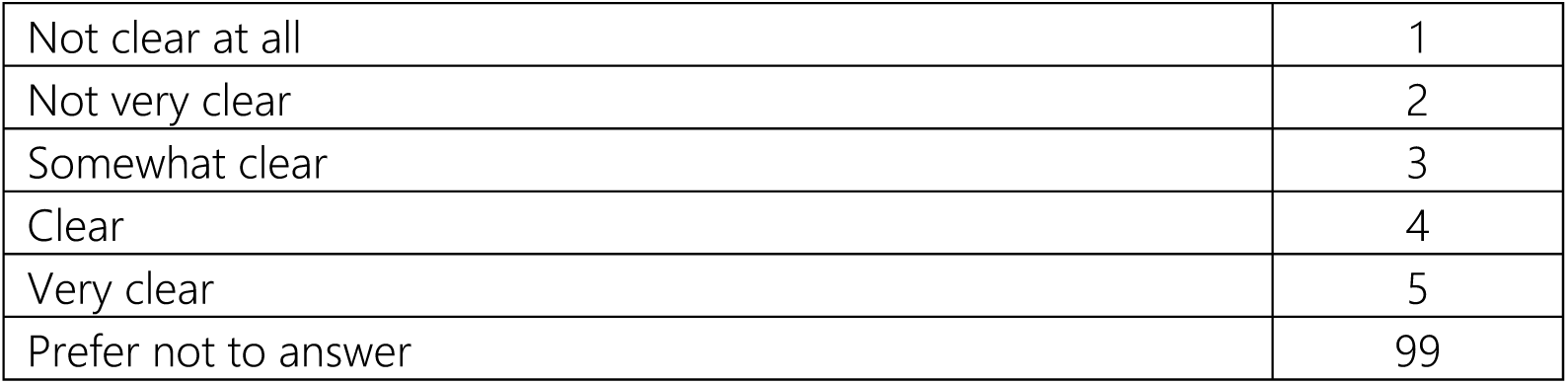 Final comments Please answer the following questions
21. How clear were the instructions to complete this survey?

22. Please use this space to make any final comments.

22. Please use this space to make any final comments.
|  |  |
| --- | --- |
| Prefer not to answer | 99 |
| OE |  |

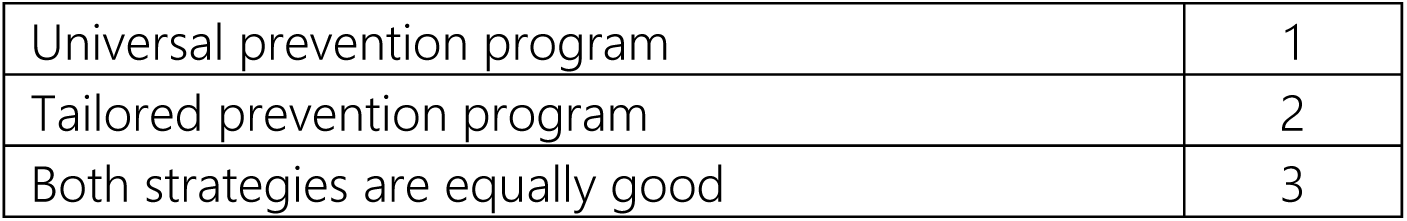

## Appendix 5: Generic survey

### Understanding public views on funding the health system

This questionnaire should take about 10-15 minutes to complete.

Your responses will be kept strictly anonymous and used only for research purposes.

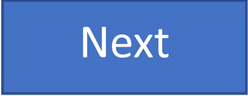

1. Your age?

|  |  |
| --- | --- |
| OE | TERMINATE if age<br>< 18 |
| Prefer not<br>to say | 99 |

[TERMINATE IF Q1=99]
2. Your gender?

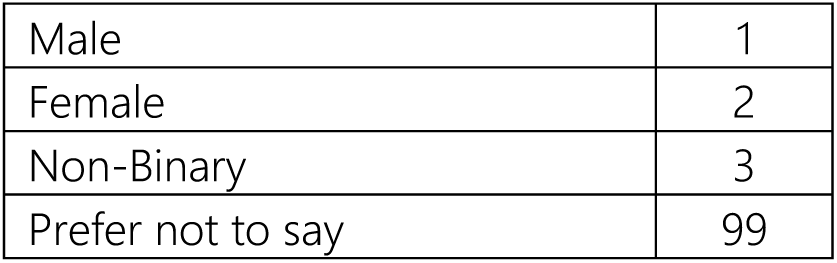

[TERMINATE IF Q2=99]
3. In which province/territory do you live?

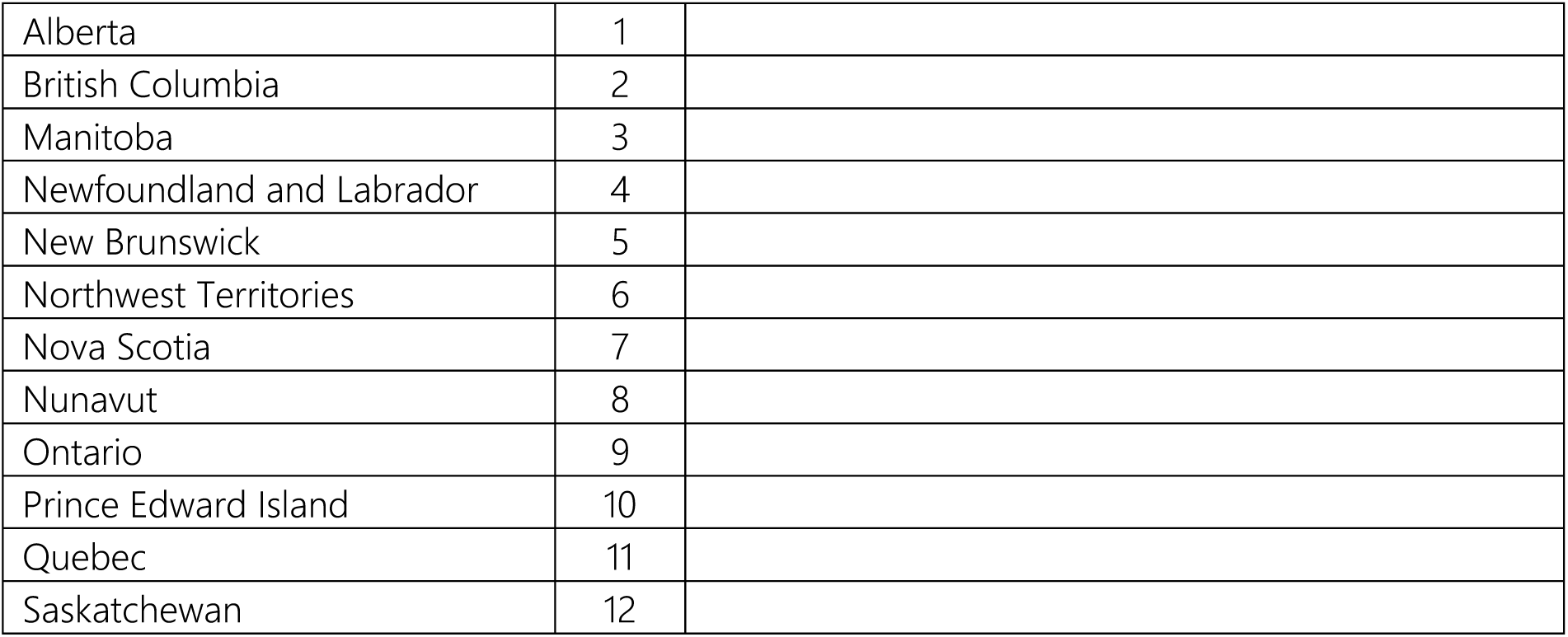
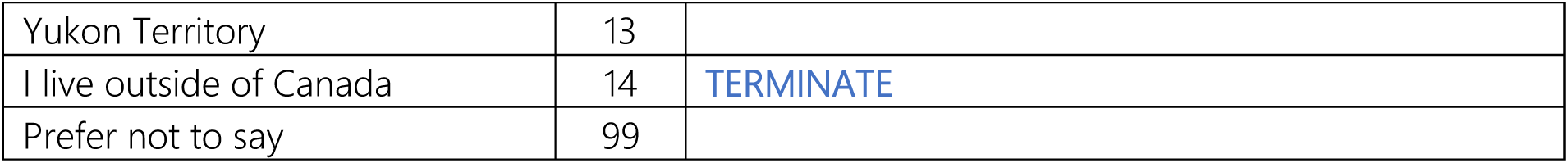

**3. In which province/territory do you live?**
|  |  |
| --- | --- |
| Alberta | 1 |
| British Columbia | 2 |
| Manitoba | 3 |
| Newfoundland and Labrador | 4 |
| New Brunswick | 5 |
| Northwest Territories | 6 |
| Nova Scotia | 7 |
| Nunavut | 8 |
| Ontario | 9 |
| Prince Edward Island | 10 |
| Quebec | 11 |
| Saskatchewan | 12 |

|  |  |  |
| --- | --- | --- |
| Yukon Territory | 13 |  |
| I live outside of Canada | 14 | TERMINATE |
| Prefer not to say | 99 |  |

[TERMINATE IF Q3=99]
4. What are the first three characters of your postal code? (Format A1A)

**4. What are the first three characters of your postal code? (Format A1A)**
|  |  |
| --- | --- |
| OE |  |
| Prefer not to say | 99 |

[TERMINATE IF Q4=99] In the next few screens, you will be provided with an introduction to this survey, with some information for you to review. You will then be asked some questions based on this information. Introduction There are differences between populations with the **highest and lowest household incomes** in Canada in terms of how long people live, their quality of life, and their access to healthcare. If we divide the population into five groups of equal size (each group = 20% of the population), then in 2019:
  - The highest-earning 20% of the population had an average household income after tax of **$160,000** per year.
  - The lowest-earning 20% of the population had an average household income after tax of **$25,000** per year.

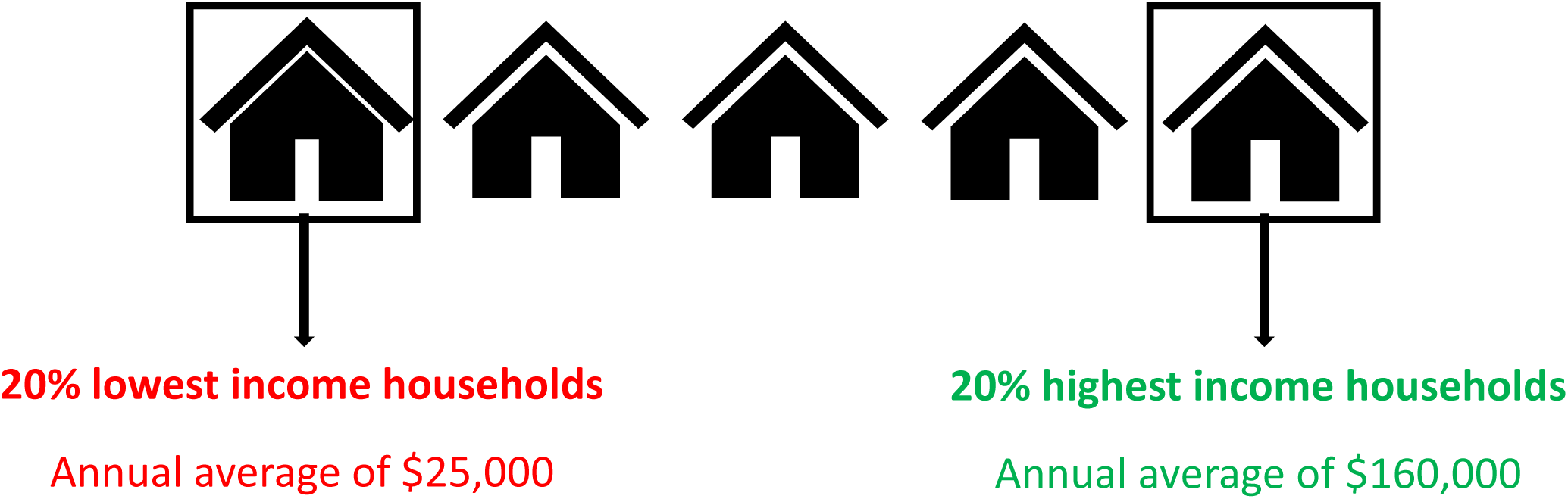
5. While actual length of life and health vary between individuals, on average, the **20% highest household income population** experience **79 years** of life in full health.
6. On the other hand, the **20% lowest household income population** experience **68 years** of life in full health. For example, someone who has 79 years in full health might have lived to 85 years old, but with less than full health. These are averages across the population of Canada. Each individual’s actual length of life and health can vary considerably from these averages.

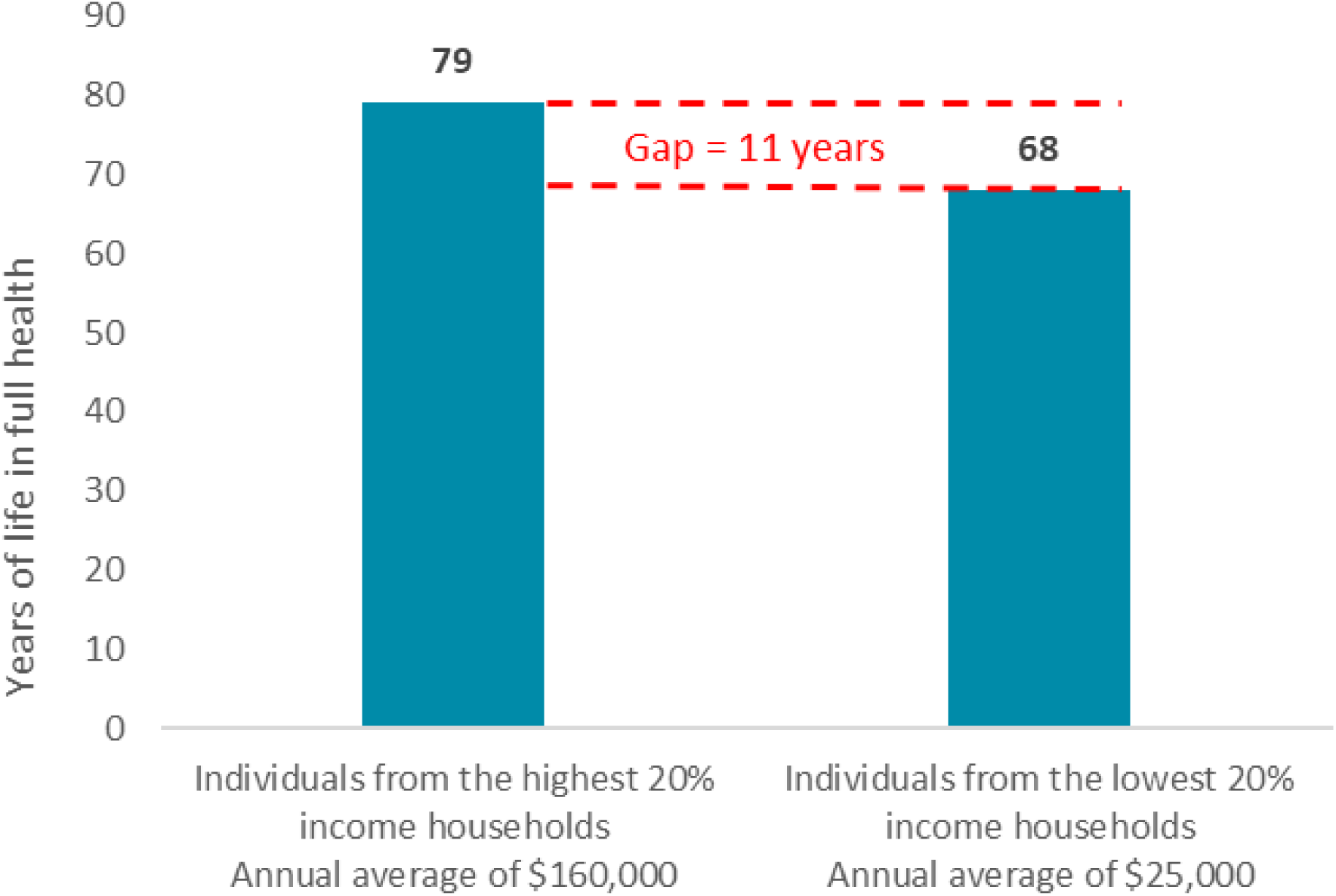

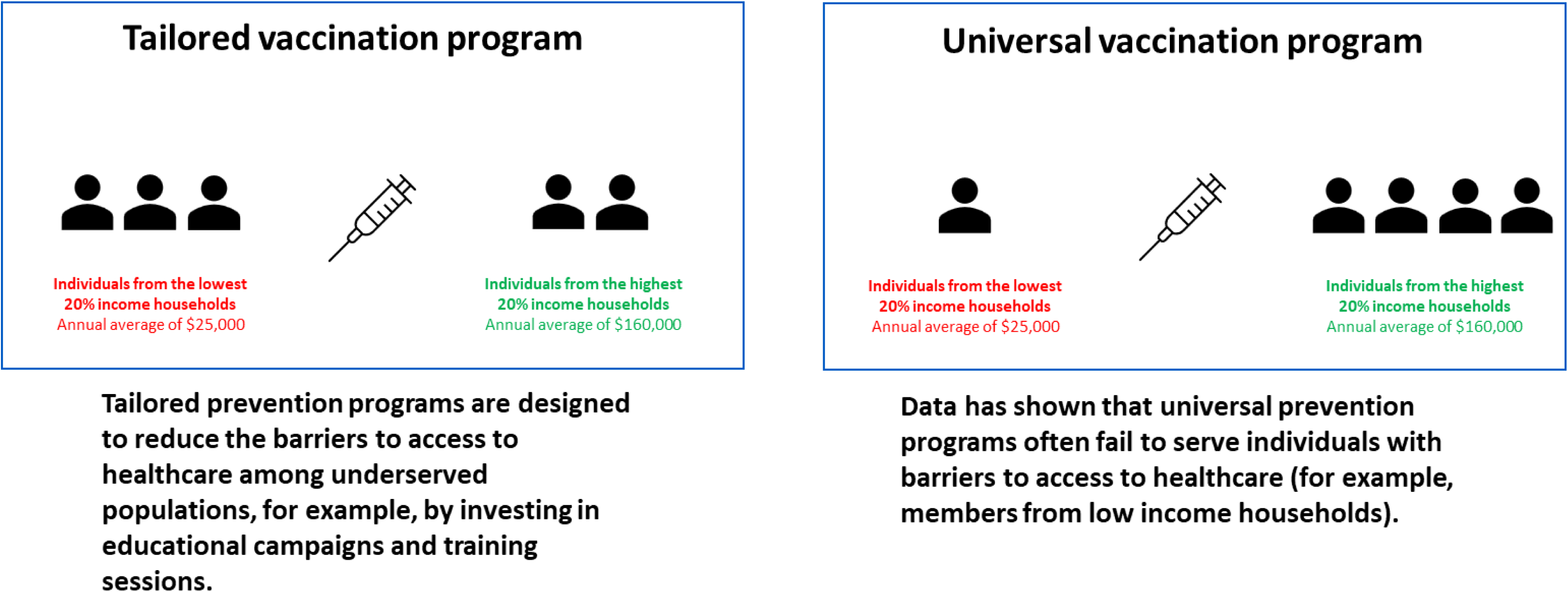

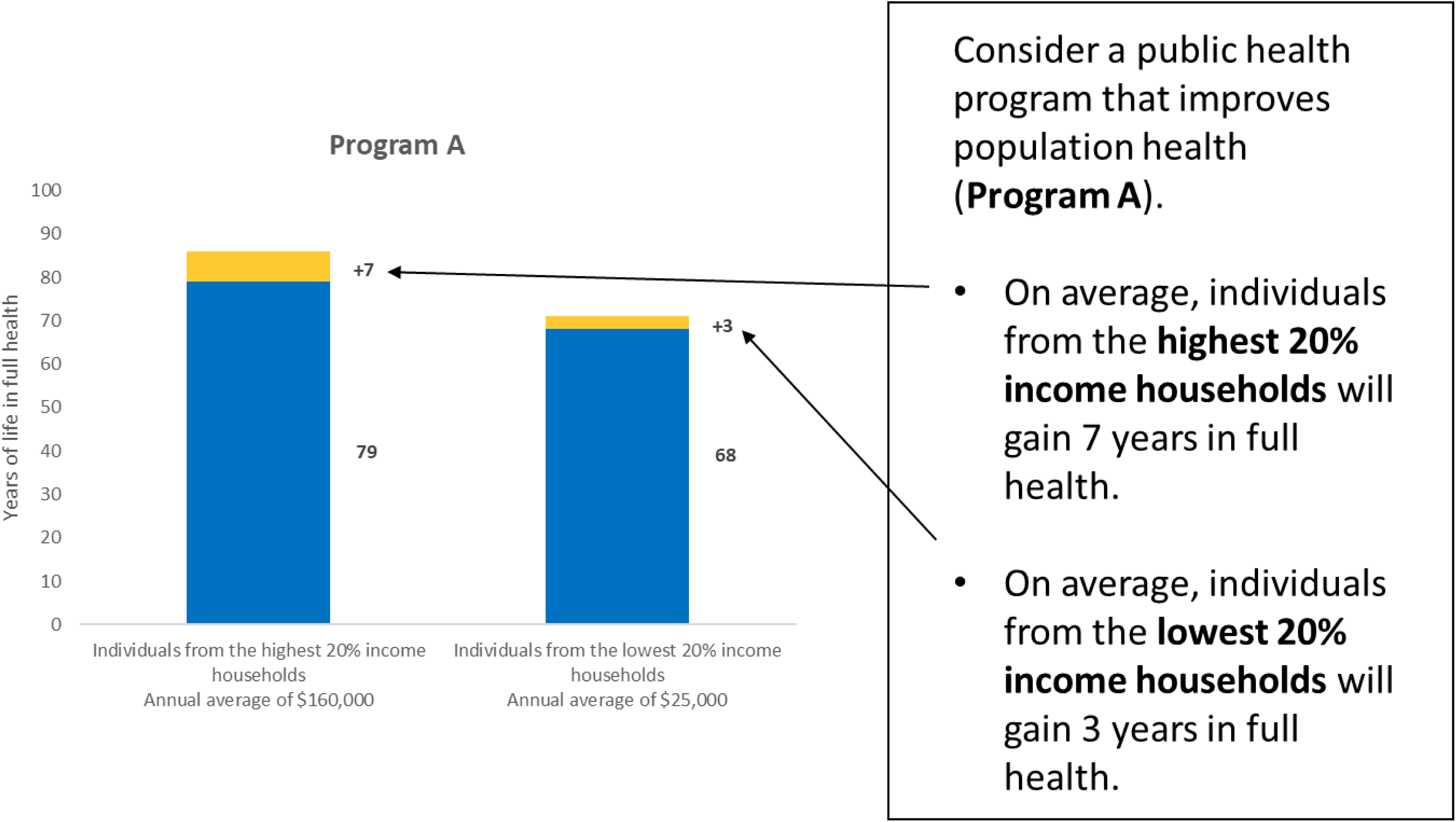

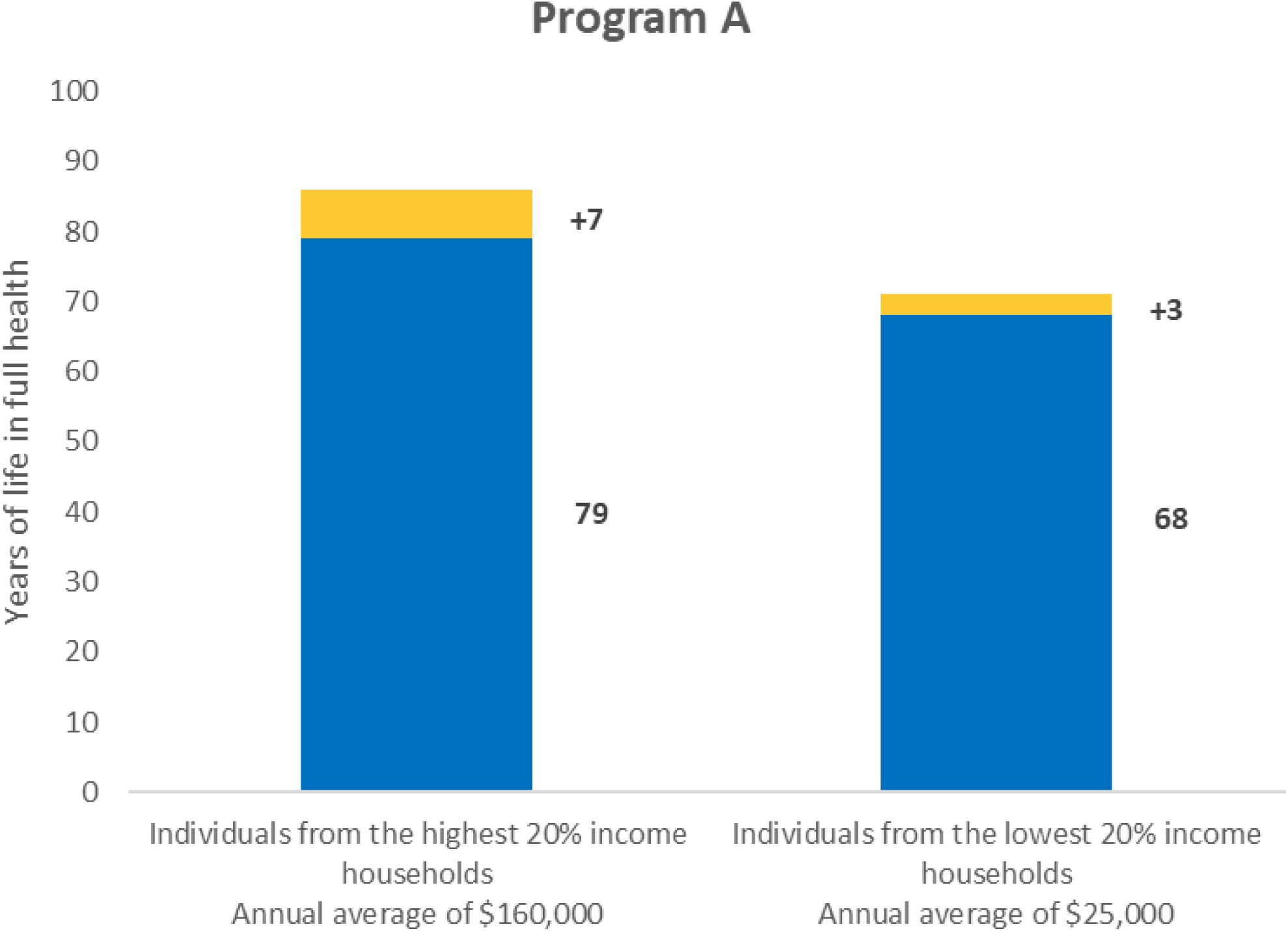
7. Which population group has the highest life expectancy AFTER implementing Program A?

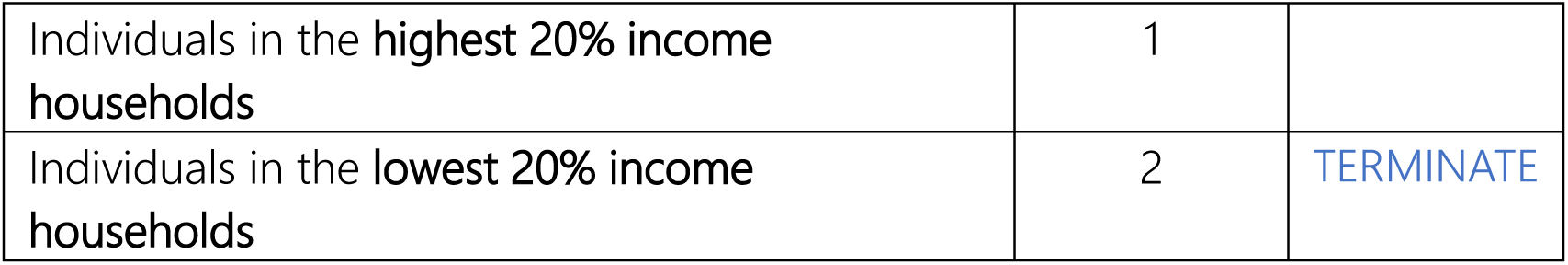

|  |  |  |
| --- | --- | --- |
| Individuals in the highest 20% income households | 1 |  |
| Individuals in the lowest 20% income households | 2 | TERMINATE |

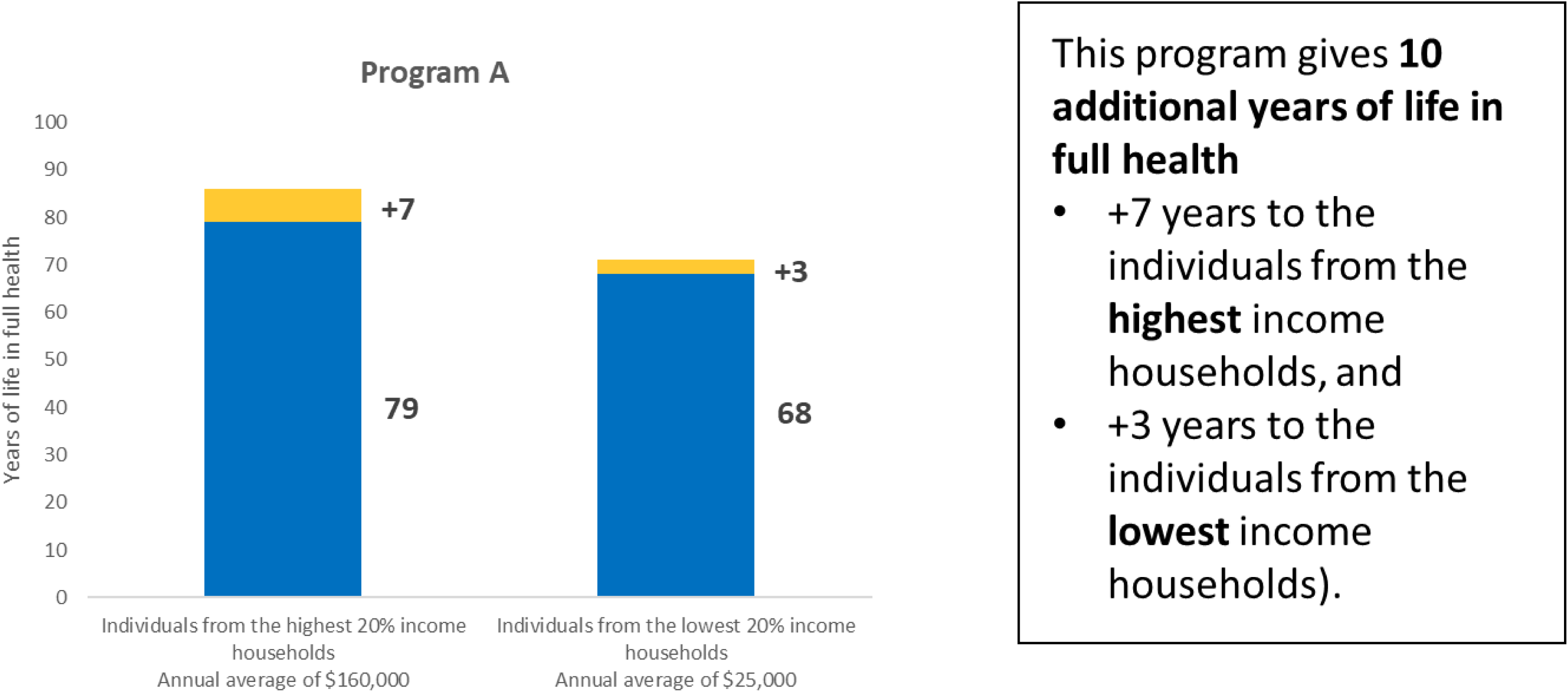

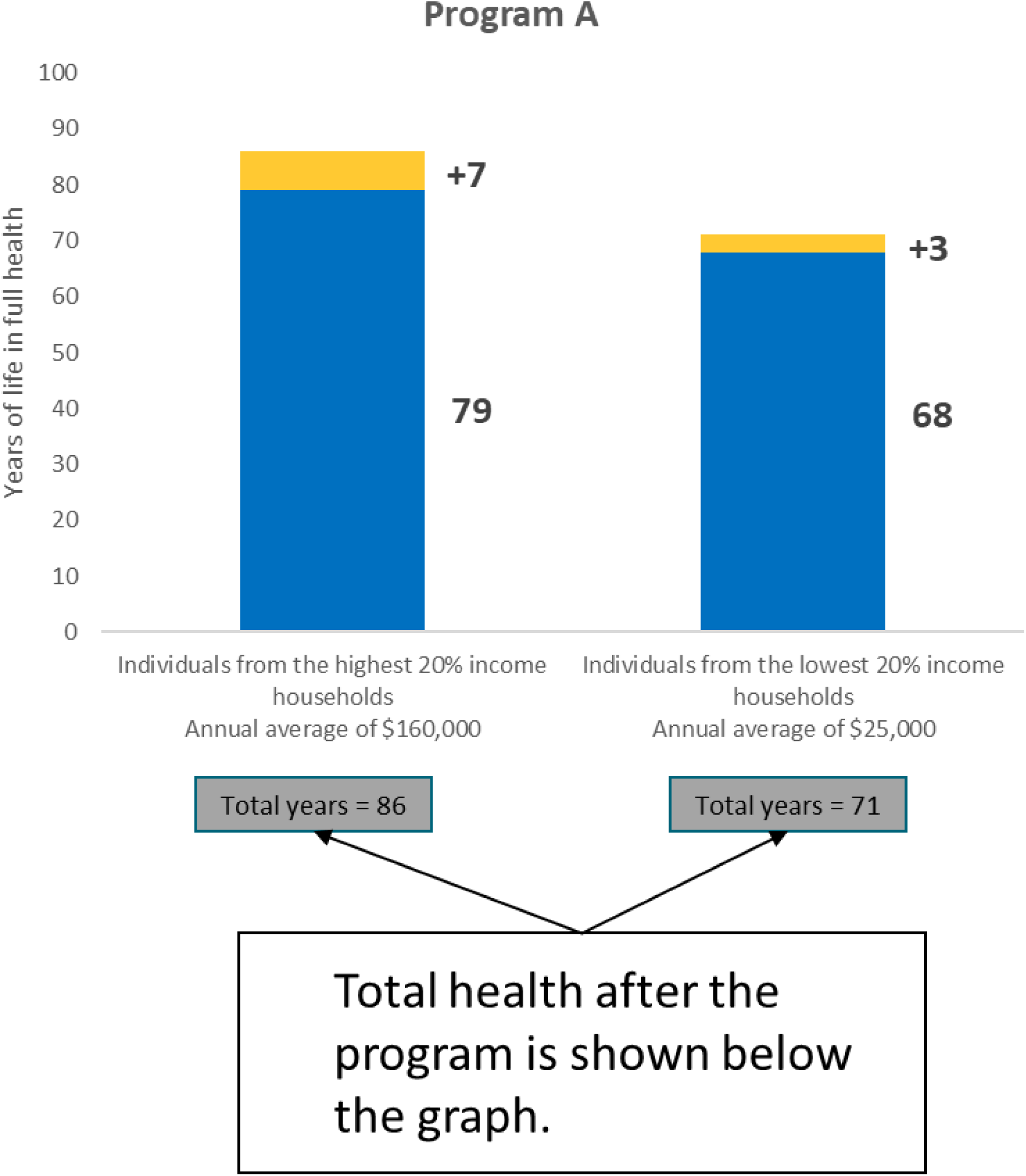

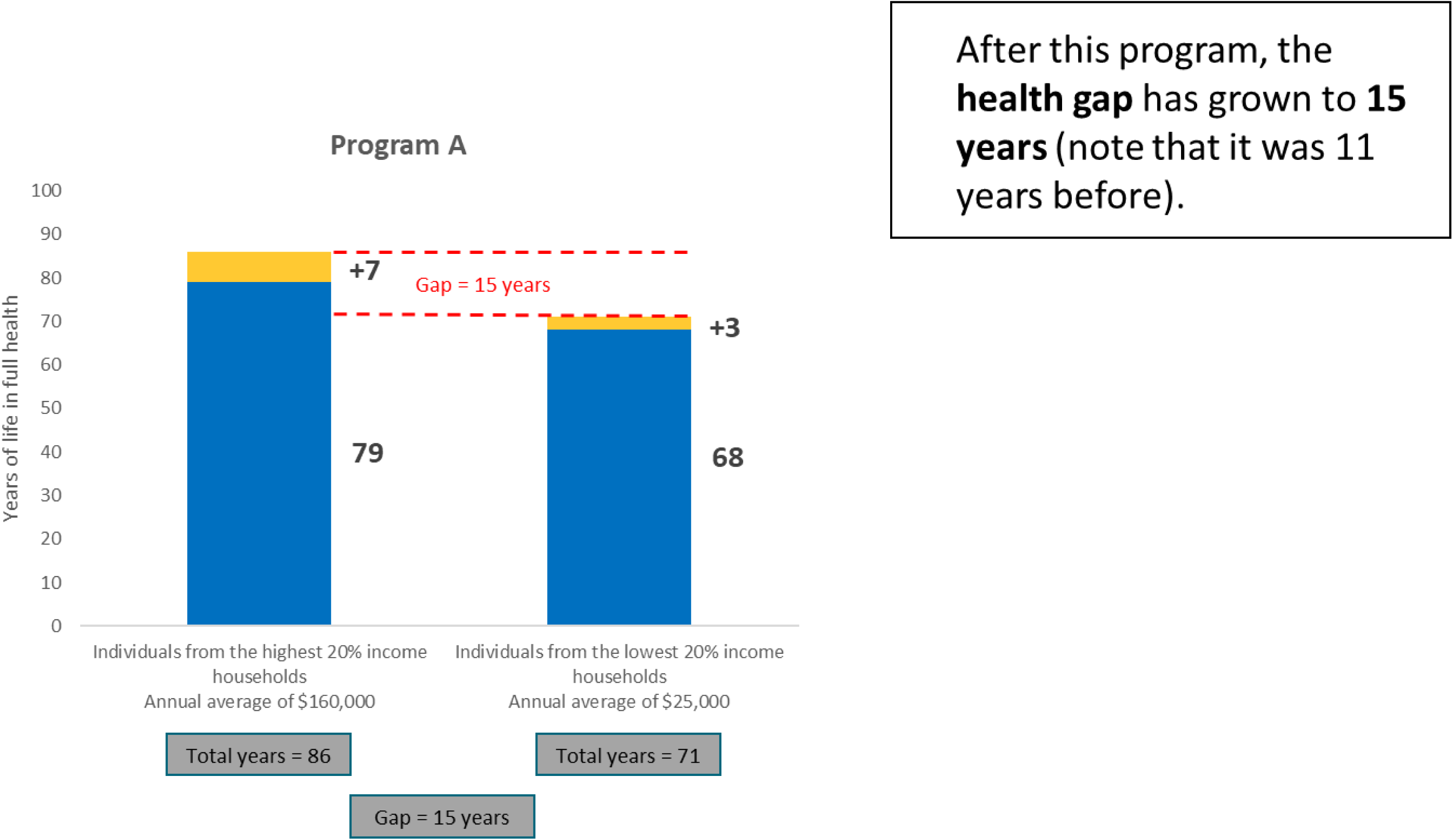

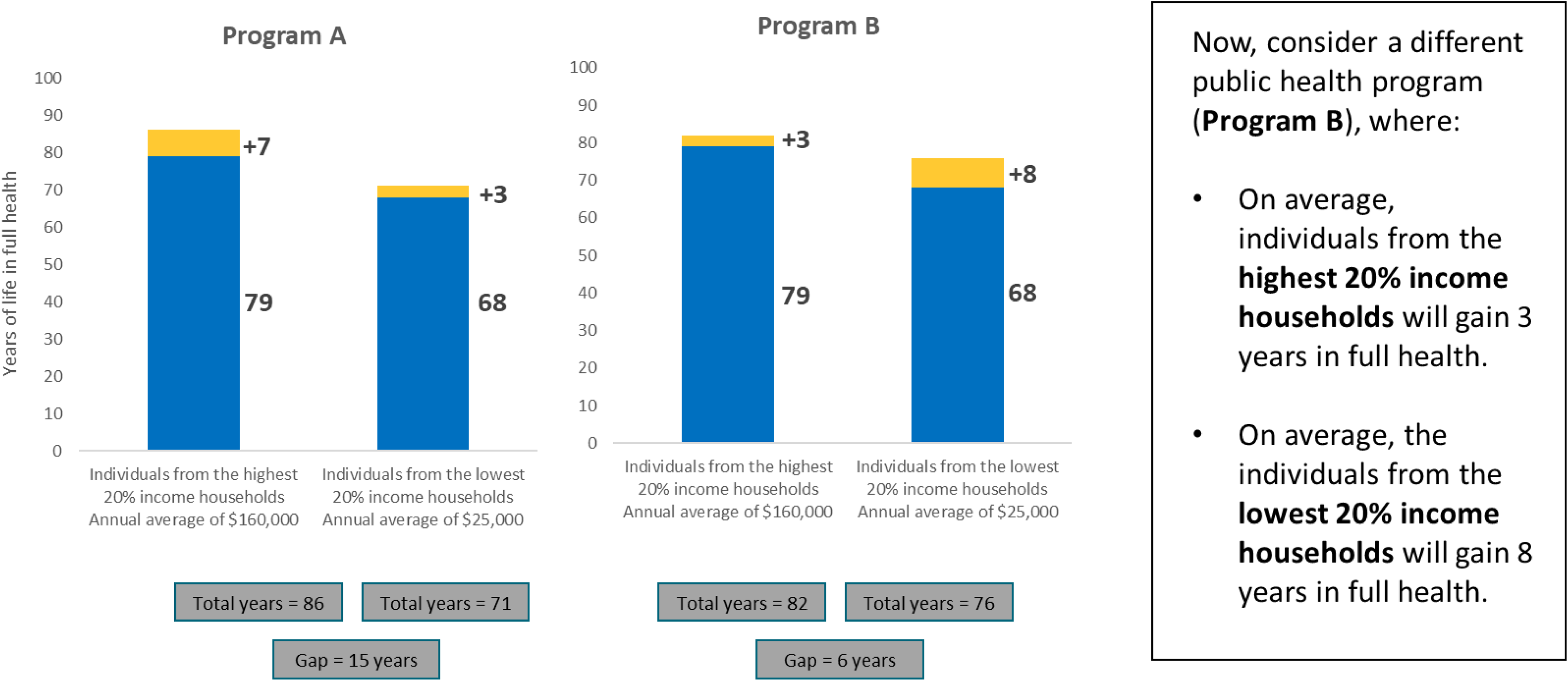

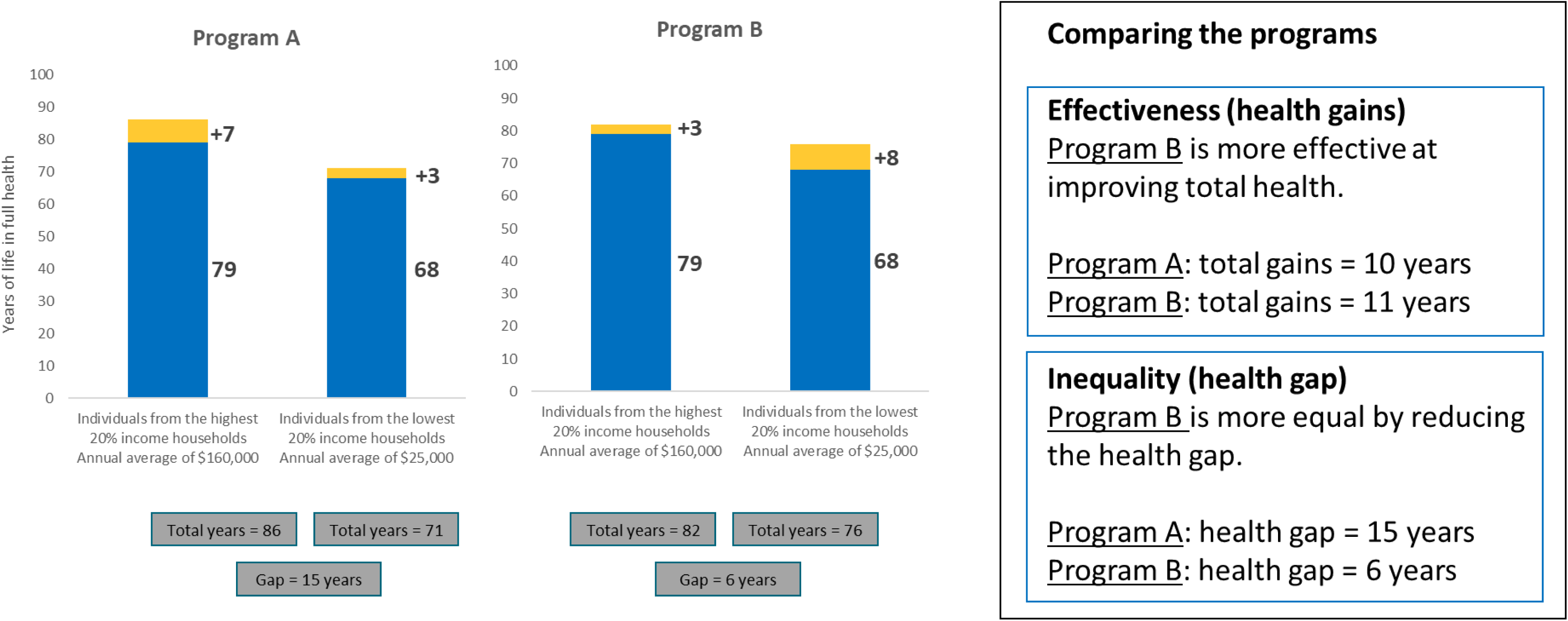

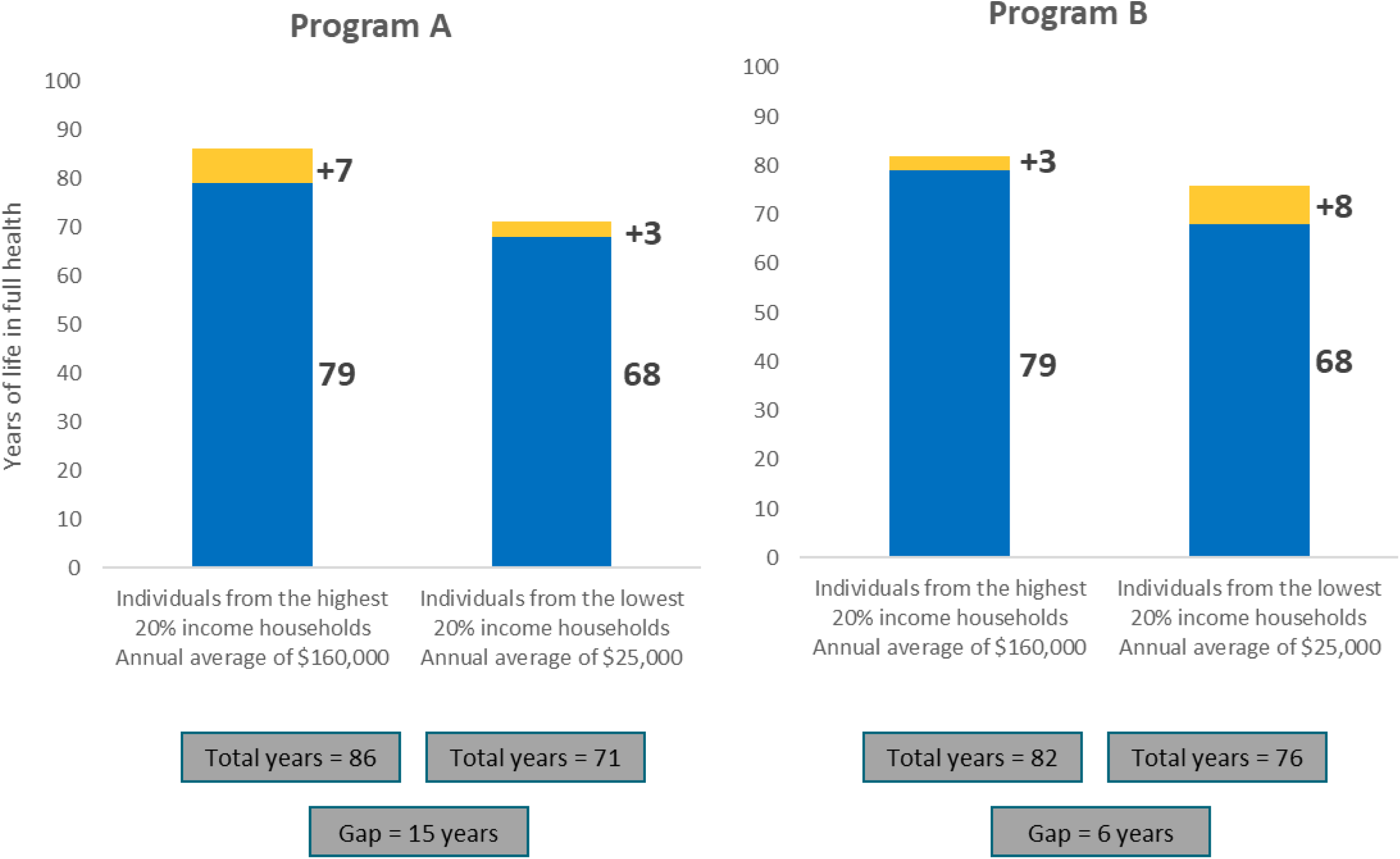
8. Which program would you choose?

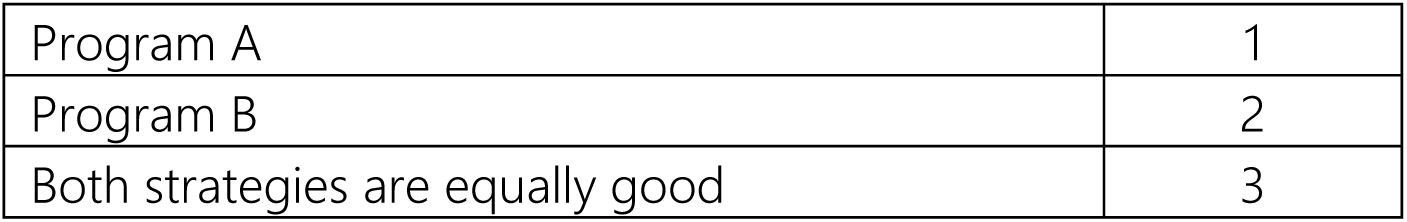 Survey questions Which program should the government choose? Answer the next 8 scenarios considering:
  - We cannot pay for both programs — a choice must be made.
  - “Either program is good” means you don’t mind which one is chosen.
  - Both programs cost exactly the same.
  - The only difference between the programs is how much health is gained by the individuals in the highest and lowest 20% income households.
  - The population in the middle is not affected.

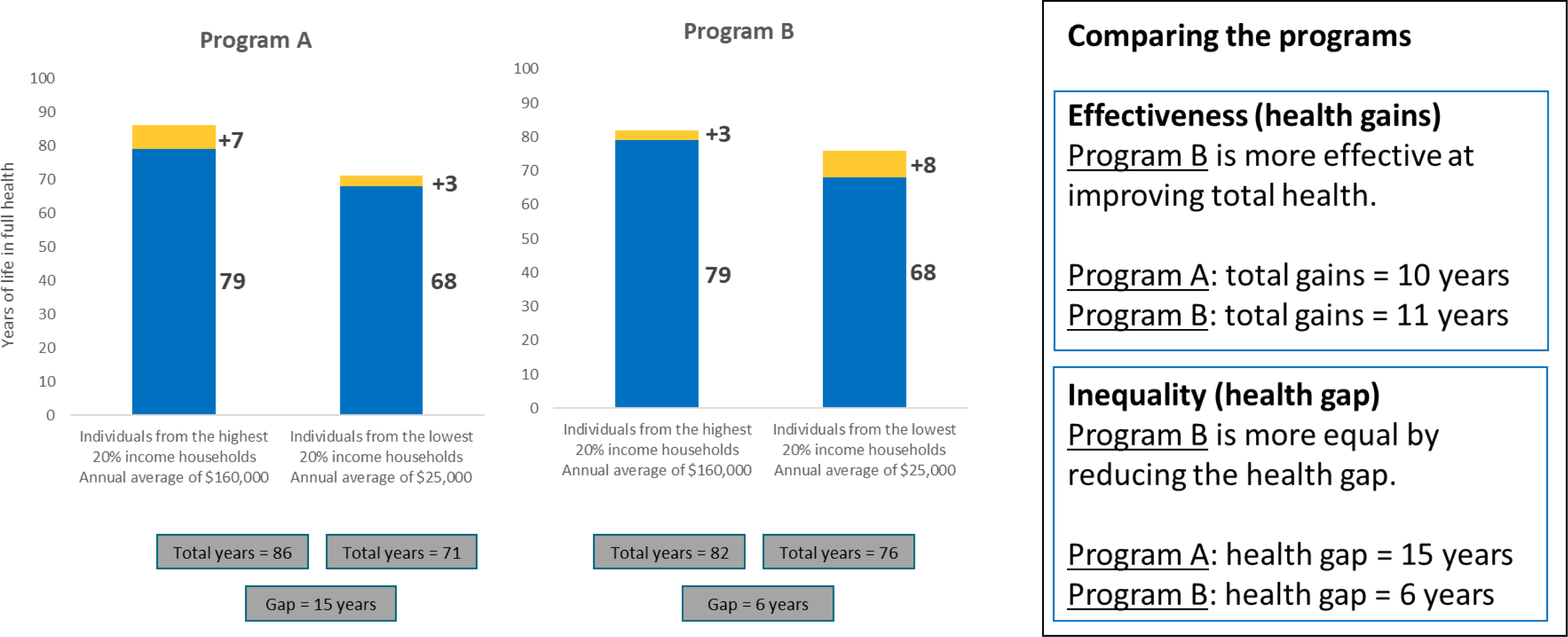
9. Which program would you choose?

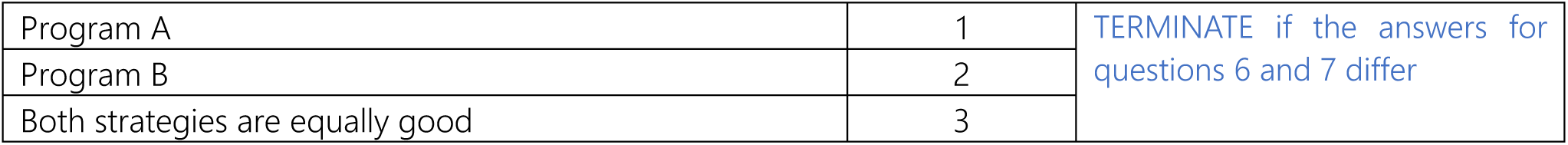

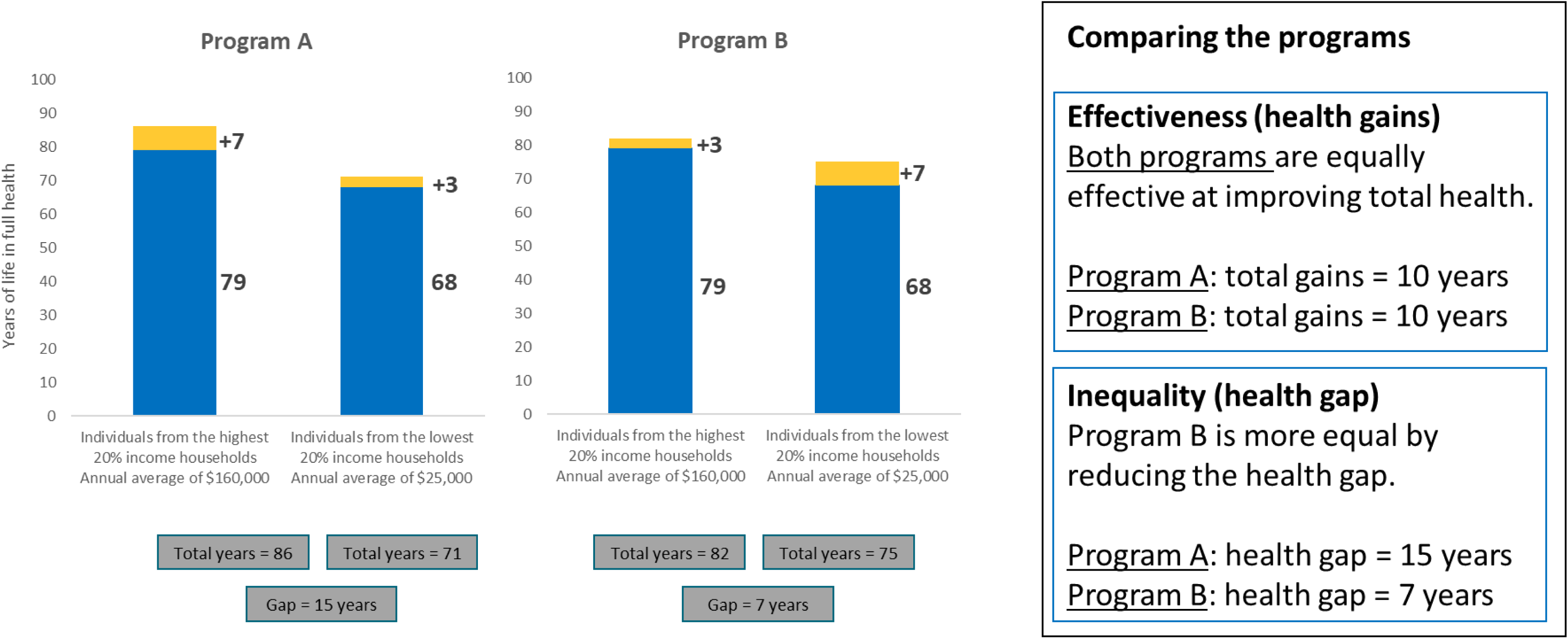
10. Which program would you choose?

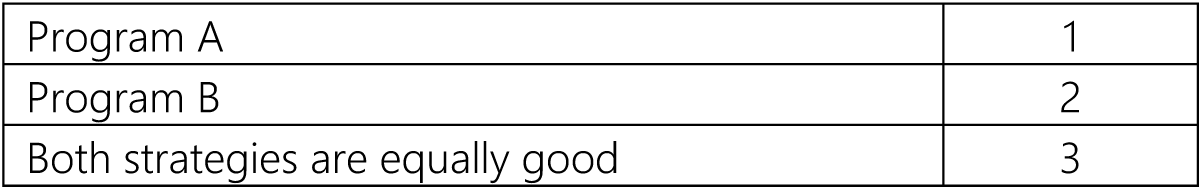

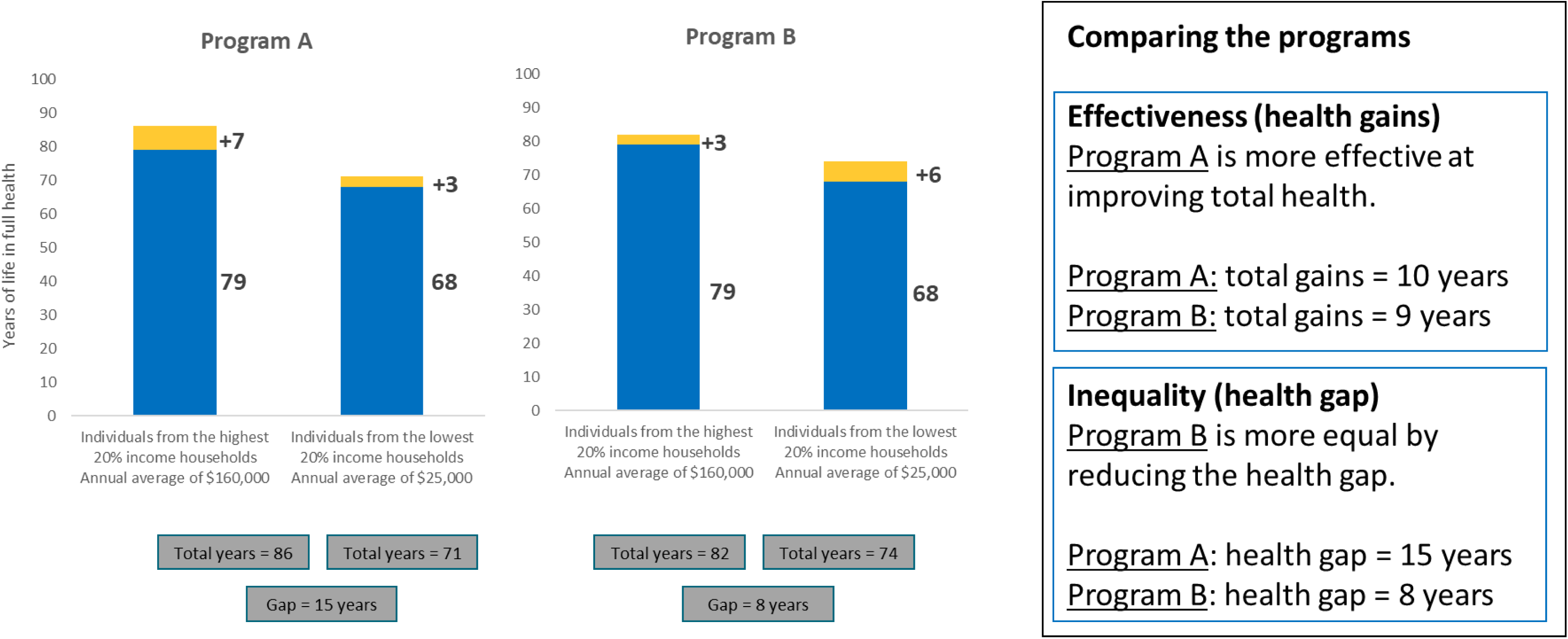
11. Which program would you choose?

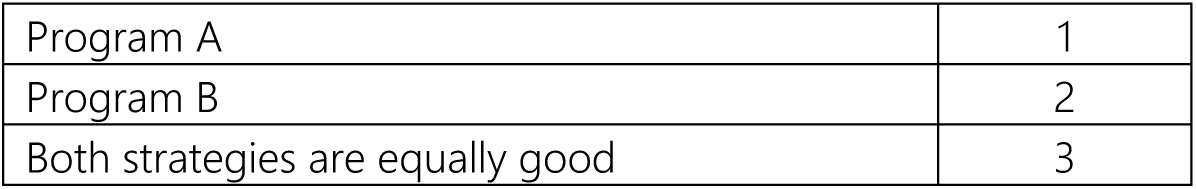

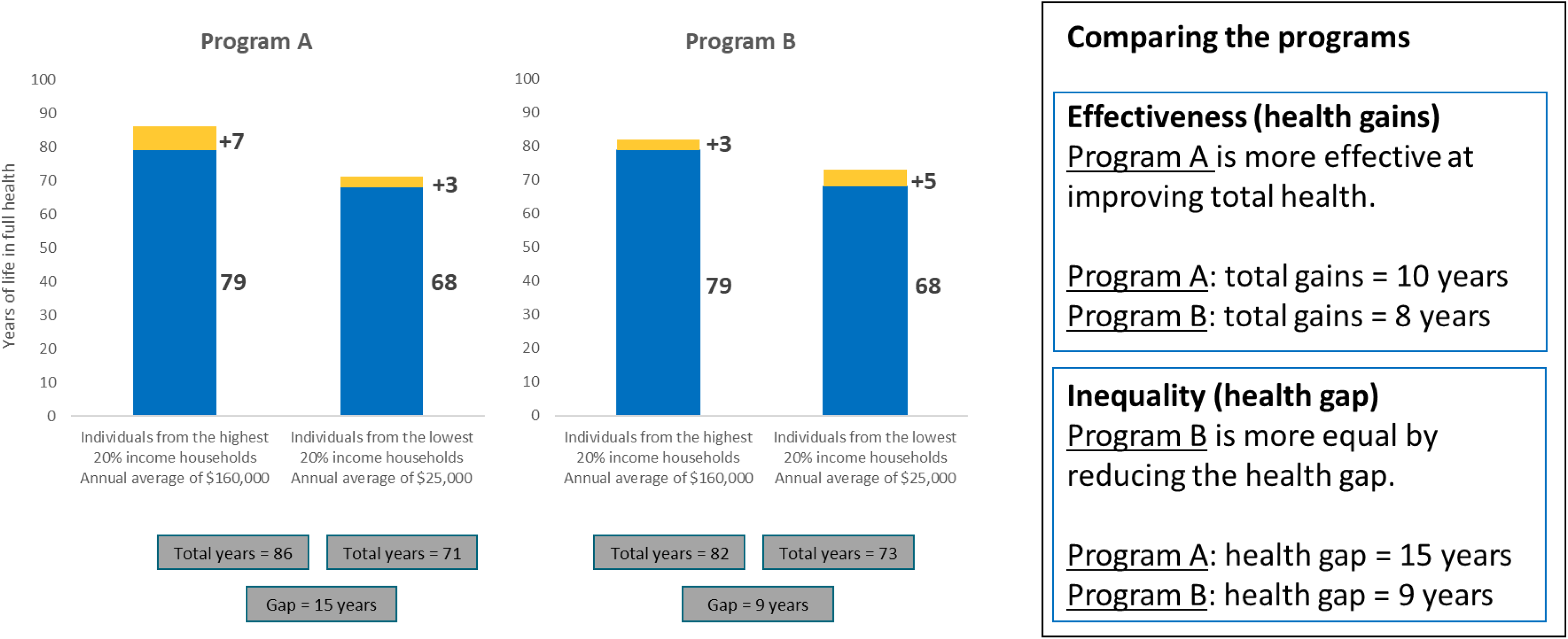
12. Which program would you choose?

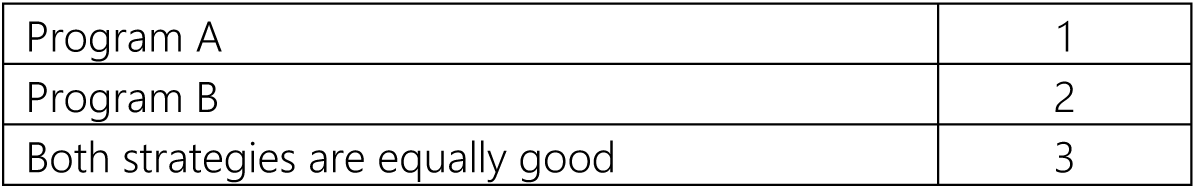

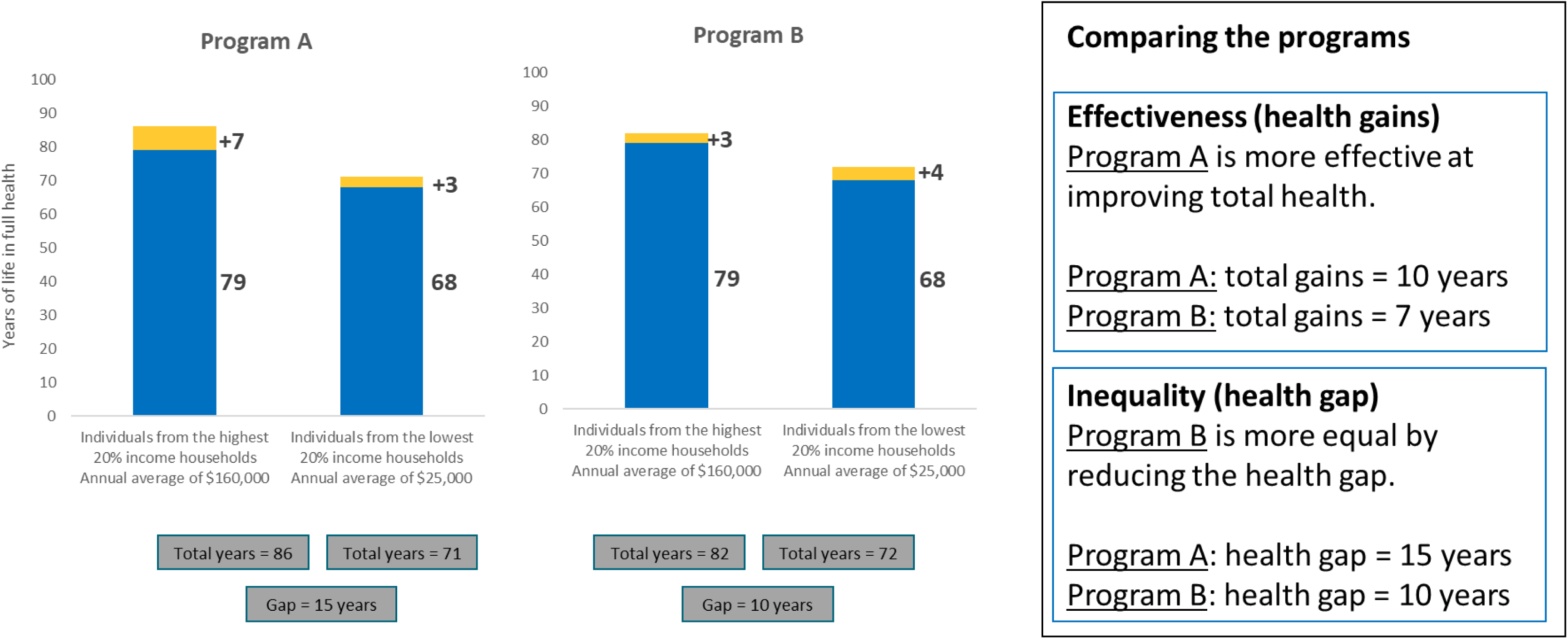
13. Which program would you choose?

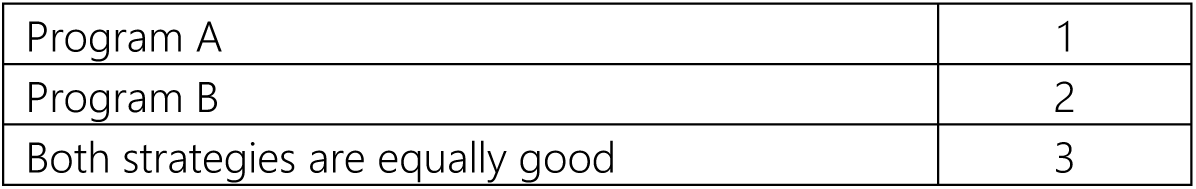

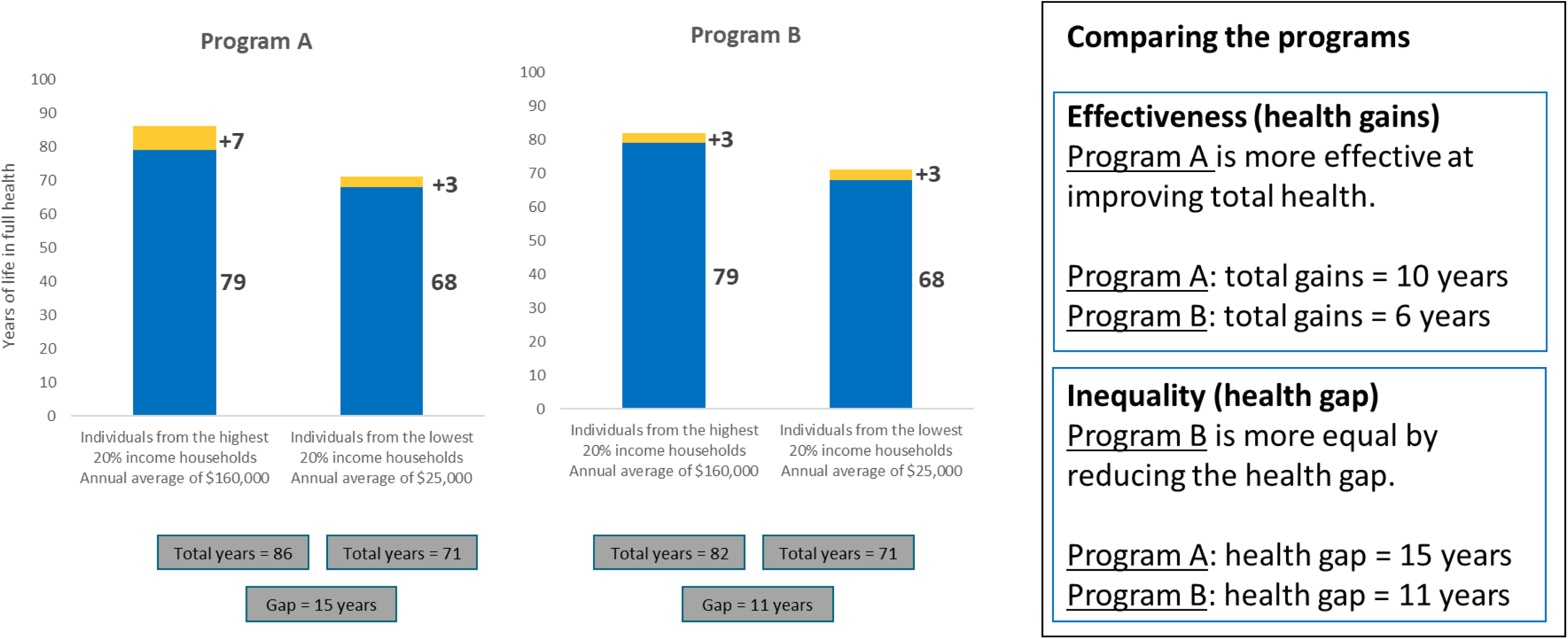
14. Which program would you choose?

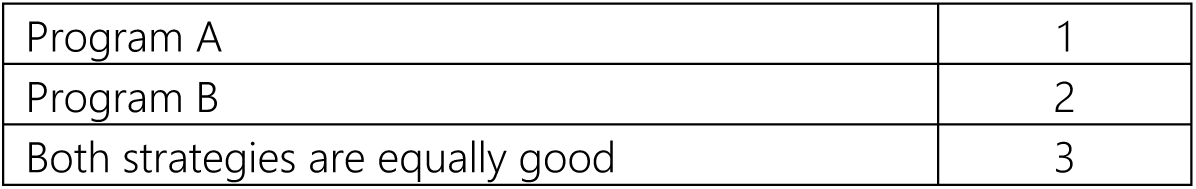

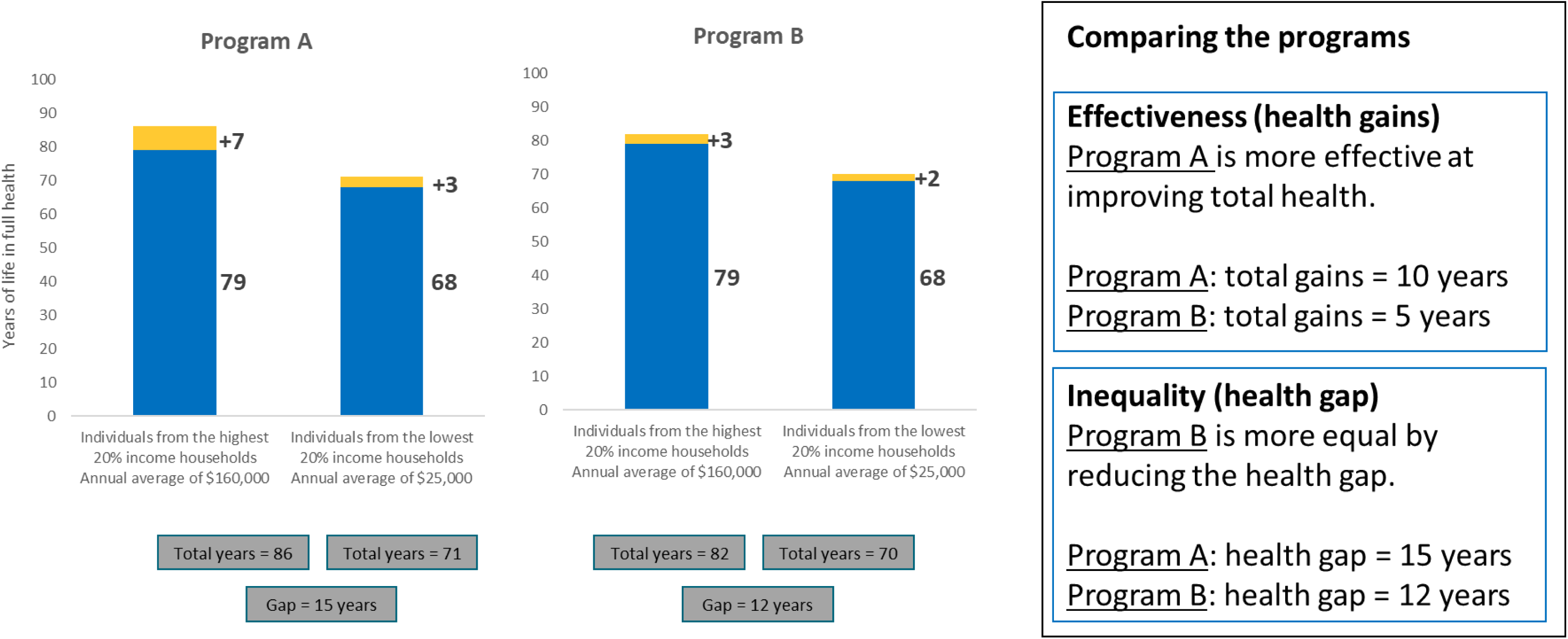
15. Which program would you choose?

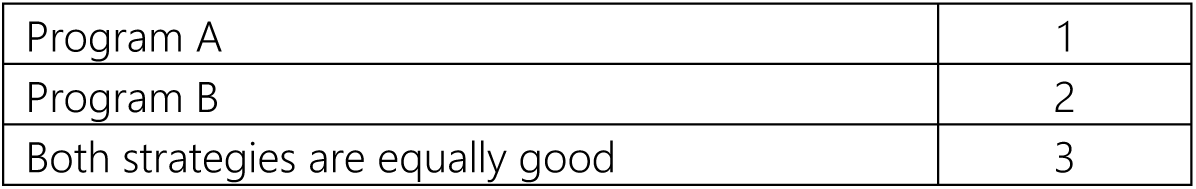

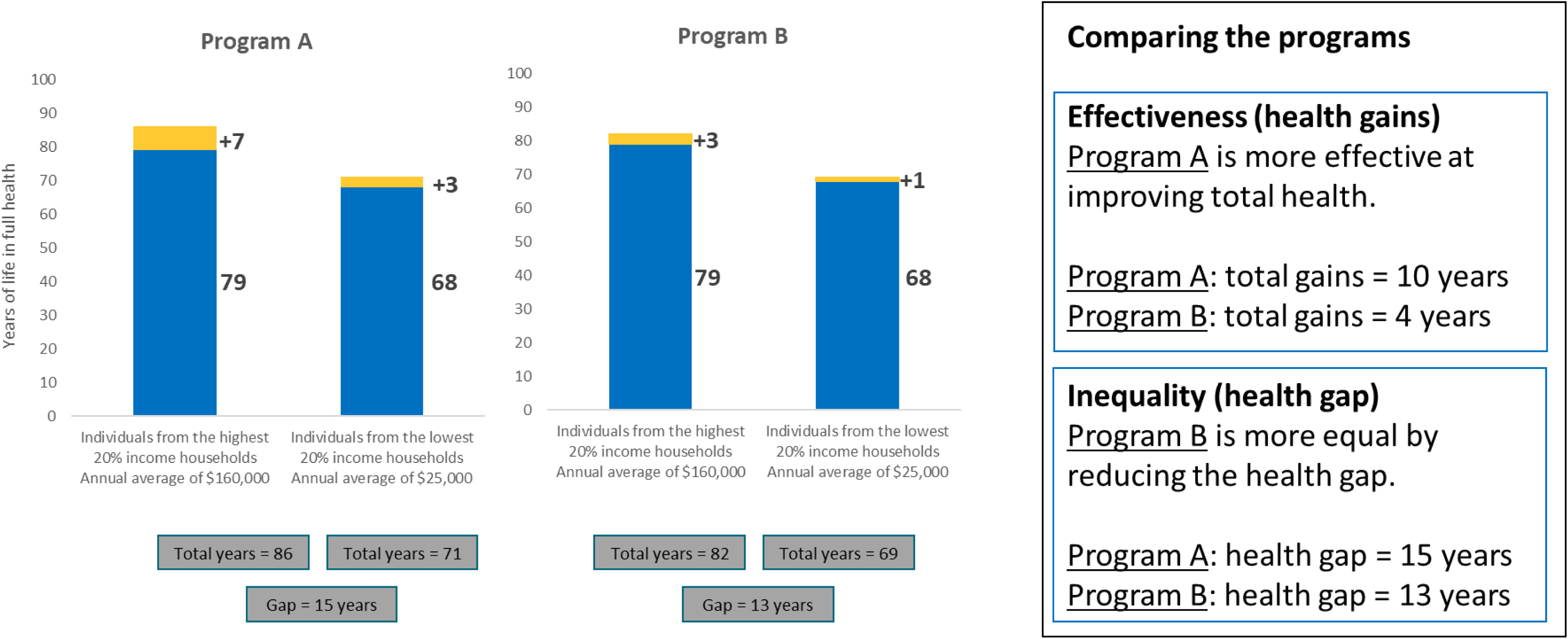
16. Which program would you choose?

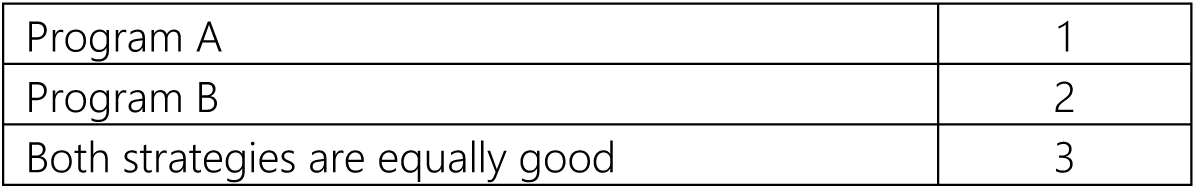 General information This information will help us understand whether people of different backgrounds may have different views. Your responses will be kept strictly anonymous and used only for research purposes.
17. How would you self-identify with respect to ethnicity and/or ancestry?

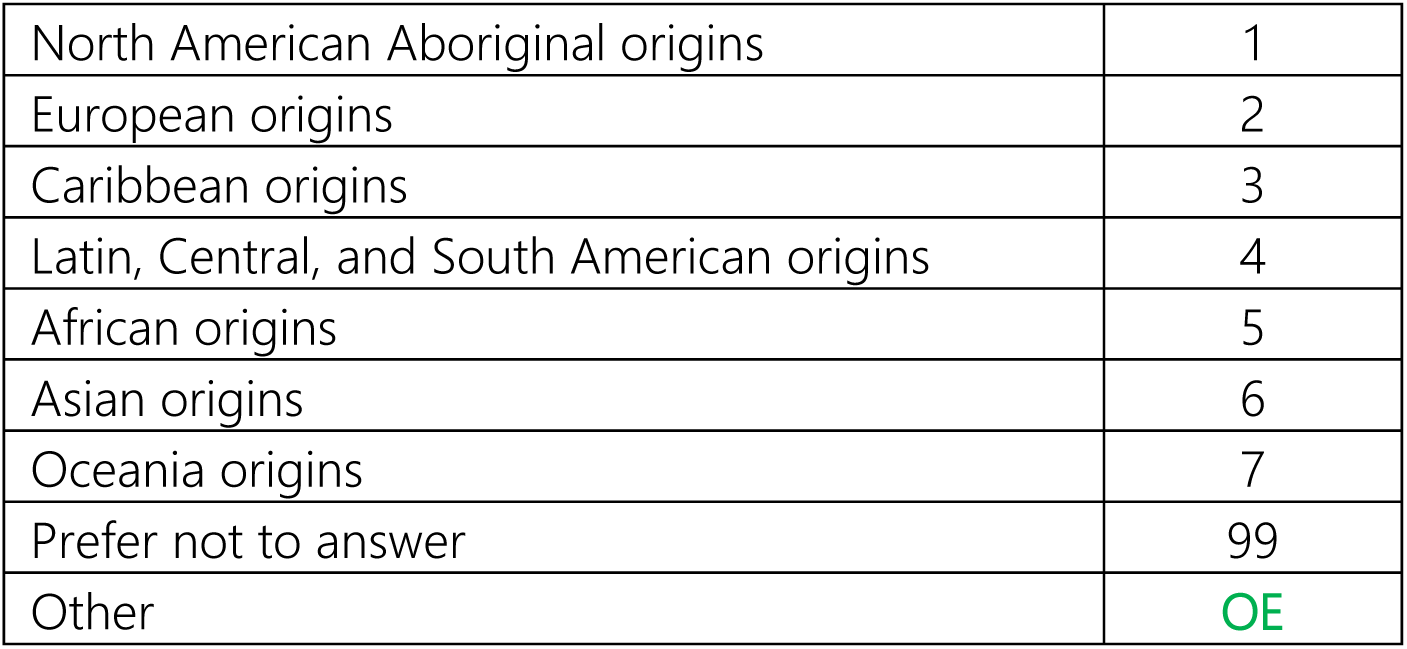
18. How do you self-identify with respect to race?

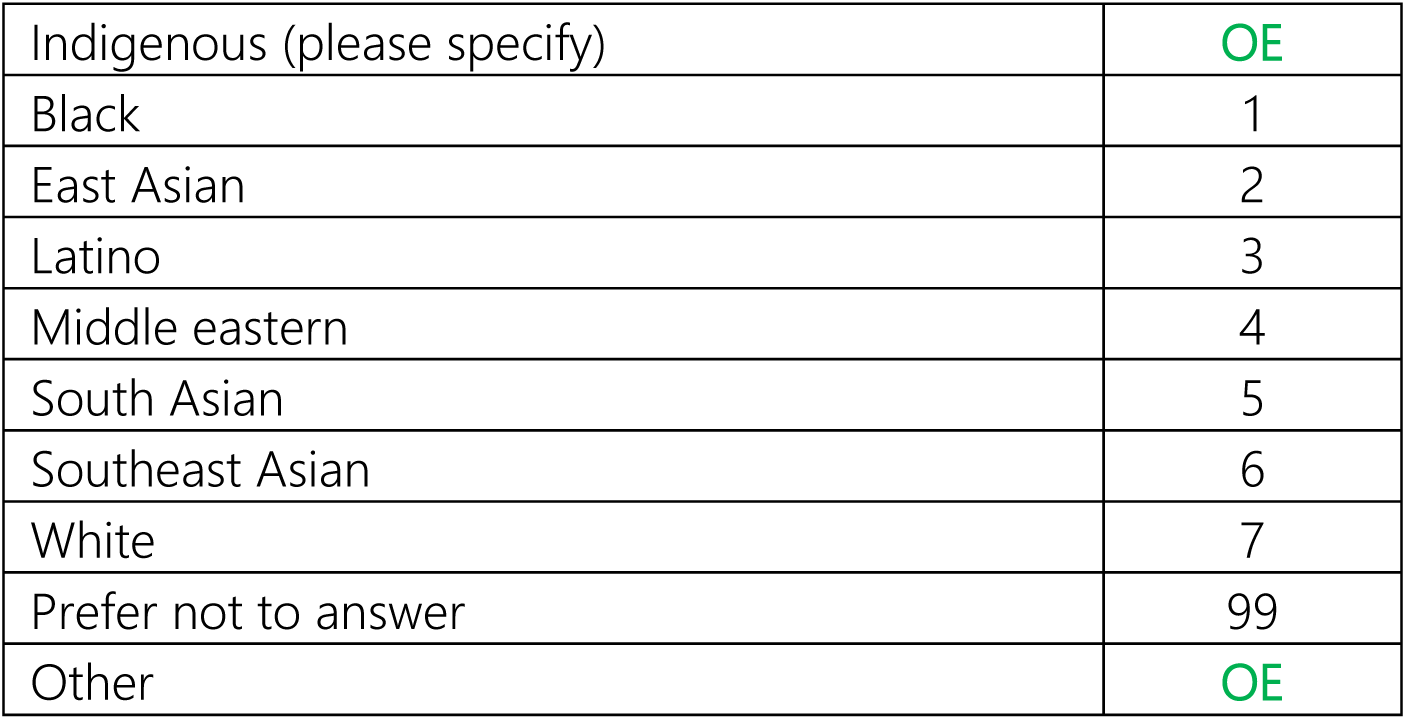
19. What is your highest level of education attainment?

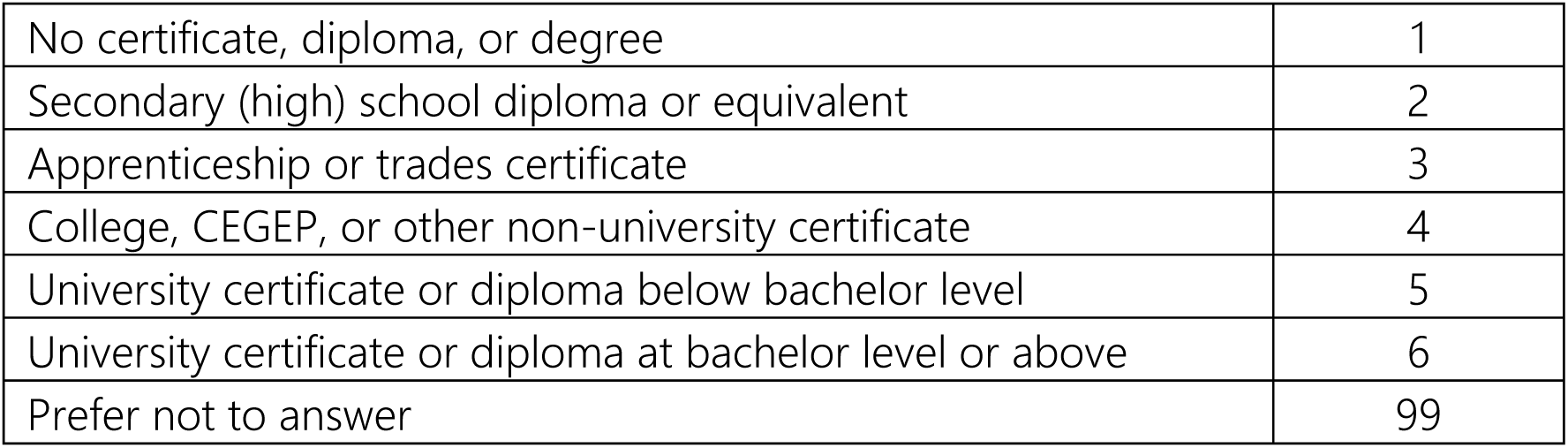
20. What was your total household income before taxes last year?

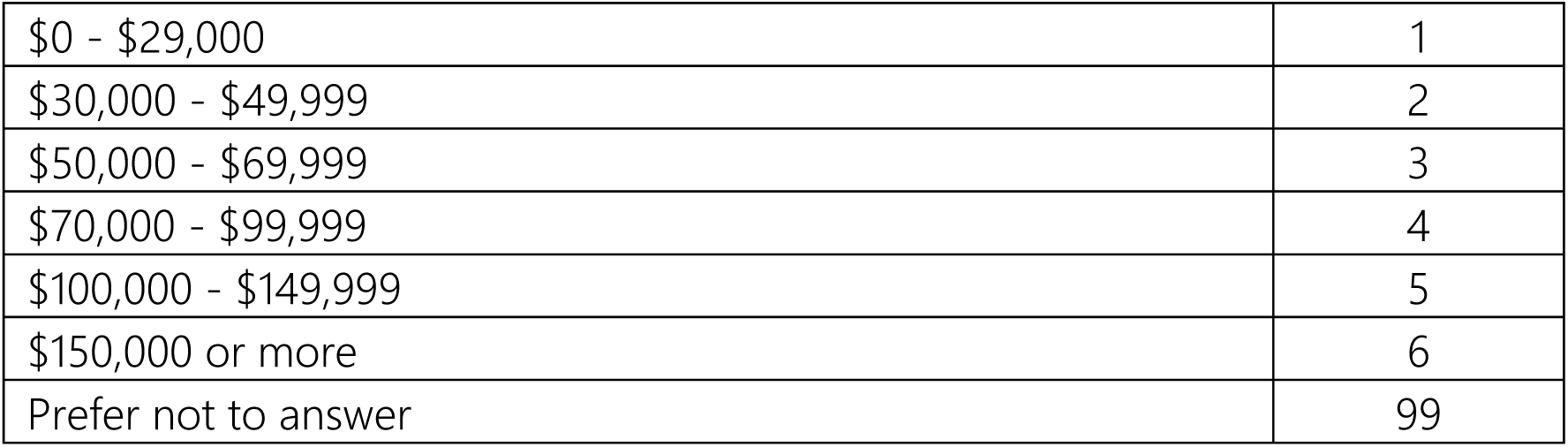
21. Including yourself, how many family members live in your household?

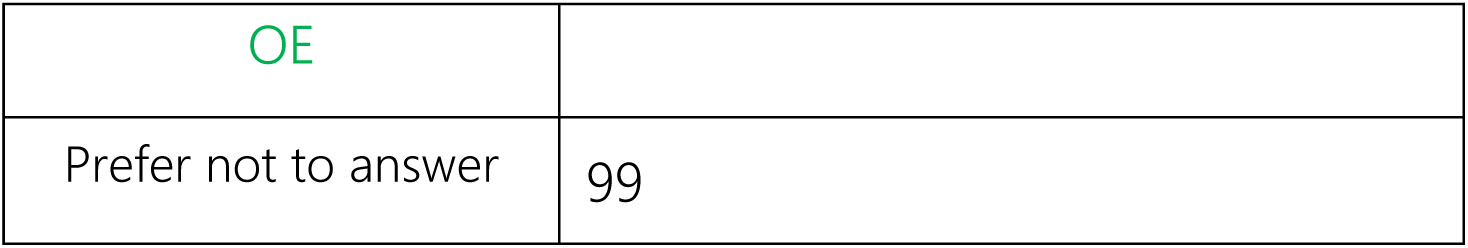 Final comments Please answer the following questions
22. How clear were the instructions to complete this survey?

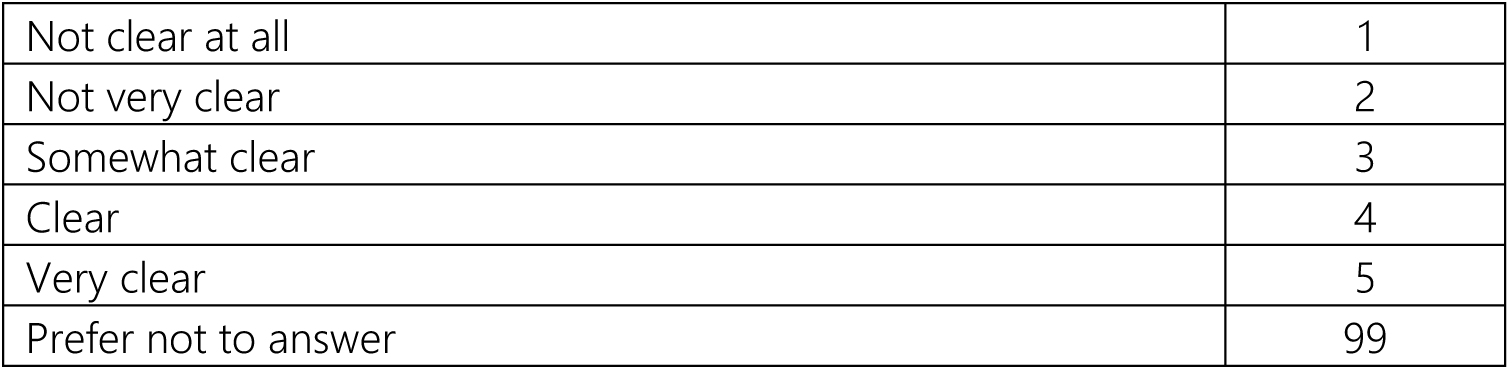
23. Please use this space to make any final comments.

21. Please use this space to make any final comments.
|  |  |
| --- | --- |
| Prefer not to answer | 99 |
| OE |  |

## Appendix 2: Stratified distributions of responses

i. Base case

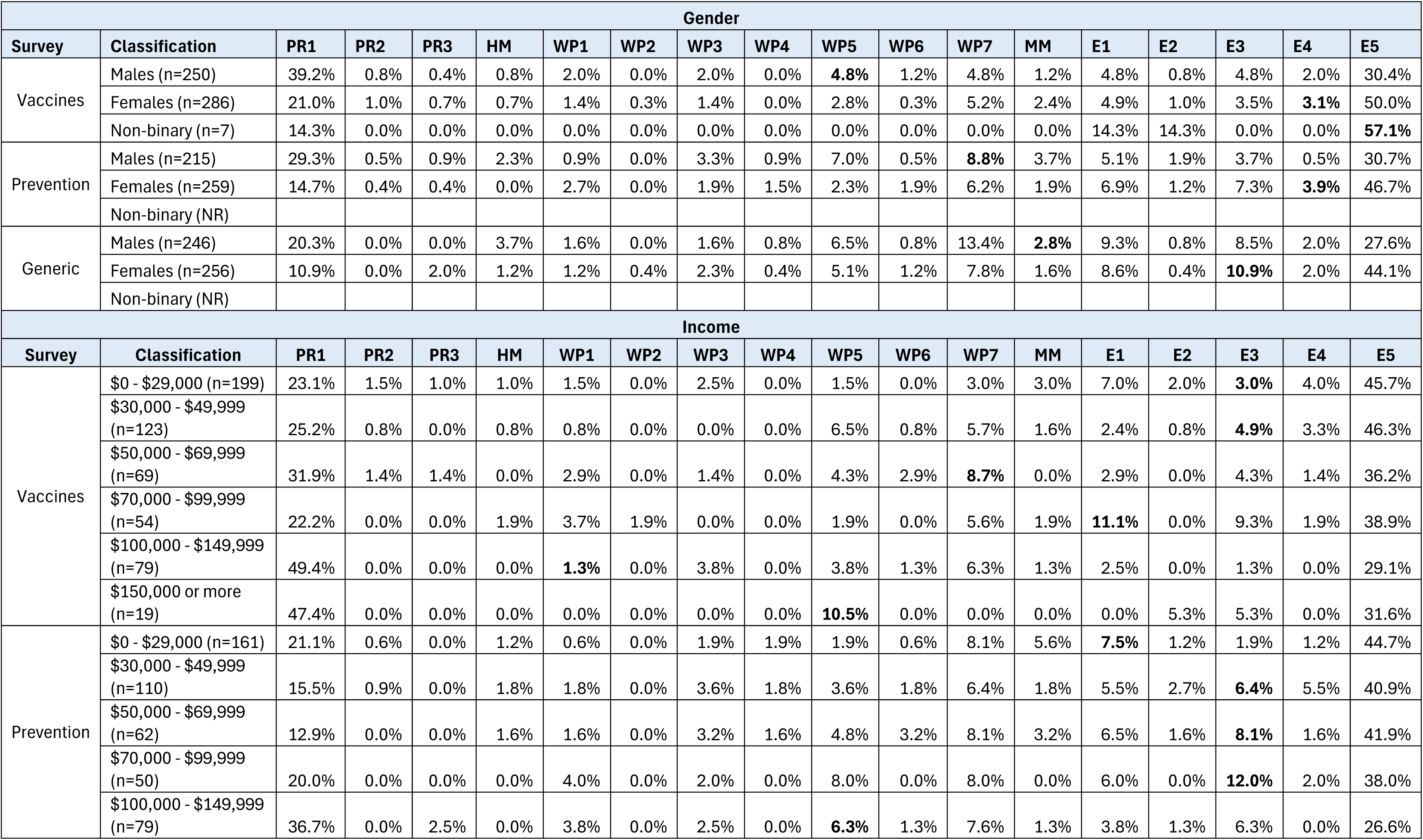
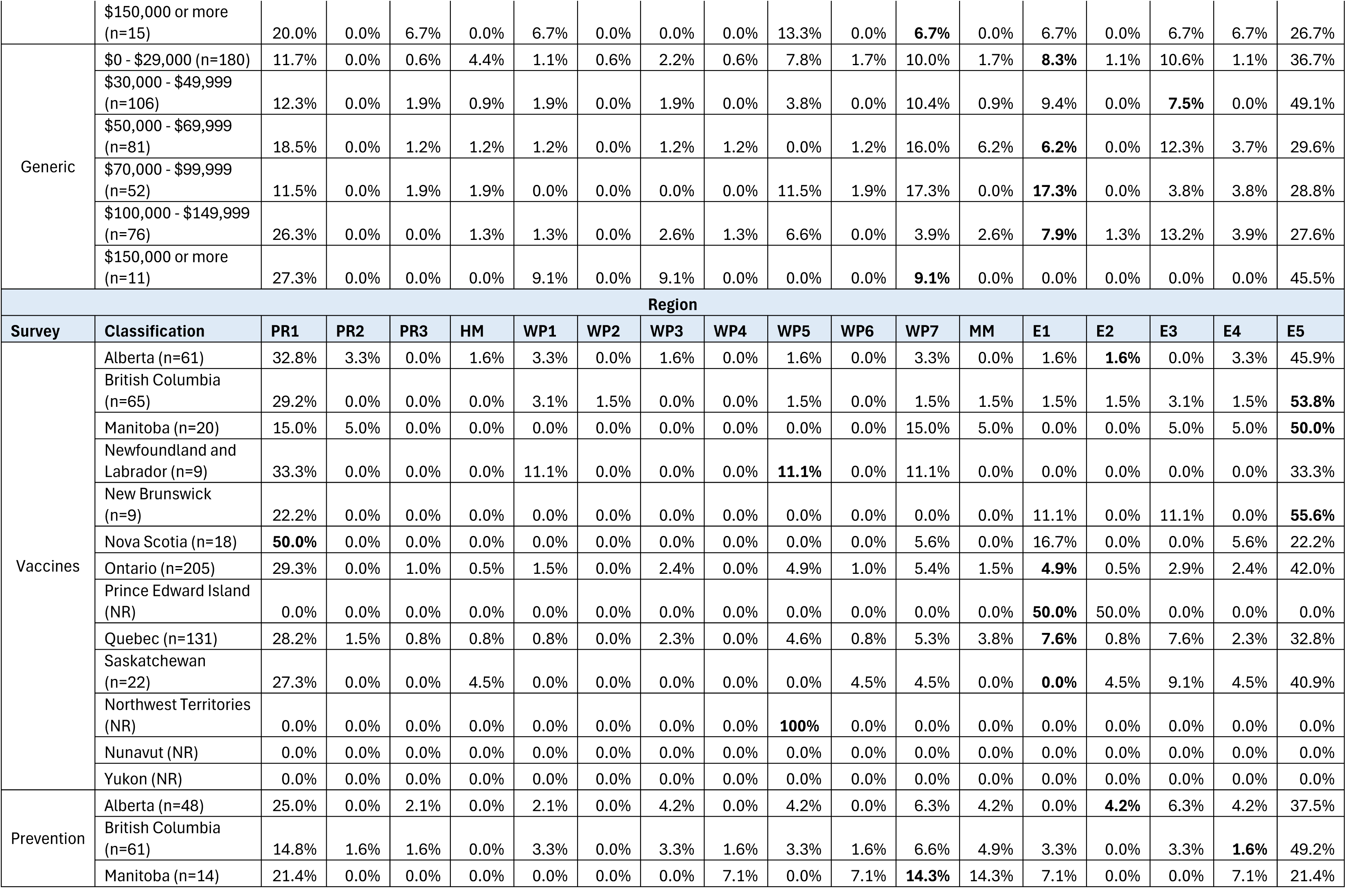
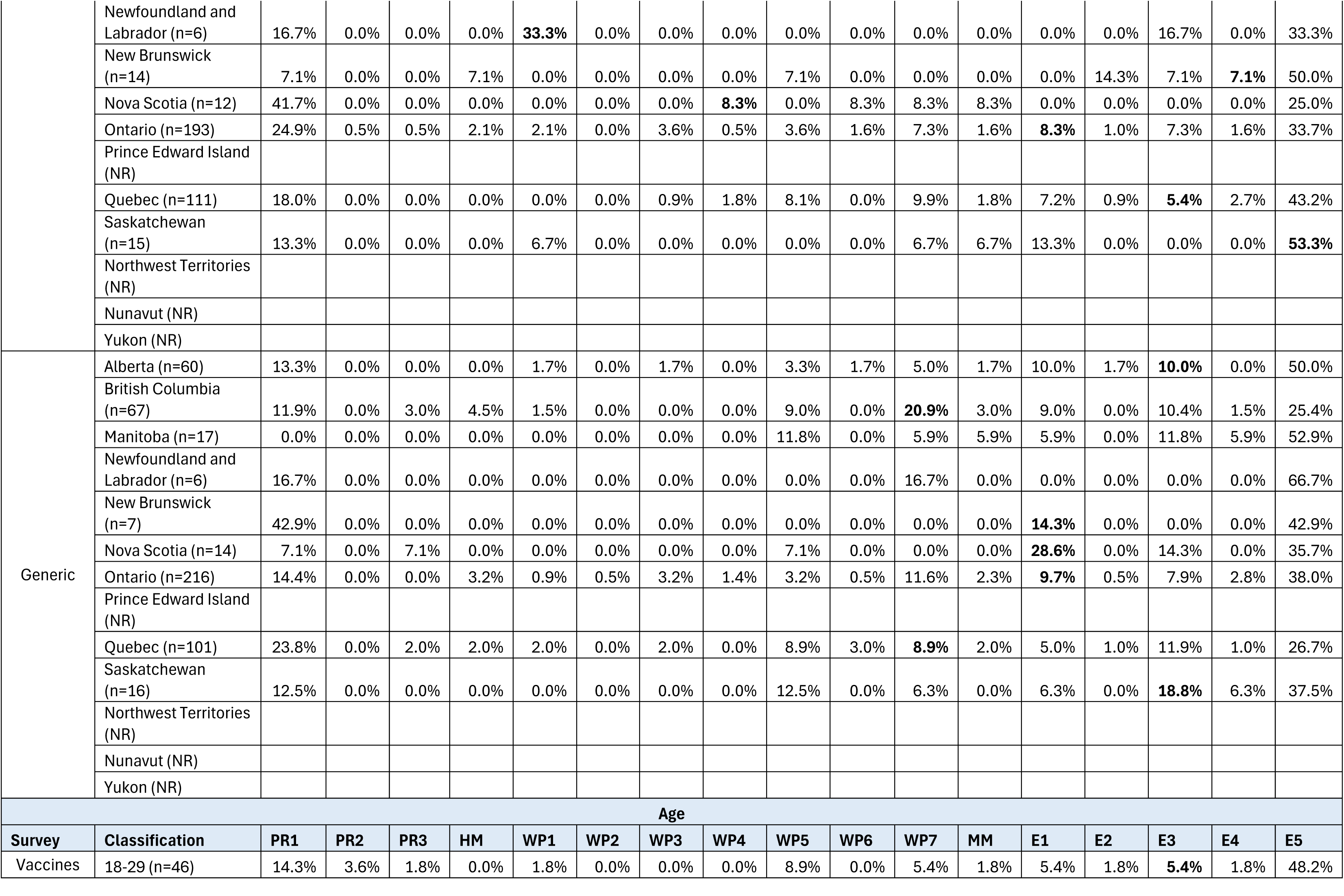
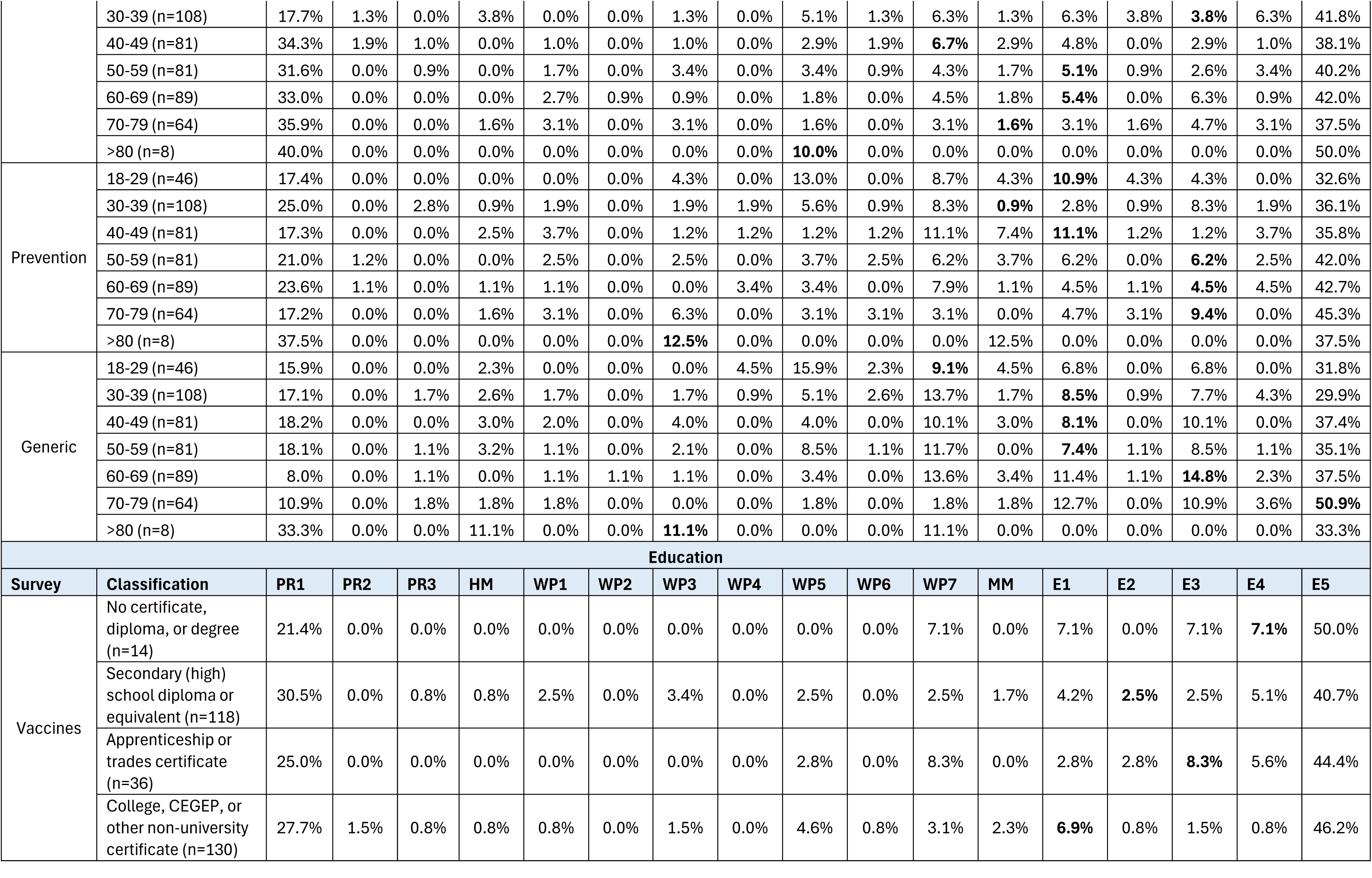
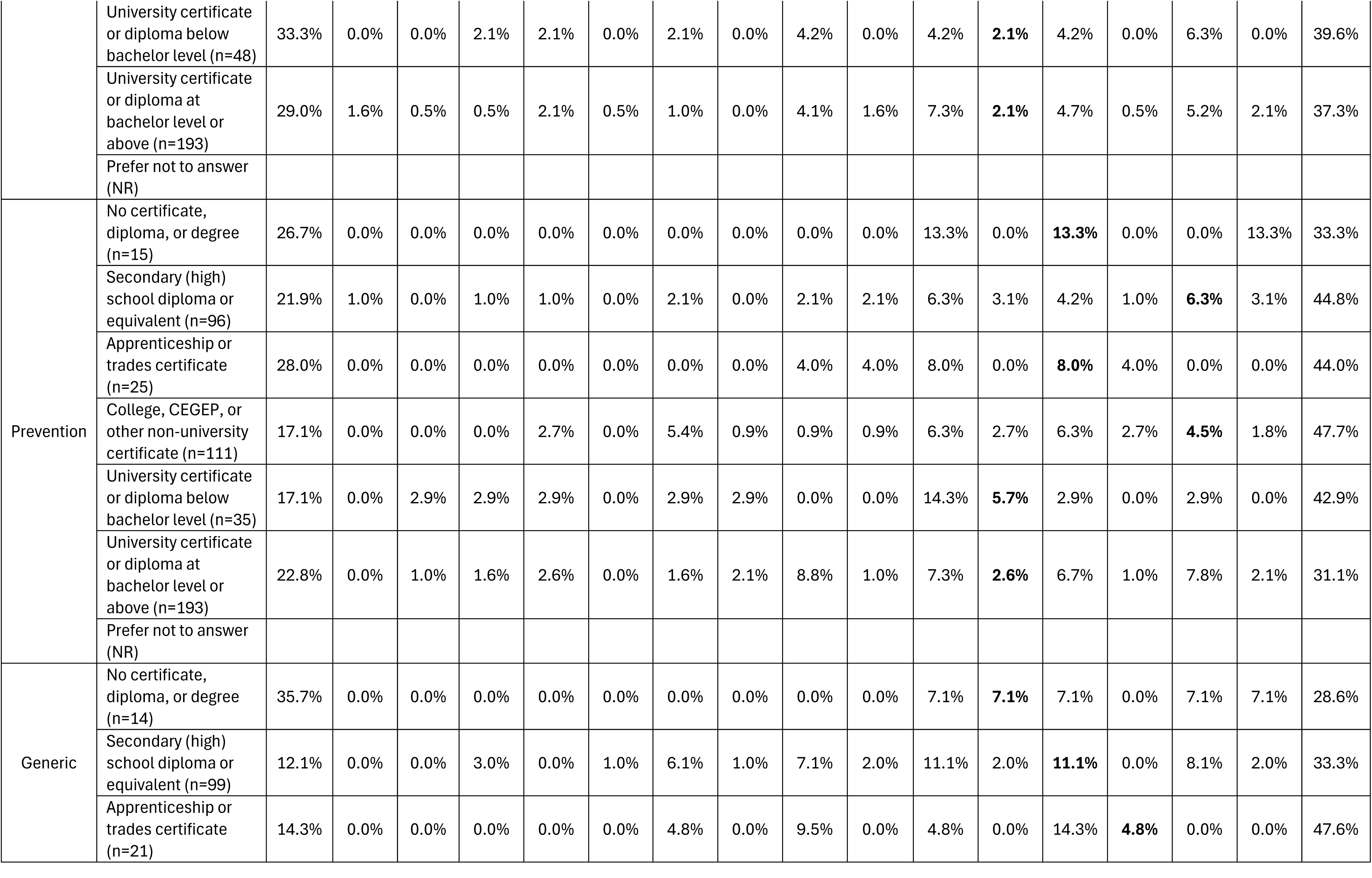
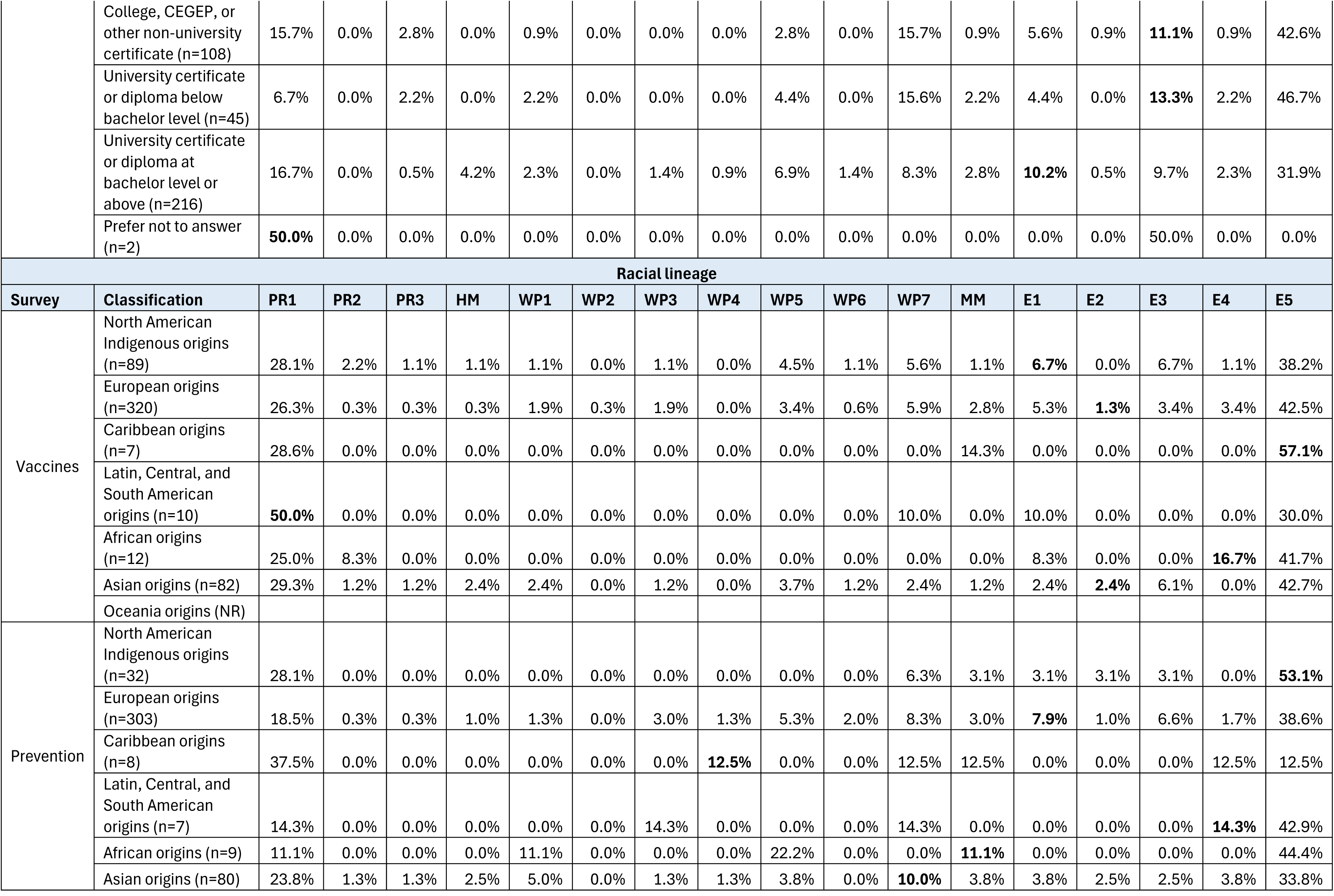
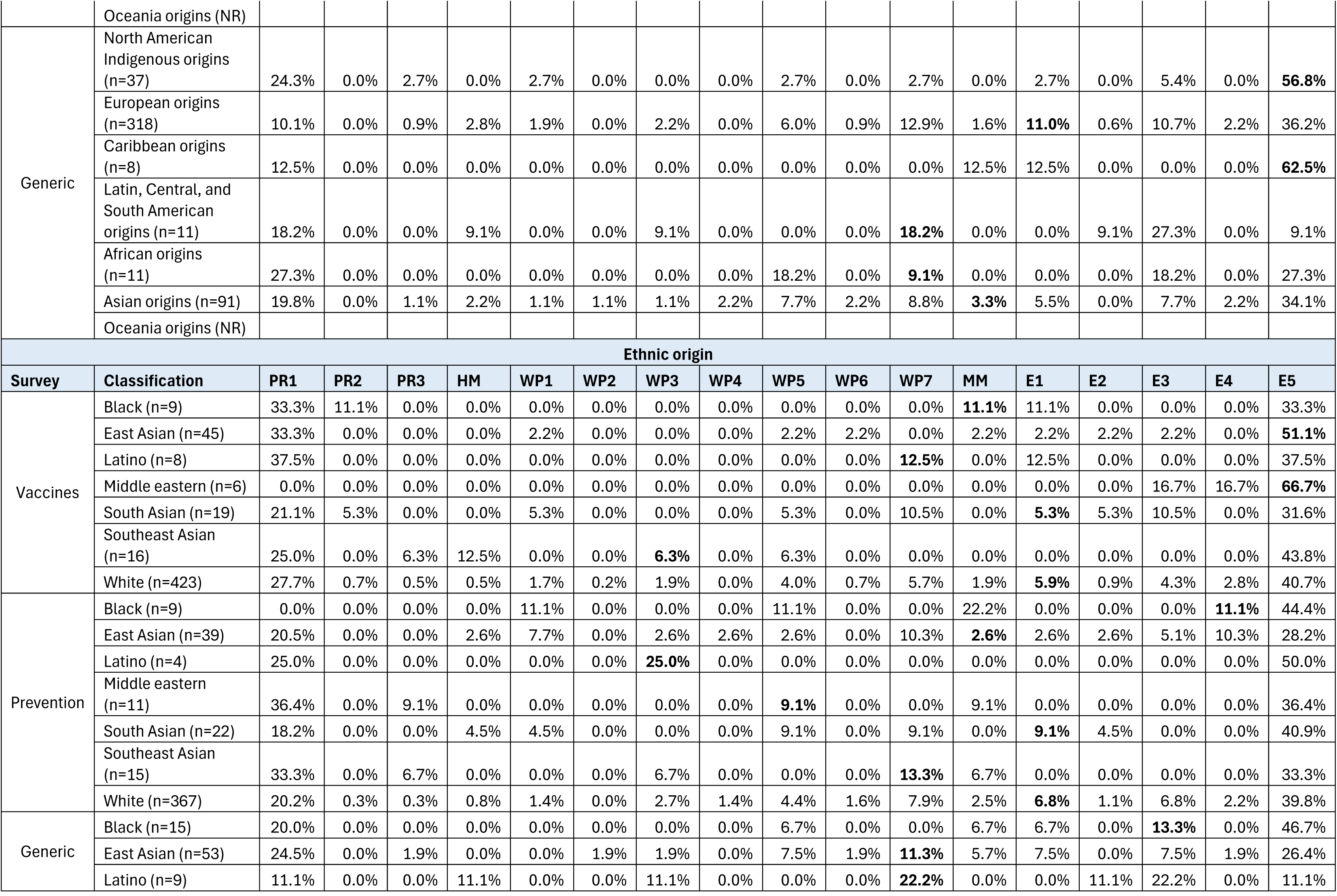
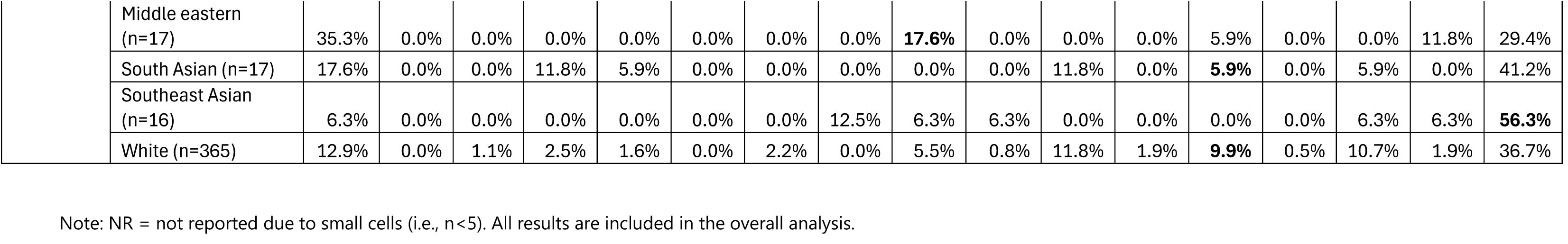
ii. Sensitivity analysis

(ii) Sensitivity analysis
| Gender |  |  |  |  |  |  |  |  |  |  |  |  |  |  |  |  |  |  |
| --- | --- | --- | --- | --- | --- | --- | --- | --- | --- | --- | --- | --- | --- | --- | --- | --- | --- | --- |
| Survey | Classification | PR1 | PR2 | PR3 | HM | WP1 | WP2 | WP3 | WP4 | WP5 | WP6 | WP7 | MM | E1 | E2 | E3 | E4 | E5 |
| Vaccines | Males (n=342) | 28.7% | 0.6% | 2.6% | 5.3% | 5.3% | 3.8% | 2.6% | **2.3%** | 5.0% | 2.9% | 4.7% | 3.2% | 4.4% | 1.5% | 3.5% | 1.5% | 22.2% |
|  | Females (n=386) | 15.5% | 1.0% | 2.6% | 5.7% | 3.9% | 2.3% | 2.3% | 1.6% | 3.9% | 3.6% | 5.4% | **2.6%** | 4.1% | 2.8% | 2.8% | 2.3% | 37.3% |
|  | Non-binary (n=9) | 11.1% | 0.0% | 0.0% | 0.0% | 11.1% | 0.0% | 0.0% | 0.0% | 0.0% | 0.0% | 0.0% | 11.1% | 11.1% | **11.1%** | 0.0% | 0.0% | 44.4% |
| Prevention | Males (n=350) | 18.0% | 1.7% | 5.4% | 8.0% | 4.3% | 1.7% | 5.4% | 3.7% | **6.3%** | 5.4% | 6.9% | 4.9% | 4.3% | 2.6% | 2.3% | 0.3% | 18.9% |
|  | Females (n=414) | 9.2% | 3.4% | 5.1% | 7.5% | 4.1% | 3.6% | 3.4% | 4.3% | 3.9% | 3.9% | **5.6%** | 2.7% | 5.1% | 2.2% | 4.6% | 2.4% | 29.2% |
|  | Non-binary (NR) |  |  |  |  |  |  |  |  |  |  |  |  |  |  |  |  |  |
| Generic | Males (n=367) | 13.6% | 1.1% | 4.1% | 7.1% | 5.4% | 3.0% | 4.6% | 2.7% | 5.2% | 3.0% | **10.9%** | 4.1% | 6.8% | 2.7% | 5.7% | 1.4% | 18.5% |
|  | Females (n=406) | 6.9% | 0.5% | 6.2% | 8.9% | 5.2% | 5.7% | 3.4% | 2.0% | 4.9% | 3.2% | **6.2%** | 2.7% | 6.4% | 2.0% | 6.9% | 1.2% | 27.8% |
|  | Non-binary (NR) |  |  |  |  |  |  |  |  |  |  |  |  |  |  |  |  |  |
| Income |  |  |  |  |  |  |  |  |  |  |  |  |  |  |  |  |  |  |
| Survey | Classification | PR1 | PR2 | PR3 | HM | WP1 | WP2 | WP3 | WP4 | WP5 | WP6 | WP7 | MM | E1 | E2 | E3 | E4 | E5 |
| Vaccines | \$0 - \$29,000 (n=269) | 17.1% | 1.1% | 4.1% | 5.6% | 3.7% | 1.9% | 3.7% | 2.2% | 2.2% | 3.0% | 3.3% | **4.1%** | 5.9% | 3.0% | 2.2% | 3.0% | 33.8% |
| | \$30,000 - \$49,999 (n=158) | 19.6% | 0.6% | 2.5% | 6.3% | 3.8% | 3.8% | 0.0% | 0.6% | 7.0% | 1.3% | **6.3%** | 1.3% | 1.9% | 2.5% | 3.8% | 2.5% | 36.1% |
| | \$50,000 - \$69,999 (n=99) | 22.2% | 1.0% | 2.0% | 5.1% | 5.1% | 4.0% | 3.0% | 2.0% | 5.1% | **5.1%** | 8.1% | 4.0% | 3.0% | 1.0% | 3.0% | 1.0% | 25.3% |
| | \$70,000 - \$99,999 (n=84) | 14.3% | 1.2% | 1.2% | 8.3% | 10.7% | 3.6% | 2.4% | 0.0% | 2.4% | 4.8% | **4.8%** | 3.6% | 8.3% | 1.2% | 6.0% | 1.2% | 26.2% |
| | \$100,000 - \$149,999 (n=103) | 37.9% | 0.0% | 1.0% | 2.9% | 2.9% | 2.9% | **2.9%** | 4.9% | 5.8% | 2.9% | 5.8% | 1.9% | 2.9% | 1.9% | 1.0% | 0.0% | 22.3% |
| | \$150,000 or more (n=24) | 37.5% | 0.0% | 0.0% | 0.0% | 4.2% | 4.2% | 0.0% | 0.0% | **8.3%** | 8.3% | 0.0% | 0.0% | 0.0% | 4.2% | 8.3% | 0.0% | 25.0% |
| Prevention | \$0 - \$29,000 (n=262) | 13.0% | 3.1% | 5.7% | 7.6% | 2.7% | 1.5% | 3.8% | 4.6% | 3.4% | **4.6%** | 7.6% | 6.1% | 5.7% | 1.1% | 1.1% | 0.8% | 27.5% |
| | \$30,000 - \$49,999 (n=175) | 9.7% | 3.4% | 4.6% | 8.6% | 4.0% | 2.9% | 5.7% | 4.0% | 4.6% | **2.9%** | 6.3% | 2.3% | 4.6% | 3.4% | 4.0% | 3.4% | 25.7% |
| | \$50,000 - \$69,999 (n=106) | 7.5% | 1.9% | 4.7% | 14.2% | 4.7% | 1.9% | 1.9% | 1.9% | 6.6% | **8.5%** | 4.7% | 3.8% | 4.7% | 2.8% | 4.7% | 0.9% | 24.5% |
| | \$70,000 - \$99,999 (n=88) | 11.4% | 0.0% | 3.4% | 6.8% | 9.1% | 2.3% | 3.4% | 3.4% | 6.8% | **6.8%** | 5.7% | 3.4% | 4.5% | 3.4% | 6.8% | 1.1% | 21.6% |
| | \$100,000 - \$149,999 (n=114) | 25.4% | 2.6% | 6.1% | 1.8% | 5.3% | 6.1% | **5.3%** | 5.3% | 5.3% | 2.6% | 5.3% | 0.9% | 2.6% | 2.6% | 4.4% | 0.0% | 18.4% |
| | \$150,000 or more (n=23) | 13.0% | 4.3% | 8.7% | 4.3% | 4.3% | 4.3% | 8.7% | **4.3%** | 8.7% | 0.0% | 4.3% | 4.3% | 4.3% | 0.0% | 4.3% | 4.3% | 17.4% |
| Generic | \$0 - \$29,000 (n=278) | 7.6% | 0.4% | 4.0% | 9.7% | 5.0% | 7.2% | 3.2% | 3.2% | 6.1% | 2.5% | **7.2%** | 4.7% | 6.1% | 1.8% | 6.8% | 0.7% | 23.7% |
| | \$30,000 - \$49,999 (n=160) | 8.1% | 1.3% | 5.6% | 6.9% | 5.6% | 1.9% | 4.4% | 1.3% | 3.8% | 1.3% | **10.0%** | 1.3% | 8.1% | 3.1% | 5.0% | 0.0% | 32.5% |
| | \$50,000 - \$69,999 (n=129) | 11.6% | 0.8% | 7.8% | 7.8% | 5.4% | 3.1% | 3.1% | 3.1% | 1.6% | **6.2%** | 12.4% | 3.9% | 3.9% | 0.8% | 7.8% | 2.3% | 18.6% |
| | \$70,000 - \$99,999 (n=88) | 6.8% | 0.0% | 5.7% | 5.7% | 4.5% | 5.7% | 4.5% | 1.1% | 10.2% | **5.7%** | 11.4% | 3.4% | 10.2% | 3.4% | 2.3% | 2.3% | 17.0% |
| | \$100,000 - \$149,999 (n=104) | 19.2% | 1.9% | 3.8% | 7.7% | 3.8% | 1.0% | 5.8% | 1.9% | **4.8%** | 1.0% | 3.8% | 3.8% | 5.8% | 2.9% | 9.6% | 2.9% | 20.2% |
| | \$150,000 or more (n=20) | 15.0% | 0.0% | 5.0% | 5.0% | 15.0% | 5.0% | **5.0%** | 0.0% | 5.0% | 5.0% | 5.0% | 0.0% | 5.0% | 5.0% | 0.0% | 0.0% | 25.0% |
| Region |  |  |  |  |  |  |  |  |  |  |  |  |  |  |  |  |  |  |
| Survey | Classification | PR1 | PR2 | PR3 | HM | WP1 | WP2 | WP3 | WP4 | WP5 | WP6 | WP7 | MM | E1 | E2 | E3 | E4 | E5 |
| Vaccines | Alberta (n=81) | 24.7% | 3.7% | 1.2% | 6.2% | 4.9% | 3.7% | 3.7% | **2.5%** | 2.5% | 2.5% | 2.5% | 1.2% | 1.2% | 2.5% | 0.0% | 2.5% | 34.6% |
|  | British Columbia (n=94) | 20.2% | 0.0% | 2.1% | 4.3% | 3.2% | 4.3% | 1.1% | 4.3% | 4.3% | 1.1% | 1.1% | **6.4%** | 2.1% | 4.3% | 3.2% | 1.1% | 37.2% |
|  | Manitoba (n=31) | 9.7% | 3.2% | 3.2% | 3.2% | 3.2% | 3.2% | 3.2% | 3.2% | 3.2% | 9.7% | **9.7%** | 3.2% | 0.0% | 3.2% | 3.2% | 3.2% | 32.3% |
|  | Newfoundland and Labrador (n=12) | 25.0% | 0.0% | 0.0% | 0.0% | **25.0%** | 8.3% | 0.0% | 0.0% | 8.3% | 0.0% | 8.3% | 0.0% | 0.0% | 0.0% | 0.0% | 0.0% | 25.0% |
|  | New Brunswick (n=15) | 13.3% | 0.0% | 0.0% | 13.3% | 6.7% | 6.7% | 0.0% | 6.7% | 0.0% | 0.0% | 0.0% | **6.7%** | 6.7% | 0.0% | 6.7% | 0.0% | 33.3% |
|  | Nova Scotia (n=22) | 40.9% | 0.0% | 4.5% | 0.0% | 0.0% | 0.0% | **4.5%** | 0.0% | 0.0% | 9.1% | 4.5% | 0.0% | 13.6% | 0.0% | 0.0% | 4.5% | 18.2% |
|  | Ontario (n=283) | 21.2% | 0.0% | 3.9% | 5.7% | 4.9% | 2.5% | 2.8% | 1.1% | 4.6% | **4.2%** | 6.0% | 2.1% | 4.6% | 1.8% | 2.1% | 1.8% | 30.7% |
|  | Prince Edward Island (NR) |  |  |  |  |  |  |  |  |  |  |  |  |  |  |  |  |  |
|  | Quebec (n=165) | 22.4% | 1.2% | 1.2% | 5.5% | 4.2% | 1.8% | 2.4% | 1.8% | 4.8% | 1.8% | **6.7%** | 4.2% | 6.7% | 1.2% | 6.1% | 1.8% | 26.1% |
|  | Saskatchewan (n=30) | 20.0% | 0.0% | 3.3% | 10.0% | 3.3% | 6.7% | 0.0% | 0.0% | 3.3% | **3.3%** | 3.3% | 0.0% | 0.0% | 6.7% | 6.7% | 3.3% | 30.0% |
|  | Northwest Territories (NR) |  |  |  |  |  |  |  |  |  |  |  |  |  |  |  |  |  |
|  | Nunavut (NR) |  |  |  |  |  |  |  |  |  |  |  |  |  |  |  |  |  |
|  | Yukon (NR) |  |  |  |  |  |  |  |  |  |  |  |  |  |  |  |  |  |
| Prevention | Alberta (n=87) | 13.8% | 1.1% | 6.9% | 10.3% | 5.7% | 2.3% | 5.7% | **6.9%** | 2.3% | **5.7%** | 5.7% | 2.3% | 1.1% | 3.4% | 3.4% | 2.3% | 20.7% |
|  | British Columbia (n=98) | 9.2% | 4.1% | 4.1% | 5.1% | 5.1% | 5.1% | 5.1% | 4.1% | 4.1% | 8.2% | **6.1%** | 3.1% | 2.0% | 1.0% | 2.0% | 1.0% | 30.6% |
|  | Manitoba (n=26) | 11.5% | 0.0% | 3.8% | 11.5% | 0.0% | 0.0% | 3.8% | 11.5% | 0.0% | **3.8%** | 11.5% | 15.4% | 3.8% | 7.7% | 0.0% | 3.8% | 11.5% |
|  | Newfoundland and Labrador (n=7) | 14.3% | 0.0% | 0.0% | 0.0% | 28.6% | 0.0% | **14.3%** | 0.0% | 0.0% | 0.0% | 0.0% | 0.0% | 0.0% | 0.0% | 14.3% | 0.0% | 28.6% |
|  | New Brunswick (n=21) | 4.8% | 0.0% | 0.0% | 14.3% | 0.0% | 0.0% | 0.0% | 4.8% | 4.8% | 14.3% | 4.8% | 0.0% | 0.0% | **9.5%** | 4.8% | 4.8% | 33.3% |
|  | Nova Scotia (n=22) | 22.7% | 0.0% | 4.5% | 4.5% | 13.6% | 0.0% | 0.0% | 4.5% | **4.5%** | 13.6% | 9.1% | 4.5% | 0.0% | 4.5% | 0.0% | 0.0% | 13.6% |
|  | Ontario (n=306) | 15.7% | 2.9% | 4.9% | 8.5% | 4.2% | 3.6% | 5.2% | 2.3% | **4.6%** | 3.9% | 5.6% | 3.6% | 6.2% | 2.0% | 4.6% | 1.0% | 21.2% |
|  | Prince Edward Island (n=6) | 0.0% | 16.7% | 0.0% | **33.3%** | 0.0% | 0.0% | 0.0% | 16.7% | 0.0% | 0.0% | 0.0% | 0.0% | 0.0% | 0.0% | 0.0% | 0.0% | 33.3% |
|  | Quebec (n=170) | 11.8% | 2.9% | 6.5% | 5.3% | 2.4% | 1.8% | 2.9% | 3.5% | 9.4% | 0.6% | **7.6%** | 4.1% | 6.5% | 1.2% | 3.5% | 1.8% | 28.2% |
|  | Saskatchewan (n=23) | 8.7% | 0.0% | 8.7% | 4.3% | 8.7% | 0.0% | 0.0% | 8.7% | 0.0% | 8.7% | **4.3%** | 4.3% | 8.7% | 0.0% | 0.0% | 0.0% | 34.8% |
|  | Northwest Territories (NR) |  |  |  |  |  |  |  |  |  |  |  |  |  |  |  |  |  |
|  | Nunavut (NR) |  |  |  |  |  |  |  |  |  |  |  |  |  |  |  |  |  |
|  | Yukon (NR) |  |  |  |  |  |  |  |  |  |  |  |  |  |  |  |  |  |
| Generic | Alberta (n=88) | 9.1% | 0.0% | 1.1% | 2.3% | 8.0% | 4.5% | 4.5% | 2.3% | 3.4% | 5.7% | 4.5% | 3.4% | **6.8%** | 3.4% | 6.8% | 0.0% | 34.1% |
|  | British Columbia (n=102) | 7.8% | 1.0% | 6.9% | 4.9% | 4.9% | 7.8% | 3.9% | 1.0% | 6.9% | 2.9% | **14.7%** | 3.9% | 5.9% | 2.9% | 6.9% | 1.0% | 16.7% |
|  | Manitoba (n=28) | 0.0% | 0.0% | 3.6% | 3.6% | 3.6% | 7.1% | 0.0% | 3.6% | 14.3% | 3.6% | 3.6% | **10.7%** | 3.6% | 0.0% | 7.1% | 3.6% | 32.1% |
|  | Newfoundland and Labrador (n=12) | 8.3% | 8.3% | 0.0% | 25.0% | 0.0% | 0.0% | **8.3%** | 0.0% | 0.0% | 0.0% | 8.3% | 0.0% | 0.0% | 8.3% | 0.0% | 0.0% | 33.3% |
|  | New Brunswick (n=18) | 16.7% | 5.6% | 5.6% | **27.8%** | 5.6% | 0.0% | 0.0% | 5.6% | 0.0% | 0.0% | 5.6% | 0.0% | 5.6% | 5.6% | 0.0% | 0.0% | 16.7% |
|  | Nova Scotia (n=24) | 4.2% | 0.0% | 8.3% | 4.2% | 8.3% | 8.3% | 4.2% | 0.0% | 4.2% | 0.0% | 0.0% | 4.2% | **16.7%** | 8.3% | 8.3% | 0.0% | 20.8% |
|  | Ontario (n=300) | 10.3% | 0.7% | 5.0% | 7.0% | 4.0% | 4.0% | 4.0% | 3.7% | 3.7% | 2.0% | **9.3%** | 3.3% | 7.7% | 0.3% | 5.7% | 2.0% | 27.3% |
|  | Prince Edward Island (NR) |  |  |  |  |  |  |  |  |  |  |  |  |  |  |  |  |  |
|  | Quebec (n=178) | 13.5% | 0.6% | 6.7% | 12.4% | 5.1% | 3.4% | 3.9% | 1.1% | **6.7%** | 3.9% | 7.9% | 3.4% | 5.1% | 3.9% | 6.7% | 0.6% | 15.2% |
|  | Saskatchewan (n=23) | 8.7% | 0.0% | 0.0% | 0.0% | 8.7% | 0.0% | 8.7% | 0.0% | 8.7% | 8.7% | **8.7%** | 0.0% | 4.3% | 0.0% | 13.0% | 4.3% | 26.1% |
|  | Northwest Territories (NR) |  |  |  |  |  |  |  |  |  |  |  |  |  |  |  |  |  |
|  | Nunavut (NR) |  |  |  |  |  |  |  |  |  |  |  |  |  |  |  |  |  |
|  | Yukon (NR) |  |  |  |  |  |  |  |  |  |  |  |  |  |  |  |  |  |
| Age |  |  |  |  |  |  |  |  |  |  |  |  |  |  |  |  |  |  |

| Survey | Classification | PR1 | PR2 | PR3 | HM | WP1 | WP2 | WP3 | WP4 | WP5 | WP6 | WP7 | MM | E1 | E2 | E3 | E4 | E5 |
| --- | --- | --- | --- | --- | --- | --- | --- | --- | --- | --- | --- | --- | --- | --- | --- | --- | --- | --- |
| Vaccines | 18-29 (n=93) | 8.6% | 2.2% | 5.4% | 6.5% | 5.4% | 7.5% | 2.2% | 1.1% | 6.5% | 4.3% | **6.5%** | 3.2% | 4.3% | 3.2% | 3.2% | 1.1% | 29.0% |
|  | 30-39 (n=108) | 13.0% | 1.9% | 2.8% | 7.4% | 4.6% | 0.9% | 2.8% | 0.9% | 6.5% | 3.7% | **6.5%** | 2.8% | 4.6% | 3.7% | 2.8% | 4.6% | 30.6% |
|  | 40-49 (n=137) | 26.3% | 1.5% | 2.9% | 2.2% | 3.6% | 2.2% | 1.5% | 2.9% | 3.6% | 2.9% | **7.3%** | 2.9% | 5.1% | 2.2% | 2.2% | 0.7% | 29.9% |
|  | 50-59 (n=147) | 25.2% | 0.0% | 1.4% | 4.8% | 4.1% | 2.7% | 2.7% | 1.4% | 4.8% | **4.8%** | 3.4% | 1.4% | 4.8% | 2.0% | 2.0% | 2.7% | 32.0% |
|  | 60-69 (n=148) | 25.0% | 0.0% | 2.0% | 6.1% | 4.1% | 2.0% | 2.0% | 3.4% | 2.0% | 2.0% | **3.4%** | 4.1% | 4.7% | 1.4% | 5.4% | 0.7% | 31.8% |
|  | 70-79 (n=87) | 26.4% | 0.0% | 2.3% | 6.9% | 6.9% | 2.3% | 3.4% | 1.1% | **2.3%** | 2.3% | 4.6% | 3.4% | 2.3% | 2.3% | 3.4% | 2.3% | 27.6% |
|  | >80 (n=17) | 23.5% | 0.0% | 0.0% | 5.9% | 5.9% | 11.8% | **5.9%** | 0.0% | 11.8% | 0.0% | 0.0% | 5.9% | 0.0% | 0.0% | 0.0% | 0.0% | 29.4% |
| Prevention | 18-29 (n=91) | 8.8% | 3.3% | 11.0% | 2.2% | 5.5% | 5.5% | 8.8% | 4.4% | **8.8%** | 2.2% | 6.6% | 4.4% | 6.6% | 3.3% | 2.2% | 0.0% | 16.5% |
|  | 30-39 (n=171) | 15.8% | 2.3% | 5.3% | 5.8% | 4.7% | 5.3% | 4.1% | 3.5% | **5.8%** | 5.8% | 6.4% | 1.8% | 2.3% | 1.8% | 5.3% | 1.2% | 22.8% |
|  | 40-49 (n=141) | 9.9% | 5.0% | 2.8% | 9.9% | 5.0% | 1.4% | 2.1% | 4.3% | 3.5% | **6.4%** | 9.2% | 6.4% | 7.1% | 3.5% | 0.7% | 2.1% | 20.6% |
|  | 50-59 (n=123) | 13.8% | 2.4% | 4.9% | 9.8% | 3.3% | 0.8% | 2.4% | 4.1% | 4.1% | **5.7%** | 4.9% | 5.7% | 4.9% | 0.0% | 4.1% | 1.6% | 27.6% |
|  | 60-69 (n=139) | 15.1% | 1.4% | 4.3% | 8.6% | 3.6% | 1.4% | 2.2% | 7.2% | 3.6% | **2.9%** | 6.5% | 2.9% | 3.6% | 3.6% | 2.9% | 2.9% | 27.3% |
|  | 70-79 (n=92) | 12.0% | 1.1% | 5.4% | 8.7% | 4.3% | 2.2% | 8.7% | 0.0% | 5.4% | **3.3%** | 3.3% | 1.1% | 4.3% | 2.2% | 6.5% | 0.0% | 31.5% |
|  | >80 (n=11) | 27.3% | 0.0% | 0.0% | 9.1% | 9.1% | 0.0% | **9.1%** | 0.0% | 0.0% | 0.0% | 0.0% | 9.1% | 9.1% | 0.0% | 0.0% | 0.0% | 27.3% |
| Generic | 18-29 (n=93) | 7.5% | 0.0% | 6.5% | 10.8% | 1.1% | 6.5% | 6.5% | 5.4% | **11.8%** | 5.4% | 8.6% | 5.4% | 4.3% | 2.2% | 3.2% | 0.0% | 15.1% |
|  | 30-39 (n=170) | 11.8% | 0.6% | 6.5% | 8.8% | 6.5% | 4.1% | 4.1% | 0.6% | 4.7% | **4.1%** | 10.0% | 1.2% | 6.5% | 1.8% | 5.3% | 2.9% | 20.6% |
|  | 40-49 (n=143) | 12.6% | 2.1% | 4.2% | 6.3% | 4.9% | 2.1% | 4.9% | 2.1% | 4.2% | 2.8% | **8.4%** | 4.2% | 6.3% | 2.1% | 7.0% | 0.0% | 25.9% |
|  | 50-59 (n=138) | 12.3% | 0.0% | 3.6% | 8.7% | 5.1% | 2.2% | 2.2% | 2.9% | 6.5% | 3.6% | **9.4%** | 3.6% | 5.8% | 3.6% | 5.8% | 0.7% | 23.9% |
|  | 60-69 (n=138) | 5.1% | 1.4% | 5.1% | 6.5% | 4.3% | 8.0% | 3.6% | 3.6% | 2.9% | 0.7% | **9.4%** | 4.3% | 7.2% | 2.9% | 9.4% | 1.4% | 23.9% |
|  | 70-79 (n=84) | 7.1% | 0.0% | 3.6% | 7.1% | 9.5% | 4.8% | 1.2% | 0.0% | 2.4% | 2.4% | 3.6% | 3.6% | **10.7%** | 1.2% | 7.1% | 2.4% | 33.3% |
|  | >80 (n=13) | 23.1% | 0.0% | 15.4% | 7.7% | **7.7%** | 0.0% | 15.4% | 0.0% | 0.0% | 0.0% | 7.7% | 0.0% | 0.0% | 0.0% | 0.0% | 0.0% | 23.1% |
| Education |  |  |  |  |  |  |  |  |  |  |  |  |  |  |  |  |  |  |
| Survey | Classification | PR1 | PR2 | PR3 | HM | WP1 | WP2 | WP3 | WP4 | WP5 | WP6 | WP7 | MM | E1 | E2 | E3 | E4 | E5 |
| Vaccines | No certificate, diploma, or degree (n=17) | 17.6% | 0.0% | 0.0% | 0.0% | 0.0% | 5.9% | 5.9% | 0.0% | 0.0% | 0.0% | 11.8% | 0.0% | 5.9% | 0.0% | **5.9%** | 5.9% | 41.2% |
|  | Secondary (high) school diploma or equivalent (n=160) | 22.5% | 0.6% | 3.8% | 5.0% | 6.3% | 1.9% | 3.1% | 1.3% | 3.1% | **3.1%** | 3.8% | 2.5% | 3.8% | 3.1% | 2.5% | 3.8% | 30.0% |
|  | Apprenticeship or trades certificate (n=46) | 19.6% | 0.0% | 2.2% | 10.9% | 0.0% | 0.0% | 0.0% | 4.3% | 2.2% | 0.0% | 6.5% | **4.3%** | 2.2% | 2.2% | 6.5% | 4.3% | 34.8% |
|  | College, CEGEP, or other non-university certificate (n=191) | 18.8% | 1.0% | 2.6% | 6.8% | 4.7% | 2.6% | 2.1% | 2.6% | 5.8% | 2.6% | **3.7%** | 4.2% | 5.8% | 3.1% | 1.0% | 0.5% | 31.9% |
|  | University certificate or diploma below bachelor level (n=68) | 23.5% | 0.0% | 4.4% | 2.9% | 5.9% | 7.4% | 2.9% | 1.5% | **4.4%** | 1.5% | 4.4% | 4.4% | 2.9% | 1.5% | 4.4% | 0.0% | 27.9% |
|  | University certificate or diploma at bachelor level or above (n=250) | 22.4% | 1.2% | 1.6% | 4.8% | 4.4% | 3.2% | 2.4% | 1.2% | 4.8% | **5.2%** | 6.4% | 2.0% | 4.4% | 1.6% | 4.0% | 1.6% | 28.8% |
|  | Prefer not to answer (n=5) | **60.0%** | 0.0% | 0.0% | 0.0% | 0.0% | 0.0% | 0.0% | 20.0% | 0.0% | 0.0% | 0.0% | 0.0% | 0.0% | 0.0% | 0.0% | 0.0% | 20.0% |
| Prevention | No certificate, diploma, or degree (n=25) | 16.0% | 8.0% | 8.0% | 0.0% | 8.0% | 4.0% | 4.0% | 0.0% | 0.0% | **4.0%** | 12.0% | 0.0% | 8.0% | 0.0% | 0.0% | 8.0% | 20.0% |
|  | Secondary (high) school diploma or equivalent (n=154) | 13.6% | 3.2% | 5.8% | 9.7% | 1.9% | 3.2% | 5.2% | 3.9% | 2.6% | **2.6%** | 4.5% | 5.2% | 2.6% | 1.9% | 3.9% | 1.9% | 27.9% |
|  | Apprenticeship or trades certificate (n=34) | 20.6% | 2.9% | 0.0% | 2.9% | 0.0% | 5.9% | 5.9% | 2.9% | 8.8% | **2.9%** | 5.9% | 0.0% | 5.9% | 2.9% | 0.0% | 0.0% | 32.4% |
|  | College, CEGEP, or other non-university certificate (n=190) | 10.0% | 3.2% | 4.2% | 10.0% | 3.2% | 1.1% | 5.3% | 3.2% | 3.2% | 4.7% | **6.3%** | 3.7% | 7.4% | 3.2% | 2.6% | 1.1% | 27.9% |
|  | University certificate or diploma below bachelor level (n=59) | 10.2% | 1.7% | 8.5% | 10.2% | 6.8% | 1.7% | 3.4% | 5.1% | **3.4%** | 5.1% | 10.2% | 5.1% | 1.7% | 0.0% | 1.7% | 0.0% | 25.4% |
|  | University certificate or diploma at bachelor level or above (n=302) | 14.6% | 1.3% | 5.3% | 5.6% | 6.0% | 3.3% | 3.3% | 5.0% | **7.6%** | 5.6% | 6.0% | 3.3% | 4.3% | 2.6% | 5.0% | 1.3% | 19.9% |
|  | Prefer not to answer (NR) |  |  |  |  |  |  |  |  |  |  |  |  |  |  |  |  |  |
| Generic | No certificate, diploma, or degree (n=21) | 23.8% | 0.0% | 4.8% | 4.8% | 0.0% | 9.5% | 0.0% | 4.8% | 0.0% | 0.0% | **9.5%** | 4.8% | 9.5% | 0.0% | 4.8% | 4.8% | 19.0% |
|  | Secondary (high) school diploma or equivalent (n=169) | 7.1% | 0.6% | 6.5% | 14.2% | 3.0% | 6.5% | 5.3% | 3.6% | **4.7%** | 2.4% | 7.7% | 4.1% | 7.7% | 1.2% | 4.7% | 1.2% | 19.5% |
|  | Apprenticeship or trades certificate (n=34) | 8.8% | 0.0% | 0.0% | 17.6% | 2.9% | 2.9% | 8.8% | 2.9% | **5.9%** | 2.9% | 2.9% | 0.0% | 8.8% | 5.9% | 0.0% | 0.0% | 29.4% |
|  | College, CEGEP, or other non-university certificate (n=174) | 9.8% | 1.1% | 6.3% | 5.7% | 6.9% | 1.7% | 2.3% | 4.0% | 3.4% | 2.9% | **12.1%** | 2.9% | 4.0% | 2.9% | 6.9% | 0.6% | 26.4% |
|  | University certificate or diploma below bachelor level (n=68) | 5.9% | 1.5% | 10.3% | 2.9% | 10.3% | 2.9% | 1.5% | 0.0% | 4.4% | 0.0% | **10.3%** | 4.4% | 2.9% | 1.5% | 8.8% | 1.5% | 30.9% |

|  | University certificate or diploma at bachelor level or above (n=310) | 11.6% | 0.6% | 3.2% | 6.1% | 5.2% | 4.8% | 4.5% | 1.0% | 6.8% | 4.5% | **7.4%** | 3.2% | 7.7% | 2.6% | 6.8% | 1.6% | 22.3% |
| --- | --- | --- | --- | --- | --- | --- | --- | --- | --- | --- | --- | --- | --- | --- | --- | --- | --- | --- |
|  | Prefer not to answer (NR) |  |  |  |  |  |  |  |  |  |  |  |  |  |  |  |  |  |
| Racial lineage |  |  |  |  |  |  |  |  |  |  |  |  |  |  |  |  |  |  |
| Survey | Classification | PR1 | PR2 | PR3 | HM | WP1 | WP2 | WP3 | WP4 | WP5 | WP6 | WP7 | MM | E1 | E2 | E3 | E4 | E5 |
| Vaccines | North American Indigenous origins (n=117) | 21.4% | 1.7% | 0.9% | 6.8% | 6.0% | 0.9% | 1.7% | 2.6% | 6.0% | **3.4%** | 6.0% | 2.6% | 5.1% | 0.0% | 5.1% | 0.9% | 29.1% |
|  | European origins (n=438) | 19.2% | 0.5% | 2.3% | 4.3% | 4.3% | 3.2% | 2.5% | 2.3% | 4.6% | 3.2% | **5.7%** | 4.1% | 4.3% | 3.0% | 2.7% | 2.5% | 31.3% |
|  | Caribbean origins (n=13) | 15.4% | 0.0% | 7.7% | 7.7% | 0.0% | 7.7% | 0.0% | 0.0% | 7.7% | **7.7%** | 0.0% | 7.7% | 7.7% | 0.0% | 0.0% | 0.0% | 30.8% |
|  | Latin, Central, and South American origins (n=16) | 31.3% | 0.0% | 12.5% | **6.3%** | 0.0% | 6.3% | 0.0% | 0.0% | 0.0% | 12.5% | 6.3% | 0.0% | 6.3% | 0.0% | 0.0% | 0.0% | 18.8% |
|  | African origins (n=18) | 16.7% | 5.6% | 5.6% | 0.0% | 5.6% | 0.0% | 11.1% | 0.0% | **5.6%** | 0.0% | 5.6% | 0.0% | 5.6% | 0.0% | 0.0% | 11.1% | 27.8% |
|  | Asian origins (n=109) | 22.0% | 0.9% | 3.7% | 7.3% | 5.5% | 3.7% | 1.8% | 0.9% | **4.6%** | 1.8% | 3.7% | 0.9% | 2.8% | 3.7% | 4.6% | 0.0% | 32.1% |
|  | Oceania origins (NR) |  |  |  |  |  |  |  |  |  |  |  |  |  |  |  |  |  |
| Prevention | North American Indigenous origins (n=67) | 14.9% | 3.0% | 4.5% | 4.5% | 9.0% | 3.0% | 3.0% | 6.0% | **6.0%** | 1.5% | 4.5% | 6.0% | 3.0% | 3.0% | 7.5% | 3.0% | 17.9% |
|  | European origins (n=437) | 16.0% | 2.5% | 5.3% | 7.6% | 4.1% | 1.8% | 3.2% | 3.7% | 4.1% | **4.8%** | 8.2% | 3.7% | 3.4% | 2.3% | 3.7% | 1.8% | 23.8% |
|  | Caribbean origins (n=13) | 7.7% | 0.0% | 7.7% | 7.7% | 15.4% | 0.0% | 0.0% | 0.0% | 0.0% | 7.7% | **30.8%** | 0.0% | 0.0% | 0.0% | 0.0% | 0.0% | 23.1% |
|  | Latin, Central, and South American origins (n=18) | 11.1% | 0.0% | 11.1% | 0.0% | 11.1% | 0.0% | 5.6% | **11.1%** | 5.6% | 5.6% | 0.0% | 0.0% | 0.0% | 5.6% | 0.0% | 0.0% | 33.3% |
|  | African origins (n=17) | 11.8% | 0.0% | 5.9% | 5.9% | 11.8% | 5.9% | 5.9% | 0.0% | **11.8%** | 5.9% | 0.0% | 5.9% | 5.9% | 0.0% | 0.0% | 0.0% | 23.5% |
|  | Asian origins (n=133) | 8.3% | 3.8% | 0.8% | 8.3% | 4.5% | 3.8% | 5.3% | 4.5% | 6.8% | **5.3%** | 4.5% | 3.0% | 6.8% | 0.8% | 6.0% | 0.8% | 27.1% |
|  | Oceania origins (NR) |  |  |  |  |  |  |  |  |  |  |  |  |  |  |  |  |  |
| Generic | North American Indigenous origins (n=63) | 14.3% | 0.0% | 4.8% | 11.1% | 3.2% | 6.3% | 3.2% | 3.2% | **4.8%** | 4.8% | 3.2% | 1.6% | 1.6% | 1.6% | 3.2% | 0.0% | 33.3% |
|  | European origins (n=480) | 6.7% | 0.6% | 4.2% | 7.5% | 6.3% | 3.3% | 4.2% | 2.9% | 5.0% | 3.3% | **10.0%** | 3.1% | 8.1% | 2.3% | 7.1% | 1.5% | 24.0% |
|  | Caribbean origins (n=16) | 6.3% | 6.3% | 0.0% | 12.5% | 6.3% | 0.0% | 0.0% | 0.0% | 12.5% | **12.5%** | 0.0% | 6.3% | 6.3% | 0.0% | 0.0% | 0.0% | 31.3% |

|  | Latin, Central, and South American origins (n=16) | 12.5% | 0.0% | 12.5% | 6.3% | 0.0% | 6.3% | **12.5%** | 0.0% | 6.3% | 0.0% | 12.5% | 0.0% | 0.0% | 6.3% | 18.8% | 0.0% | 6.3% |
| --- | --- | --- | --- | --- | --- | --- | --- | --- | --- | --- | --- | --- | --- | --- | --- | --- | --- | --- |
|  | African origins (n=24) | 12.5% | 4.2% | 25.0% | **8.3%** | 4.2% | 8.3% | 0.0% | 0.0% | 8.3% | 0.0% | 4.2% | 4.2% | 0.0% | 0.0% | 8.3% | 0.0% | 12.5% |
|  | Asian origins (n=132) | 13.6% | 0.0% | 3.8% | 7.6% | 4.5% | 6.1% | 4.5% | 1.5% | 6.8% | **3.8%** | 6.8% | 3.8% | 4.5% | 2.3% | 5.3% | 1.5% | 23.5% |
|  | Oceania origins (NR) |  |  |  |  |  |  |  |  |  |  |  |  |  |  |  |  |  |
| Ethnic origin |  |  |  |  |  |  |  |  |  |  |  |  |  |  |  |  |  |  |
| Survey | Classification | PR1 | PR2 | PR3 | HM | WP1 | WP2 | WP3 | WP4 | WP5 | WP6 | WP7 | MM | E1 | E2 | E3 | E4 | E5 |
| Vaccines | Black (n=16) | 18.8% | 6.3% | 6.3% | 6.3% | 6.3% | 0.0% | **6.3%** | 0.0% | 0.0% | 6.3% | 6.3% | 6.3% | 12.5% | 0.0% | 0.0% | 0.0% | 18.8% |
|  | East Asian (n=59) | 25.4% | 0.0% | 1.7% | 5.1% | 3.4% | 5.1% | 0.0% | 0.0% | 3.4% | 3.4% | **3.4%** | 1.7% | 3.4% | 3.4% | 1.7% | 0.0% | 39.0% |
|  | Latino (n=17) | 17.6% | 0.0% | 11.8% | 5.9% | 5.9% | **11.8%** | 5.9% | 0.0% | 0.0% | 11.8% | 5.9% | 0.0% | 5.9% | 0.0% | 0.0% | 0.0% | 17.6% |
|  | Middle eastern (n=6) | 0.0% | 0.0% | 0.0% | 0.0% | 0.0% | 0.0% | 0.0% | 0.0% | 0.0% | 0.0% | 0.0% | 0.0% | 0.0% | 0.0% | 16.7% | 16.7% | **66.7%** |
|  | South Asian (n=27) | 14.8% | 3.7% | 7.4% | 7.4% | 14.8% | **3.7%** | 0.0% | 0.0% | 3.7% | 0.0% | 7.4% | 0.0% | 3.7% | 3.7% | 7.4% | 0.0% | 22.2% |
|  | Southeast Asian (n=22) | 18.2% | 0.0% | 4.5% | 18.2% | 0.0% | 4.5% | **9.1%** | 4.5% | 4.5% | 0.0% | 0.0% | 0.0% | 0.0% | 4.5% | 0.0% | 0.0% | 31.8% |
|  | White (n=564) | 20.7% | 0.7% | 2.1% | 5.0% | 4.6% | 2.7% | 2.5% | 2.1% | 4.6% | 3.0% | **5.5%** | 3.0% | 5.0% | 2.3% | 3.4% | 2.1% | 30.7% |
| Prevention | Black (n=22) | 13.6% | 0.0% | 4.5% | 13.6% | 9.1% | 4.5% | **4.5%** | 0.0% | 4.5% | 9.1% | 9.1% | 4.5% | 4.5% | 0.0% | 0.0% | 0.0% | 18.2% |
|  | East Asian (n=77) | 6.5% | 5.2% | 1.3% | 10.4% | 2.6% | 3.9% | 5.2% | 3.9% | 3.9% | 5.2% | **6.5%** | 0.0% | 7.8% | 1.3% | 6.5% | 1.3% | 28.6% |
|  | Latino (n=14) | 14.3% | 0.0% | 14.3% | 0.0% | 14.3% | 0.0% | **7.1%** | 0.0% | 7.1% | 0.0% | 0.0% | 0.0% | 0.0% | 7.1% | 0.0% | 0.0% | 35.7% |
|  | Middle eastern (n=19) | 5.3% | 5.3% | 0.0% | 15.8% | 5.3% | 0.0% | 10.5% | 5.3% | **10.5%** | 10.5% | 5.3% | 5.3% | 10.5% | 0.0% | 0.0% | 0.0% | 10.5% |
|  | South Asian (n=28) | 21.4% | 0.0% | 0.0% | 0.0% | 3.6% | 0.0% | 3.6% | 3.6% | 10.7% | **10.7%** | 7.1% | 3.6% | 0.0% | 0.0% | 3.6% | 0.0% | 32.1% |
|  | Southeast Asian (n=29) | 3.4% | 3.4% | 0.0% | 6.9% | 6.9% | 6.9% | 3.4% | 10.3% | **10.3%** | 0.0% | 3.4% | 6.9% | 6.9% | 0.0% | 10.3% | 0.0% | 20.7% |
|  | White (n=537) | 14.7% | 2.6% | 5.8% | 7.6% | 3.9% | 2.8% | 3.9% | 3.5% | 4.3% | **4.7%** | 6.9% | 3.9% | 4.1% | 2.4% | 3.2% | 1.7% | 24.0% |
| Generic | Black (n=29) | 10.3% | 3.4% | 13.8% | 6.9% | 3.4% | 10.3% | 0.0% | 0.0% | **10.3%** | 3.4% | 0.0% | 3.4% | 3.4% | 0.0% | 6.9% | 0.0% | 24.1% |
|  | East Asian (n=76) | 17.1% | 0.0% | 2.6% | 7.9% | 3.9% | 6.6% | 3.9% | 0.0% | 5.3% | **5.3%** | 7.9% | 5.3% | 6.6% | 2.6% | 5.3% | 1.3% | 18.4% |
|  | Latino (n=15) | 6.7% | 0.0% | 13.3% | 13.3% | 0.0% | 13.3% | **6.7%** | 0.0% | 6.7% | 0.0% | 13.3% | 0.0% | 0.0% | 6.7% | 13.3% | 0.0% | 6.7% |
|  | Middle eastern (n=22) | 27.3% | 4.5% | 9.1% | 4.5% | 0.0% | 0.0% | 0.0% | 0.0% | **13.6%** | 4.5% | 0.0% | 0.0% | 4.5% | 0.0% | 0.0% | 9.1% | 22.7% |
|  | South Asian (n=29) | 10.3% | 0.0% | 10.3% | 13.8% | 6.9% | 6.9% | **6.9%** | 0.0% | 3.4% | 0.0% | 6.9% | 3.4% | 3.4% | 0.0% | 3.4% | 0.0% | 24.1% |
|  | Southeast Asian (n=26) | 3.8% | 0.0% | 0.0% | 3.8% | 3.8% | 7.7% | 7.7% | 7.7% | 7.7% | 3.8% | **3.8%** | 0.0% | 0.0% | 7.7% | 3.8% | 3.8% | 34.6% |
|  | White (n=557) | 8.4% | 0.7% | 4.7% | 7.9% | 5.9% | 3.1% | 4.1% | 2.5% | 4.8% | 2.7% | **9.7%** | 3.4% | 7.4% | 2.3% | 7.0% | 1.3% | 24.1% |

Note: NR = not reported due to small cells (i.e., n<5). All results are included in the overall analysis.

## References

1. Turner HC, Archer RA, Downey LE, Isaranuwatchai W, Chalkidou K, Jit M, et al. An Introduction to the Main Types of Economic Evaluations Used for Informing Priority Setting and Resource Allocation in Healthcare: Key Features, Uses, and Limitations. Front Public Health. 2021 Aug 25;9:722927.

2. Solar O, Irwin A. A conceptual framework for action on the social determinants of health. [Internet]. World Health Organization; Available from: https://iris.who.int/bitstream/handle/10665/44489/9789241500852_eng.pdf?sequence=1

3. Cookson R, Mirelman AJ. Equity in HTA: what doesn’t get measured, gets marginalised. Isr J Health Policy Res. 2017;6:38.

4. Neumann PJ, Garrison LP, Willke RJ. The History and Future of the “ISPOR Value Flower”: Addressing Limitations of Conventional Cost-Effectiveness Analysis. Value Health. 2022 Apr;25(4):558–65.

5. Kristensen FB, Lampe K, Wild C, Cerbo M, Goettsch W, Becla L. The HTA Core Model®-10 Years of Developing an International Framework to Share Multidimensional Value Assessment. Value Health J Int Soc Pharmacoeconomics Outcomes Res. 2017 Feb;20(2):244–50.

6. Round J, Paulden M. Incorporating equity in economic evaluations: a multi-attribute equity state approach. Eur J Health Econ. 2018 May;19(4):489–98.

7. Asaria M, Griffin S, Cookson R. Distributional Cost-Effectiveness Analysis: A Tutorial. Med Decis Mak Int J Soc Med Decis Mak. 2016 Jan;36(1):8–19.

8. Cadham CJ, Prosser LA. Eliciting Trade-Offs Between Equity and Efficiency: A Methodological Scoping Review. Value Health. 2023 Jun;26(6):943–52.

9. Ward T, Mujica-Mota RE, Spencer AE, Medina-Lara A. Incorporating Equity Concerns in Cost-Effectiveness Analyses: A Systematic Literature Review. PharmacoEconomics. 2022 Jan;40(1):45–64.

10. CADTH. Guidelines for the economic evaluation of health technologies: Canada. 4th edition [Internet]. 2017. Available from: https://www.cadth.ca/guidelines-economic-evaluation-health-technologies-canada-4th-edition

11. National Advisory Commitee on Immunization. Guidelines for the economic evaluation of vaccination programs in Canada [Internet]. Government of Canada; 2023. Available from: https://www.canada.ca/en/public-health/services/immunization/national-advisory-committee-on-immunization-naci/methods-process/incorporating-economic-evidence-federal-vaccine-recommendations/guidelines-evaluation-vaccination-programs-canada.html

12. Axler R, Reddy A, Husein F, Mittmann N. Equity-Focused Health Technology Assessment at CADTH. Can J Health Technol [Internet]. 2023 Oct 18 [cited 2024 Jul 4];3(10). Available from: https://www.canjhealthtechnol.ca/index.php/cjht/article/view/757

13. Hurley J, Mentzakis E, Walli-Attaei M. Inequality aversion in income, health, and income-related health. J Health Econ. 2020 Mar;70:102276.

14. Ali S, Tsuchiya A, Asaria M, Cookson R. How Robust Are Value Judgments of Health Inequality Aversion? Testing for Framing and Cognitive Effects. Med Decis Making. 2017 Aug;37(6):635–46.

15. Sharma A, Minh Duc NT, Luu Lam, Thang T, Nam NH, Ng SJ, Abbas KS, et al. A Consensus-Based Checklist for Reporting of Survey Studies (CROSS). J Gen Intern Med. 2021 Oct;36(10):3179–87.

16. Government of Canada. Addressing inequities in infectious disease. CCDR [Internet]. 2016;42(S1). Available from: https://www.canada.ca/en/public-health/services/reports-publications/canada-communicable-disease-report-ccdr/monthly-issue/2016-42/ccdr-volume-42s-1-february-18-2016/ccdr-volume-42s-1-february-18-2016-social-determinants-health-2.html

17. AskingCanadians. AskingCanadians Panel Book: Providing Innovative Data Collection Solutions [Internet]. Sago; 2024. Available from: https://sago.com/wp-content/uploads/2024/05/Sago-AskingCanadians-Panel-Book.pdf

18. McNamara S, Holmes J, Stevely AK, Tsuchiya A. How averse are the UK general public to inequalities in health between socioeconomic groups? A systematic review. Eur J Health Econ. 2020 Mar;21(2):275–85.

19. Egleston BL, Miller SM, Meropol NJ. The impact of misclassification due to survey response fatigue on estimation and identifiability of treatment effects. Stat Med. 2011 Dec 30;30(30):3560–72.

20. Salman AA, Kopp BJ, Thomas JE, Ring D, Fatehi A. What Are the Priming and Ceiling Effects of One Experience Measure on Another? J Patient Exp. 2020 Dec;7(6):1755–9.

21. Statistics Canada. Gender and sex at birth variables - Gender of person [Internet]. 2021. Available from: https://www23.statcan.gc.ca/imdb/p3Var.pl?Function=DEC&Id=410445

22. Public Health Ontario. Collecting Information on Ethnic Origin, Race, Income, Household Size, and Language Data: A Resource for Data Collectors [Internet]. 2021. Available from: https://www.publichealthontario.ca/-/media/documents/ncov/he/2021/03/aag-race-ethnicity-income-language-data-collection.pdf?la=en

23. Statistics Canada. List of ethnic or cultural origins 2021 [Internet]. 2021. Available from: https://www23.statcan.gc.ca/imdb/p3VD.pl?Function=getVD&TVD=1310929

24. Statistics Canada. Educational attainment of person [Internet]. 2021. Available from: https://www23.statcan.gc.ca/imdb/p3Var.pl?Function=DECI&Id=1313690

25. Statistics Canada. Study: Rising prices and the impact on the most financially vulnerable: A profile of those in the bottom family income quintile [Internet]. The Daily; 2023. Available from: https://www150.statcan.gc.ca/n1/daily-quotidien/230208/dq230208a-eng.htm

26. Statistics Canada. Health-adjusted life expectancy, by sex [Internet]. 2019. Available from: https://www150.statcan.gc.ca/t1/tbl1/en/tv.action?pid=1310037001&pickMembers%5B0%5D=2.1&pickMembers%5B1%5D=4.6&cubeTimeFrame.startYear=2015+%2F+2017&cubeTimeFrame.endYear=2015+%2F+2017&referencePeriods=20150101%2C20150101

27. Robson M, Asaria M, Cookson R, Tsuchiya A, Ali S. Eliciting the Level of Health Inequality Aversion in England. Health Econ. 2017 Oct;26(10):1328–34.

28. Cookson R, Griffin S, Norheim OF, Culyer AJ, editors. Distributional Cost-Effectiveness Analysis: Quantifying Health Equity Impacts and Trade-Offs [Internet]. 1st ed. Oxford University Press; 2020 [cited 2024 Jul 16]. Available from: https://academic.oup.com/book/29892

29. Cookson R, Griffin S, Norheim OF, Culyer AJ, Chalkidou K. Distributional Cost-Effectiveness Analysis Comes of Age. Value Health. 2021 Jan;24(1):118–20.

30. Dubé E, Gagnon D, MacDonald N. Between persuasion and compulsion: The case of COVID-19 vaccination in Canada. Vaccine. 2022 Jun;40(29):3923–6.

31. Ismail SJ, Hardy K, Tunis MC, Young K, Sicard N, Quach C. A framework for the systematic consideration of ethics, equity, feasibility, and acceptability in vaccine program recommendations. Vaccine. 2020 Aug;38(36):5861–76.

